# Effect of antihypertensive drug treatment on long-term blood pressure reduction: An individual patient-level data meta-analysis of 352,744 Participants from 51 large-scale randomised clinical trials

**DOI:** 10.1101/2021.02.19.21252066

**Authors:** The Blood Pressure Lowering Treatment Trialists’ Collaboration

**Affiliations:** Individual members of working group/authorship, committees and collaborative team are listed at the end of the manuscript

**Author notes:** Corresponding author: BPLTTC, Kazem Rahimi, FRCP DM MSc FESC, Deep Medicine, NDWRH, University of Oxford, Hayes House 1F, 75 George Street, Oxford OX1 2BQ, UK.

**Keywords:** blood pressure, antihypertensive agents, randomised clinical trials, meta-analysis

## Abstract

**Objectives:** Evidence from randomised trials on long-term blood pressure (BP) reduction from pharmacologic treatment is limited. To investigate the effects of antihypertensive drugs on long-term BP change and examine its variation over time and among people with different clinical characteristics

**Design:** Individual participant-level data meta-analysis

**Setting and data source:** The Blood Pressure Lowering Treatment Trialists’ Collaboration involving 51 large-scale long-term randomised clinical trials

**Participants:** 352,744 people (42% women) with mean age of 65 years and mean baseline systolic/diastolic BP of 152/87 mmHg, of whom 18% were current smokers, 50% had cardiovascular disease, 29% had diabetes, and 72% were taking antihypertensive treatment at baseline

**Intervention:** Pharmacological BP-lowering treatment

**Outcome:** Difference in longitudinal changes in systolic and diastolic BP between randomised treatment arms over an average follow-up of four years

**Result:** Drugs were effective in lowering BP, with the maximum effect becoming apparent after 12-month follow-up and with gradual attenuation towards later years. Based on measures taken ≥12 months post-randomisation, more intense BP-lowering treatment reduced systolic/diastolic BP (95% confidence interval) by −11.2 (−11.4 to −11.0)/−5.6 (−5.8 to −5.5) mmHg than less intense treatment; active treatment by −5.1 (−5.3 to −5.0)/−2.3 (−2.4 to −2.2) mmHg lower than placebo, and active arm by −1.4 (−1.5 to −1.3)/−0.6 (−0.7 to −0.6) mmHg lower than the control arm for drug class comparison trials. BP reductions were consistently observed across a wide range of baseline BP values and ages, and by sex, history of cardiovascular disease and diabetes, and prior antihypertensive treatment use.

**Conclusion:** Pharmacological agents were effective in lowering long-term BP among individuals with a wide range of characteristics, but the net between-group reductions were modest, which is partly attributable to the intended trial goals.

## Introduction

To help address the immense burden of raised blood pressure on death and disability, clinical guidelines for managing hypertension developed in the US, Europe and other countries have invariably lowered the recommended blood pressure targets for patients at high risk of cardiovascular disease.^1–7^ These recommendations have been underpinned by evidence from large-scale randomised clinical trials (RCTs) and their meta-analyses which have shown substantial reductions in cardiovascular risk with more intensive blood pressure-lowering treatment, and largely independently of baseline blood pressure values.^8–17^ For most hypertensive patients, the lowered blood pressure targets inevitably lead to a larger gap between their usual blood pressure and the recommended target value,^18 19^ requiring more intensive pharmacological treatment.

Attributing changes in repeated measures of blood pressure of an individual patient to treatment is unreliable since measurements are subject to random fluctuations, regression to the mean, non-pharmacological effects, and other sources of variability which tend to exceed true variability in treatment response.^20–22^ To guide clinical decision-making and to help with the interpretation of changing blood pressure measurements during clinical encounters, it is necessary to have reliable information about the expected effects of drug treatment on blood pressure levels from randomised comparisons. With the growing emphasis on stratified medicine, physicians and patients alike would also be interested to know whether the expected treatment response varies according to important clinical characteristics, such as age, sex, preexisting condition, prior use of antihypertensive treatment, or baseline blood pressure value.

To date, randomised evidence on the quantitative effect of antihypertensive drugs on blood pressure has largely come from efficacy trials with usually small numbers of highly selected participants and follow-up durations of just a few weeks.^23^ Pooled evidence from these RCTs using information from individual participants’ repeated blood pressure measurements currently does not exist, which might explain why clinical practice guidelines currently provide no direct guidance on the expected magnitudes of blood pressure reduction with the various proposed treatment strategies and whether these reductions are expected to be different in particular patient groups.

To address this evidence gap, we leveraged the extended resource provided by the third and current phase of the Blood Pressure Lowering Treatment Trialists’ Collaboration (BPLTTC). This phase, which was initiated in 2013, now includes 51 trials accruing individual participant-level information from 352,744 participants with detailed information about repeated blood pressure measurements over several years.^24^ Using this resource, we conducted a meta-analysis of individual participant-level data to quantify the unconfounded effects of blood pressure-lowering drugs on long-term blood pressure, identify trial- and participant-level sources of heterogeneity, and examine the consistency of these effects across important patient subgroups.

## Methods

Details of the design of the current phase of the BPLTTC (www.bplttc.org), including the identification of eligible trials as well as data collection and harmonisation, have been described previously,^24^ with the protocol registered with PROSPERO (CRD42018099283). In brief, RCTs were eligible for inclusion if there was randomisation of patients between a blood pressure-lowering agent and a placebo arm or inactive control, between various blood pressure-lowering intensities, or between various blood pressure-lowering drugs. In addition, RCTs were required to have a minimum of 1000 person-years of follow-up in each randomly allocated arm to minimise the risk of small-study effects.^25^ Trials were not eligible if conducted exclusively in patients with heart failure, investigating short-term interventions (e.g. in acute care settings), without a drug comparison arm, or without description of randomization process. The protocol for the current analyses was reviewed and approved by the BPLTTC Steering Committee prior to data analysis.

### Comparison groups

We assigned each participant according to their random allocation in the individual trials, either to the active (or treatment) arm or to the control group (Supplementary **Table S1**). For trials that compared drug(s) with placebo, we assigned those in the drug(s) and placebo arms to active and control groups, respectively. For interventions that compared effects based on blood pressure-lowering targets (e.g. more versus less intense treatment to reduce baseline blood pressure below a pre-specified threshold or to achieve a pre-defined magnitude of reduction in blood pressure), participants allocated to treatment arms aiming to achieve greater blood pressure reduction were assigned to the active group, and those allocated to achieve less reduction to the control group. Both these placebo-controlled trials and trials comparing effects based on blood pressure reduction intensity were also collectively classified as ‘*blood pressure difference*’ trials. Trials that compared effects between different drug classes on clinical outcomes were classified as ‘*drug class comparison*’ trials. For these trials, we retrospectively assigned treatment allocations for the drug class achieving greater blood pressure reduction to the active group; where the difference was null, we assigned treatment arms to the active group for those randomised to receive the novel or newer drug class and to the control group if randomized to receive standard or usual drug class therapy.

### Outcomes

The outcomes for this study were mean systolic and diastolic blood pressure differences between active and control groups, as detailed below.

### Statistical analysis

We describe baseline characteristics of the trials and their participants, and report percentages and mean values, as appropriate. To conduct the meta-analysis of repeated blood pressure measurements over time, we used a one-stage approach,^26 27^ and applied linear mixed models to estimate the effect of treatment on blood pressure between comparison arms. We developed and compared models that accounted for clustering by trial and potential variability due to baseline blood pressure as well as other trial- and participant-level sources of heterogeneity, and determined the best fitting model for our data.^26–29^ Details of model development and selection are described in the **Supplementary Methods**. Our primary model was based on fixed treatment effect and fixed time effect but allowing for random intercepts at trial and participant levels, and a random slope for follow-up duration at participant level.

We estimated blood pressure values and their difference between comparison groups across all follow-up, and at specific time points during the follow-up, separately by trial design. As the early phase of the treatment may involve adjustments to optimise treatment regimens such as dosage titration,^30^ blood pressure difference between treatment arms may not be maximally achieved until after this period. We therefore also analysed results with and without inclusion of blood pressure measurements taken less than 12 months after randomisation. As detailed follow-up blood pressure measurements were not accessible to the collaboration for two trials,^31 32^ we used published aggregate information on achieved blood pressure for each comparison arm to estimate individual-level follow-up values (online **Supplementary Methods**).

We then investigated the effects of treatment on long-term blood pressure across important subgroups, as defined by participants’ baseline blood pressure, age, sex, history of cardiovascular disease and diabetes, and prior use of antihypertensive medication, and assessed heterogeneity in blood pressure reduction across these subgroups by comparing models with and without an interaction term for the characteristic of interest and treatment allocation. The models were adjusted for baseline blood pressure, age at recruitment and sex (except when used as stratification factors). In a sensitivity analysis, we ran analyses that excluded data from each trial.

We used likelihood ratio test (for nested models) and the Akaike information criterion (for non-nested models) to compare models, and report estimates with their 95% confidence interval and P values, which were tested at 5% significance level (two-tailed). We used R (version 3.4.4)^33^ to analyse the data.

## Results

### Characteristics of trials and participants in the BPLTTC

The 51 included trials comprised of eight blood pressure-lowering intensity trials, 21 placebo-controlled trials, and 29 drug class comparison trials (**Table 1**), mostly conducted between 1990 and 2009 (8 trials conducted after 2009). Seven trials included both comparisons between drug classes as well as either intensity of blood pressure-lowering or between active treatment and placebo. On average, the trials had four years of follow-up and eight blood pressure measurements collected after baseline.

**Table 1.** Characteristics of trials and participants.

| Characteristics | Blood pressure difference trials |  |  | Drug comparison trials | All trials |
| --- | --- | --- | --- | --- | --- |
|  | BP-lowering intensity | Placebo-controlled | All BP difference trials |  |  |
| TRIALS |  |  |  |  |  |
| No. of trials | 8 | 21 | 29 | 29 | 51<sup>†</sup> |
| No. of trials by year of end of study |  |  |  |  |  |
| Before 1990 | 0 | 2 | 2 | 0 | 2 |
| 1990 to 1999 | 2 | 7 | 9 | 7 | 14 |
| 2000 to 2009 | 2 | 10 | 12 | 19 | 27 |
| After 2009 | 4 | 2 | 6 | 3 | 8 |
| Mean (SD) trial duration (years) | 4 (2) | 4 (2) | 4 (2) | 4 (2) | 4 (2) |
| Mean (median) no. of follow-up BP measures | 14 (13) | 7 (6) | 8 (8) | 8 (7) | 8 (8) |
| PARTICIPANTS |  |  |  |  |  |
| No. of participants (% women) | 24,994 (45) | 112,934 (35) | 137,933 (37) | 224,038 (44) | 352,744 (42) |
| % (n/N) Caucasian/European ethnicity | 41 (9842 / 23,883) | 68 (58,851 / 86,908) | 62 (68,693 / 110,791) | 64 (118,128 / 185,351) | 64 (182,927 / 286,912) |
| % (N) current smoker at baseline<sup>b</sup> | 16 (3990 / 24,968) | 16 (17,702 / 111,190) | 16 (21,692 / 136,158) | 20 (44,173 / 220,708) | 18 (64,112 / 348,779) |
| Mean (SD) baseline SBP/DBP | 148 (19) / 82 (12) | 146 (21) / 83 (11) | 146 (20) / 83 (11) | 156 (21) / 90 (12) | 152 (21) / 87 (12) |
| % (N) participants by baseline SBP (mmHg) |  |  |  |  |  |
| <120 / <70 | 6 (1396) / 15 (3806) | 8 (9176) / 9 (10,037) | 8 (10,572) / 10 (13,843) | 3 (7027) / 5 (10,410) | 5 (17,050) / 7 (23,803) |
| 120 to 129 / 70 to 79 | 11 (2843) / 24 (6075) | 13 (15,063) / 24 (26,927) | 13 (17,906) / 24 (33,002) | 6 (12,969) / 14 (31,330) | 9 (30,178) / 18 (63,091) |
| 130 to 139 / 80 to 89 | 20 (4886) / 34 (8593) | 17 (19,674) / 39 (43,738) | 18 (24,560) / 38 (52,331) | 11 (23,906) / 28 (62,292) | 13 (47,303) / 32 (111,297) |
| 140 to 149 / 90 to 99 | 18 (4597) / 18 (4609) | 18 (20,590) / 21 (24,043) | 18 (25,187) / 21 (28,652) | 18 (41,220) / 30 (67,403) | 18 (64,729) / 27 (93,322) |
| 150 to 159 / 100 to 109 | 16 (4059) / 6 (1433) | 14 (16,246) / 7 (7355) | 15 (20,305) / 6 (8788) | 19 (42,509) / 18 (39,839) | 17 (60,693) / 14 (47,538) |
| ≥160 / ≥110 | 29 (7190) / 2 (452) | 28 (32,114) / 1 (750) | 29 (39,304) / 1 (1202) | 43 (95,833) / 5 (12,188) | 40 (132,134) / 4 (13,018) |
| Mean (SD) age (years) at baseline | 65 (10) | 65 (10) | 65 (10) | 65 (9) | 65 (9) |
| % (N) of participants by age at baseline |  |  |  |  |  |
| <50 years | 6 (1486) | 5 (5596) | 5 (7082) | 4 (9542) | 4 (14,858) |
| 50 to 59 years | 25 (6155) | 22 (24,668) | 22 (30,823) | 24 (52,819) | 23 (80,544) |
| 60 to 69 years | 33 (8363) | 39 (44,374) | 38 (52,737) | 39 (87,144) | 39 (136,570) |
| 70 to 79 years | 28 (6970) | 26 (28,921) | 26 (35,891) | 28 (63,119) | 28 (97,917) |
| ≥80 years | 8 (2020) | 8 (9342) | 8 (11,362) | 5 (11,369) | 6 (22,679) |
| % (N) with condition at baseline<sup>‡</sup> |  |  |  |  |  |
| Cardiovascular disease | 19 (4837/24,994) | 66 (72,209/110,020) | 57 (77,046/135,014) | 45 (98,944/221,993) | 50 (174,005/349,827) |
| Coronary heart disease | 14 (3559/24,994) | 41 (45,591/110,008) | 36 (49,150/135,002) | 38 (67,766/177,363) | 38 (115,001/305,185) |
| Stroke | 3 (692/23,900) | 34 (32,650/95,800) | 28 (33,342/119,700) | 11 (17,830/168,003) | 18 (51,046/281,619) |
| Diabetes | 31 (7768/24,994) | 36 (36,179/100,697) | 35 (43,947/125,691) | 26 (58,404/223,654) | 29 (98,603/340,417) |
| Chronic kidney disease | 33 (4854/14,799) | 9 (2845/25,789) | 19 (7699/40,588) | 17 (18,917/108,612) | 17 (24,289/145,895) |
| % (N) previously on BP-lowering medication<sup>‡</sup> | 37 (9294 / 24,994) | 71 (73,833 / 103,766) | 65 (83,127 / 128,760) | 77 (119,454 / 155,069) | 72 (199,581/275,742) |
| Mean (SD) body mass index<sup>‡</sup> (kg/m<sup>2</sup>) | 29 (6) | 28 (5) | 28 (5) | 28 (5) | 28 (5) |

BP – blood pressure; SBP – systolic blood pressure; DBP – diastolic blood pressure; <sup>†</sup>Some trials provided data to more than one trial design; <sup>‡</sup>Data limited to those with relevant information and N refers to the denominator for number of participants with information on the relevant variable.

The trials included 352,744 randomised participants (42% women) with a mean age of 65 years at baseline, including 6% aged ≥80 years. The mean baseline systolic/diastolic blood pressure was 152/87 mmHg (73% with ≥140 mmHg systolic and 44% with ≥90 mmHg diastolic blood pressure) across all trials. For blood pressure-lowering intensity, placebo-controlled and drug class comparison trials, the mean systolic/diastolic blood pressure at baseline were 148/82 mmHg, 146/83 mmHg, and 156/90 mmHg, respectively, and the proportion of participants using antihypertensive drugs at baseline were 37%, 71% and 77%, respectively. At baseline, half of all participants had had a history of cardiovascular disease and a third a history of diabetes. Baseline blood pressure was higher for older persons, in women, and among those with lower body mass index, without cardiovascular disease or diabetes, and no prior use of antihypertensive medications, as compared to their counterparts (Supplementary **Table S2**). Further details about study methods, design and risk of bias assessment for each trial are shown in Supplementary **Table S3** to **S7**.

### Temporal blood pressure patterns by treatment allocation

The temporal pattern of systolic and diastolic blood pressure in the active and control groups are shown in **Figure 1** (additional information in Supplementary **Table S8**). Across all trial designs, blood pressure initially fell during the first few months of follow-up in both study arms. For blood pressure-lowering intensity and placebo-controlled trials, blood pressure diverged in the early follow-up period, and this divergence increased over time with blood pressure levels in the active arm being lowest at around two years after baseline. For drug class comparison trials, blood pressure levels in both comparison arms remained similar across the follow-up period although the values for both comparison arms substantially fell during follow-up. The mean blood pressure achieved in the active arm of blood pressure-lowering intensity trials was substantially lower than those achieved in the active arms of the other trial designs. Results for all blood pressure difference trials are shown in Supplementary **Figure S2**.

**Figure 1.**
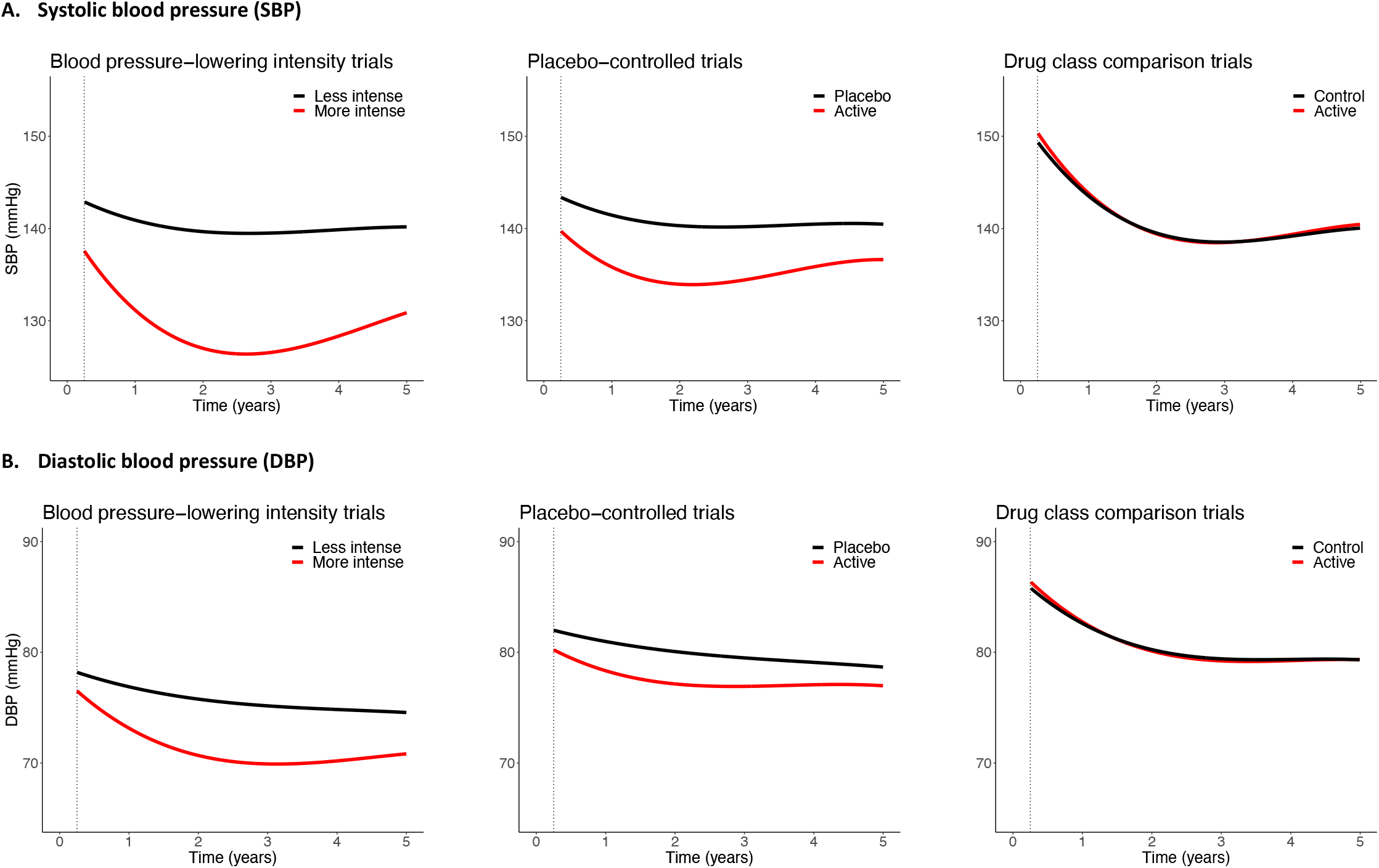
Blood pressure trajectories according to different trial designs. Results are in red for active group and black for control group, from three months to five years of follow-up. Estimates were based on separate models for treatment and control groups, with random intercepts at individual and trial levels, a random slope for time at the individual level (see Method for details) and adjusted for baseline blood pressure, age and sex. Baseline systolic/diastolic blood pressure for active and control groups were: Blood pressure-lowering trials=148/82 mmHg; Placebo-controlled trials=146/83 mmHg, and; Drug class comparison trials=156/90 mmHg. Estimated blood pressure at specific time points shown in Supplementary **Table S7**. Results for all blood pressure difference trials in Supplementary **Figure S2**.

### Achieved net blood pressure reduction by follow-up period

**Figure 2** (with additional details in Supplementary **Table S9**) illustrates the varying estimates of the difference in blood pressure between comparison groups at fixed follow-up times. Consistent with the patterns of absolute blood pressure levels for the active and control groups, the estimated difference in systolic and diastolic blood pressure achieved between these groups tended to be lower in the earlier than in the later follow-up periods. For blood pressure-lowering intensity trials, the difference in mean reductions in systolic and diastolic blood pressure within six months from baseline were −5.4 (95% CI −5.6 to −5.1) mmHg and −2.7 (95% CI −2.9 to −2.6) mmHg, respectively, and over −10-mmHg and −5-mmHg reductions, respectively, based on measures taken at later follow-up periods. Similar patterns were seen for placebo-controlled trials (and blood pressure difference trials, details shown in Supplementary **Figure S3**), albeit this group achieved smaller magnitudes in mean blood pressure reduction across all follow-up periods. Mean reductions were least for drug class comparison trials, which remained around −2/−1 mmHg systolic/diastolic blood pressure many years into the trial period.

**Figure 2.**
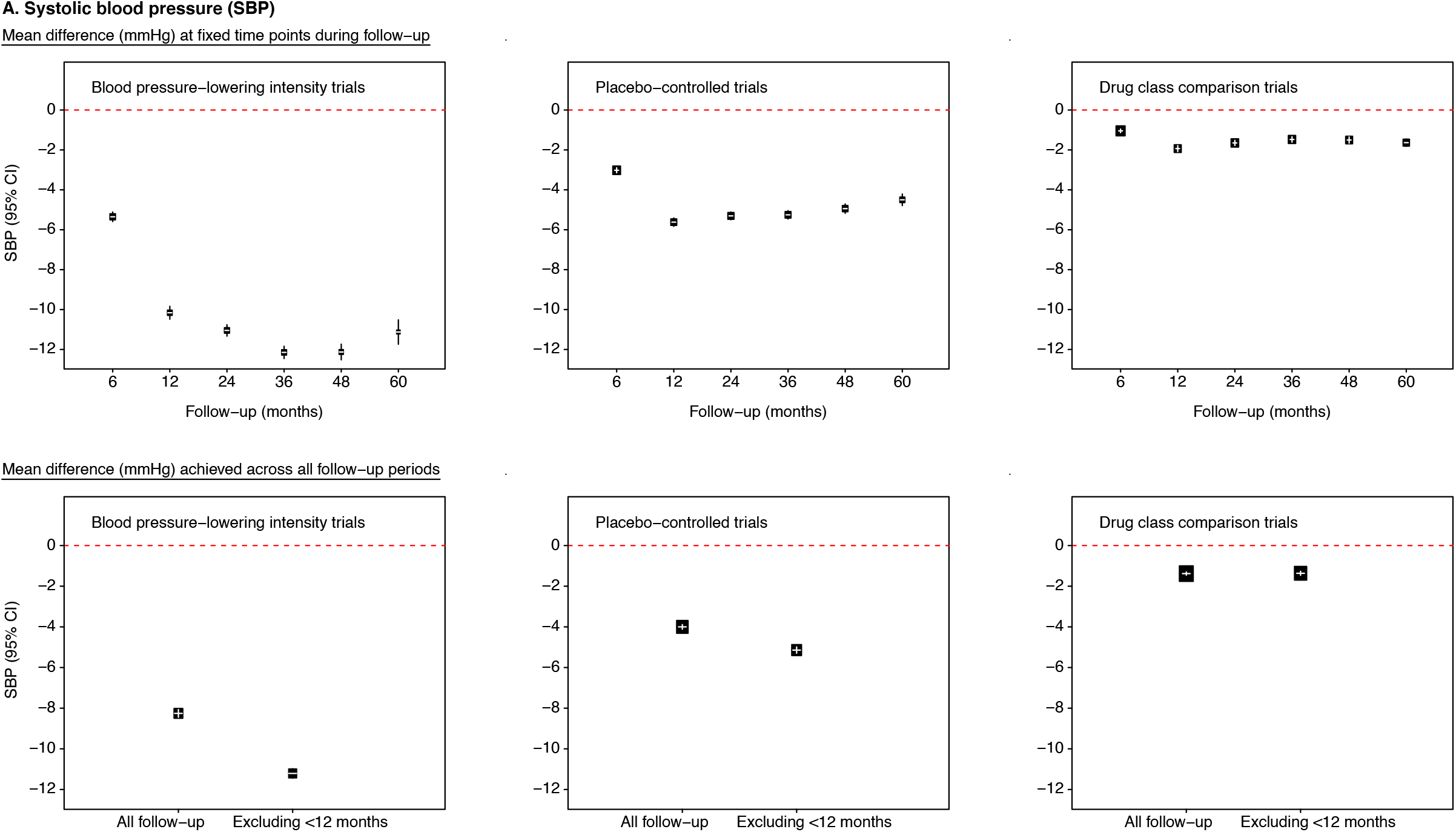
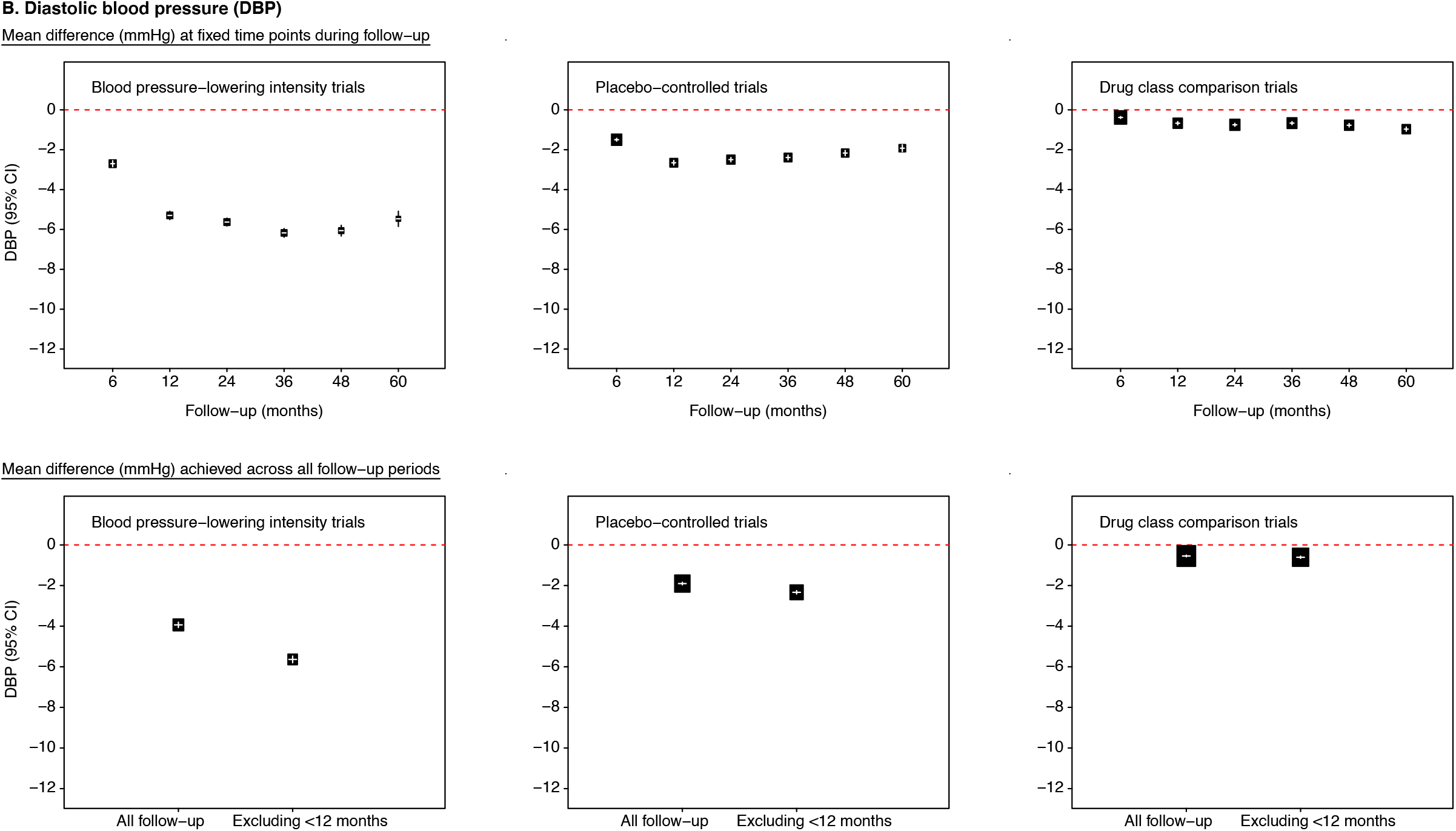
Effects of blood pressure-lowering treatment on mean blood pressure at fixed follow-up time points and across all follow-up period. For mean difference at fixed follow-up time periods, estimates were based on separate models for each time period with a fixed treatment effect and random intercept for individuals. For mean difference achieved across all time period (showing results based on all follow-up blood pressure measures and measures obtained from 12 months until end of follow-up), estimates were based on fixed treatment effect and random intercepts at individual and trial levels, a random slope for time at the individual level. All mean difference values were adjusted for baseline blood pressure, age and sex. The area of the square is inversely proportional to the variance of the estimated difference. Negative values indicate lower blood pressure in the active than in the control group. Additional information provided in Supplementary **Table S8** and **S9**, and results for all blood pressure difference trials are in Supplementary **Figure S3**.

### Estimating overall achieved blood pressure reduction between comparison groups

The time-related blood pressure differences between comparison groups affected the overall achieved reduction in blood pressure. Estimates based on blood pressure measures obtained across all follow-up period tended to be relatively smaller in magnitude than when the treatment phase of <12 months was excluded (**Figure 2** with further details in online **Figure S3** and **Table S10**). For example, for blood pressure-lowering intensity and placebo-controlled trials, the overall mean systolic and diastolic blood pressure reductions across the whole follow-up time were −8.3 (−8.4 to −8.1) mmHg and −3.9 (−4.1 to −3.8) mmHg, respectively; when estimating these differences based on follow-up measurements taken ≥12 months from baseline, achieved reductions were over −11.2 (−11.4 to −11.0) mmHg and −5.6 (−5.8 to −5.5) mmHg, respectively.

### Effects of treatment on blood pressure reduction across different subgroups

Focusing on blood pressure differences from ≥12 months from baseline, **Figure 3** (and Supplementary **Figure S4**) shows mean blood pressure differences between comparison groups by different baseline characteristics. While there were some variations in the magnitudes of blood pressure reduction across different subgroups, the effects of treatment in reducing blood pressure were evident across all the subgroups considered, even among those with baseline systolic blood pressure <120 mmHg and diastolic blood pressure <70 mmHg. For drug class comparison trials, blood pressure differences overall and across subgroups were consistently small (**Figure 4**).

**Figure 3.**
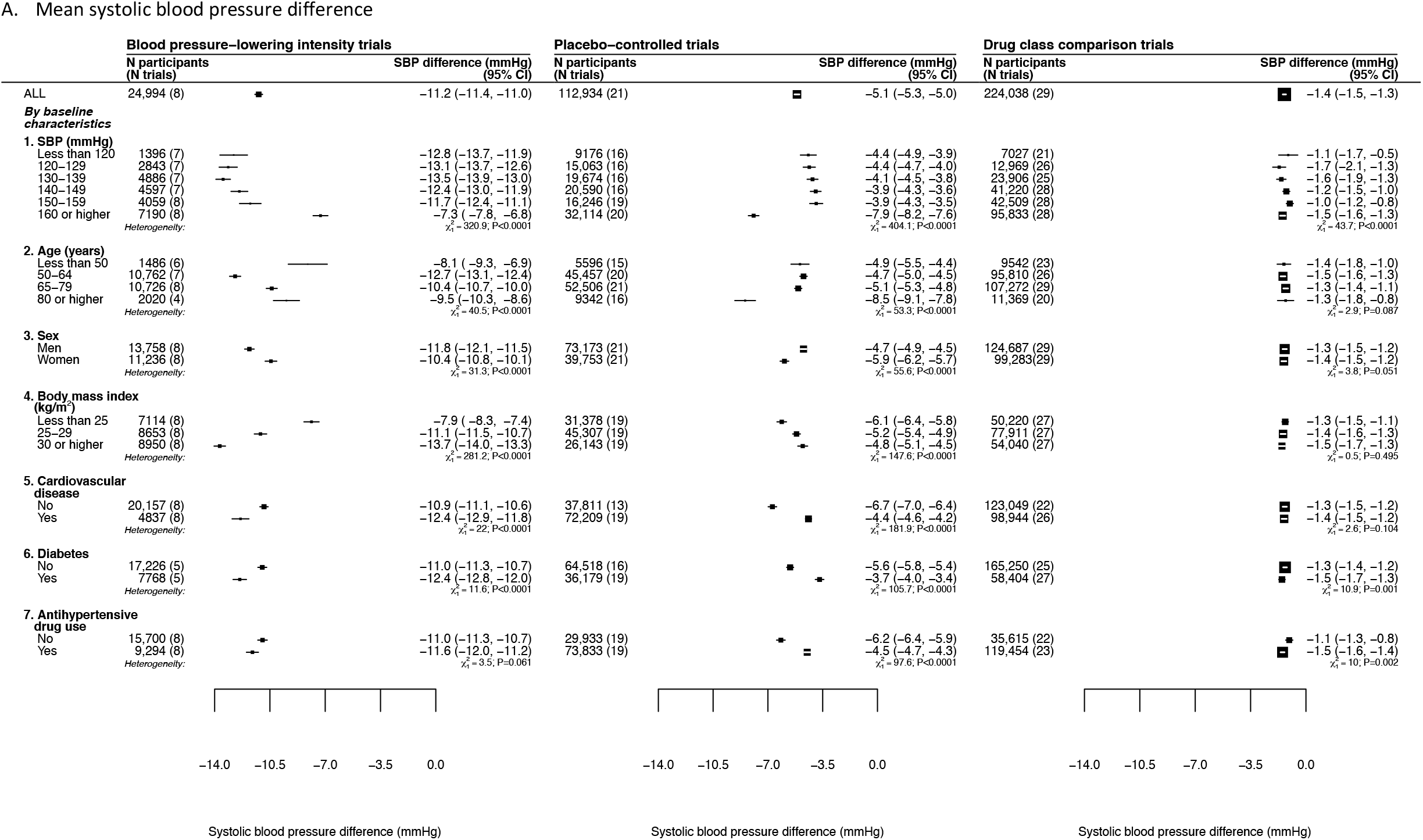
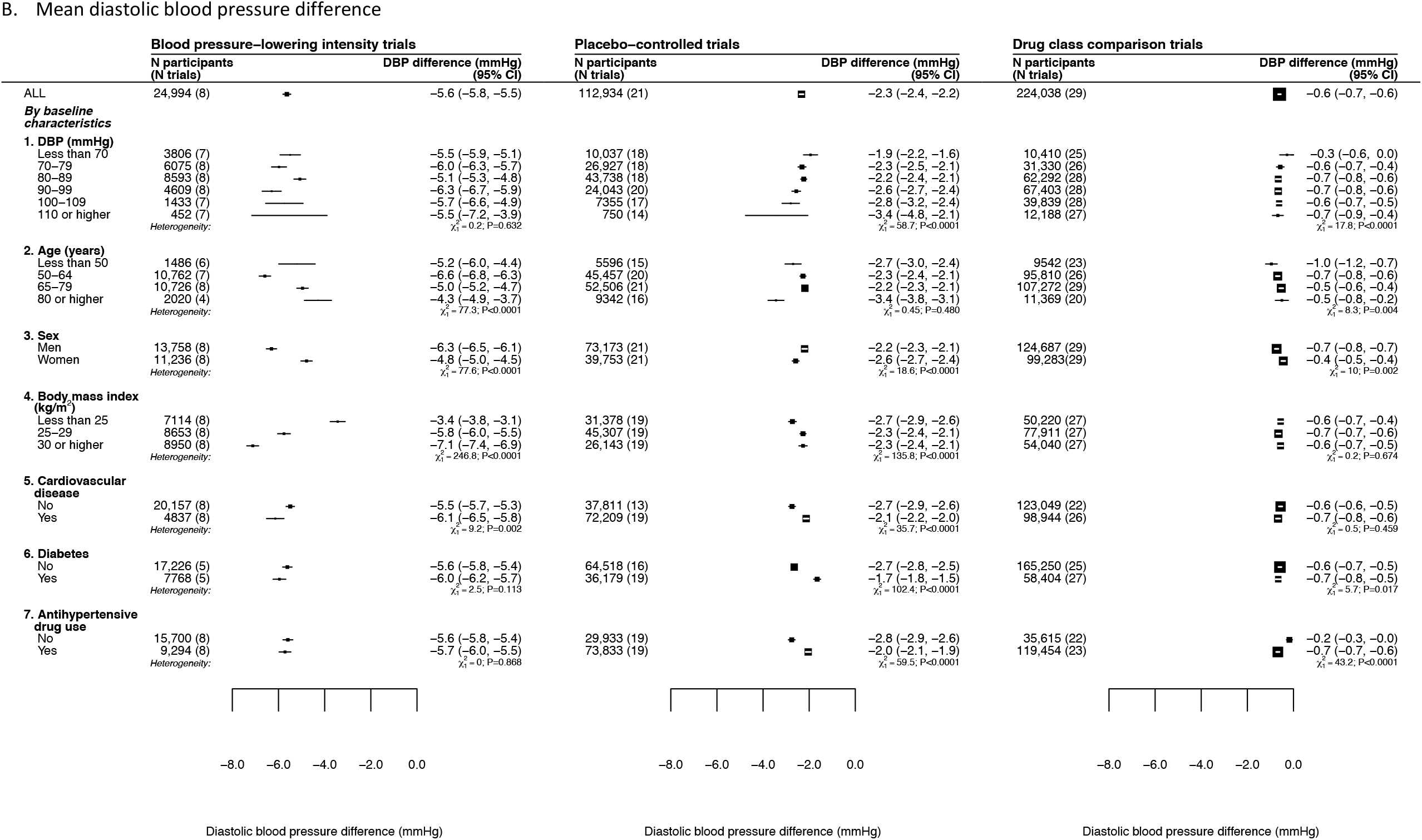
Effects of blood pressure-lowering treatment on mean blood pressure, by baseline characteristics. Estimates based on fixed treatment effect and random intercepts at individual and trial levels, a random slope for time at the individual level (see Method for details) and adjusted for baseline blood pressure, age and sex except when these variables are used as stratification factors. The area of the square is inversely proportional to the variance of the estimated difference. Negative values indicate lower blood pressure in the active than in the control group. Results for all blood pressure difference trials are in Supplementary **Figure S4**. To provide context of background blood pressure levels, baseline blood pressure by these subgroups are shown in Supplementary **Table S2**.

### Sensitivity analyses

Blood pressure differences achieved by each trial are reported in Supplementary **Figure S5**, and results after excluding one trial at a time largely showed similar results as the overall estimates within each trial type (Supplementary **Figure S6**). Further, Supplementary **Table S11** shows how our chosen final model compares with an approach that simply estimates blood pressure difference from baseline and follow-up measures, which assumes fixed effects for treatment over time (both at trial and participant levels) with random intercept at trial level. Despite the similarities in estimating blood pressure difference between comparison groups, the model that we used in our analysis fitted the data better than a model that did not take into account potential time-related variations and other individual-level factors in treatment effects.

## Discussion

This largest meta-analysis of individual-level data of 352,744 randomised participants provides detailed information on the overall and stratified effects of antihypertensive treatment on long-term blood pressure reduction from 51 large-scale RCTs. We find that the patterns of blood pressure reduction varied by time after randomisation as well as the intended trial intervention. On average, the predicted maximum effect of intervention became apparent about a year from randomisation, with some gradual attenuation many years later during follow-up. The design of the trials had a major influence on net achieved blood pressure reduction, with blood pressure-lowering intensity trials achieving the largest mean reduction of over 11-mmHg systolic blood pressure after the first year of treatment. The effects were evident largely regardless of blood pressure at baseline, and across different patient subgroups, as defined by their age, sex, body size, history of cardiovascular disease or diabetes, and prior use of antihypertensive treatment.

Prior to our study, much of the randomised evidence on the expected effect of anti-hypertensive drugs on blood pressure has been based on published information from efficacy trials.^23^ In a meta-analysis of 354 of such trials involving ≈56,000 participants, Law et al showed that, at standardised doses, the five major classes of antihypertensive drugs led to similar magnitudes of blood pressure reduction and that their combination had an additive effect in lowering blood pressure. The study suggested that at a pre-treatment systolic/diastolic blood pressure of 150/90 mmHg, half-standard dosages of one, two or three drugs could lead to systolic blood pressure reductions of 6.7 mmHg, 13.3 mmHg and 19.9 mmHg, respectively. Our study is not directly comparable with their work, but it is notable that the mean blood pressure reductions observed in our study were less pronounced than their estimates, and that full effects became evident only after a several months after initiating therapy. This discrepancy could be due to a number of reasons. In the study by Law et al., follow-up duration was relatively short (2 to 16 weeks), with some trials having potentially restricted their analysis to fully adherent participants which could have biased estimates. In contrast, we included large-scale trials with long-term follow-up, and performed analysis including all available information irrespective of treatment adherence as per intention-to-treat. Indeed, for several trials, the proportion receiving non-study antihypertension drugs were relatively higher in the placebo than in the active arm (Supplementary **Table S5**). By design, intervention strategies in many trials included in our study focused on achieving a target blood pressure level or reduction, so the maximal physiologically feasible effect on blood pressure reduction may not have been achieved. In addition, a substantial proportion of the trial participants were already on antihypertensive drugs at baseline, which could have further underestimated the magnitude of blood pressure reduction in the long-term (although this would not be expected to have an impact on the net between-group blood pressure reductions reported in our study).

Current clinical practice guidelines typically recommend a stepwise approach with gradual intensification of antihypertensive treatment over weeks and monitoring of its response for the treated individual.^1–6^ However, there is no clear guidance as to what change in blood pressure to expect when initiating treatment. To gauge treatment response based on repeated measurements of an individual without a counterfactual or ‘standard’ to compare against is difficult because of the multitude of other causes of blood pressure change.^20 21^ By providing evidence for the expected quantitative effects of antihypertensives on long-term blood pressure reduction, our study findings help mitigate exaggerated attributions of change in blood pressure to treatment while providing reassurance about achievable reductions in various groups of ‘at-risk’ patients.

Clinical practice guidelines also tend to define specific blood pressure targets that should be achieved for hypertension to be considered as ‘controlled,’ which is currently complicated by the differing target levels set by different national guidelines.^1–6^ Although setting a single target has some practical advantages, it assumes that it is achievable through full implementation of the guidelines. However, population blood pressure follows a distribution, with mean systolic blood pressure values close to 130 mmHg in Western populations, and a substantial proportion with values >140 mmHg (e.g., for adults aged ≥60 years, over 60% will have this blood pressure level).^34–36^ In our study, some of the most intensive treatment strategies in the clinical outcome studies that have shaped our evidence-base for use of antihypertensive were able to achieve an average of about 10 to 15 mmHg systolic blood pressure reductions within a few months to several years (e.g., SPRINT achieved 15 mmHg systolic blood pressure reduction [Supplementary **Figure S5**]). It would mean that with current evidence-based treatment recommendations, achieving a controlled blood pressure for many whose current blood pressure is above 150 mmHg would be difficult to attain with the trialled regimens of pharmacologic treatment.^37^ This is not to dispute that physiologically larger blood pressure reductions might be achievable but to flag the limited evidence on clinical effects of pharmacological blood pressure reductions of >20 mmHg over long term. The achieved blood pressure reduction estimated in our pooled analysis has implications not just for patients but also for health care providers whose performance will be assessed based on their patients achieving controlled blood pressure. Discussions about alternative monitoring strategies, such as the number of prescribed antihypertensives^38^ for an individual as opposed to using a single blood pressure target for all, are certainly needed.

There is also considerable controversy surrounding guideline recommendations for blood pressure management in specific patient groups. For instance, while the US guidelines have interpreted the current evidence-base to justify similar recommendations for people with our without pre-existing cardiovascular disease,^1^ the UK guidelines have kept a higher blood pressure threshold for people without cardiovascular disease due to lack of any direct evidence of efficacy in this patient group.^39^ Although there were some variations in the treatment effects in our stratified analyses according to a range of clinical characteristics at baseline, these differences were likely an artefact of trial design. Nonetheless, blood pressure reduction was evident across different subgroups that we have investigated. Our study shows that blood pressure reduction was evident across a wide range of baseline blood pressure, in younger and older adults, in women and in men. The treatment was also effective in people with and without excess weight, a history of cardiovascular disease or diabetes, and reported prior use of antihypertensive medications.

Unlike meta-analysis based on aggregate data, our individual participant-level data meta-analysis allowed us to investigate potential sources of heterogeneity in treatment effects both at trial and individual levels. In our pre-specified analyses, we stratified trials into three groups based on trial objectives and design.

Unsurprisingly, there was little difference in magnitude of blood pressure reduction between comparison arms of drug comparison trials, although blood pressure values substantially fell from baseline in both arms. Part of this reduction is likely due to regression to the mean, given the high baseline blood pressure of patients in these trials.^20^ By contrast, the other trial designs achieved greater blood pressure reduction, with some variability among different subgroups. Although clinically of little relevance, these apparent heterogeneities in treatment effects might have some implications for research, and should be accounted for, when synthesising RCT data to assess antihypertensive treatment on blood pressure-mediated disease outcomes. The extent to which these findings will have an impact on existing evidence-base, which have either been based on published information on average blood pressure differences for each trial^8^ or have not adjusted for achieved blood pressure differences between trials,^40^ requires further investigation.

A number of limitations need to be considered when interpreting our findings. Whilst we endeavoured to include as many eligible trials as possible, investigators or data custodians of some trials could not be contacted (particularly for older trials) or were unwilling to take part in the collaboration. Nevertheless, the trials included in our collaboration generally have low risk of bias. Short-term effects of blood pressure-lowering agents are well-established,^23^ and our findings extend these effects over a relatively longer period of follow-up of four years on average. However, there remains limited randomised evidence to demonstrate treatment effects beyond this period as few trials had follow up longer than five years. We were unable to compare drug classes based on standardised dosages, as most treatment interventions allowed titration or addition of other drug classes to achieve specific treatment goals. Indeed, investigators were allowed to add non-study antihypertensive treatment in some trials, which could have led to the dilution of treatment effects between trial arms or subgroups. We did not have full access to information about such non-study drugs to be able to quantify its effects. Yet an important strength of our study is that it permitted comparison across subgroups while maintaining the advantage of the random allocation to treatment groups.

Our study highlights the role of pharmacological agents in effectively reducing blood pressure over several years across individuals with a wide range of characteristics, albeit the achieved between-group reductions, even with the intensive blood pressure-lowering regimens, were relatively modest. Given that large-scale trials have shown the effects of pharmacological blood pressure reduction on improving clinical outcomes, the modest blood pressure reductions estimated in our study should still be clinically meaningful. Indeed, the estimates of long-term blood pressure reduction in this study could be helpful in setting realistic treatment goals in the pharmacologic management of raised blood pressure.

## Supporting information

PRISMA checklist

## Data Availability

Our data sharing agreements with our collaborators limit us from sharing the original data to third parties. However, a governance framework exists for collaborative projects with external research investigators.

## Authors and Working Group

Dexter Canoy, MD,^1-4^ Emma Copland, MSc,^1,2^ Milad Nazarzadeh, MSc,^1,2^ Rema Ramakrishnan, PhD,^1,2^ Ana-Catarina Pinho-Gomes, MD,^2,5,6^ Abdul Salam PhD,^4,7^ Prof Jamie Dwyer, MD,^8^ Prof Farshad Farzadfar, MD,^9^ Prof Johan Sundström, MD,^10^ Prof Mark Woodward, PhD,^11,12,13^ Prof Barry R Davis, MD,^14^ Kazem Rahimi, FRCP^1-3^

^1^ Deep Medicine, Oxford Martin School, University of Oxford, 34 Broad St., Oxford OX1 3BD, UK

^2^ Nuffield Department of Women’s and Reproductive Health, University of Oxford, Hayes House, 75 George St., Oxford OX1 2BQ, UK

^3^ NIHR Oxford Biomedical Research Centre, Oxford University Hospitals NHS Foundation Trust, Room 4503, Corridor B, Level 4, John Radcliffe Hospital, Headley Way, Headington, Oxford OX3 9DU, UK

^4^ Faculty of Medicine, University of New South Wales, Sydney NSW 2052, Australia

^5^ Department of Community Medicine, Information and Health Decision Sciences, Centre for Health Technology and Services Research, Faculty of Medicine, University of Porto, 4200 - 319 Porto, Portugal

^6^ School of Population Health & Environmental Sciences, Faculty of Life Sciences & Medicine, King’s College London, 3rd Floor, Addison House, Guy’s Campus, London, SE1 1U

^7^ The George Institute for Global Health, Nagarjuna Circle, Road No. 3, Punjagutta, Hyderabad 500082, India

^8^ Vanderbilt University Medical Center, 1211 Medical Center Drive, Nashville, TN 37232, USA

^9^ Endocrinology and Metabolism Institute, Tehran University of Medical Sciences, 2^nd^ Floor, No.10, Jalal Al-e-Ahmad Highway, Tehran, Iran

^10^ Department of Medical Sciences, Uppsala University, Akademiska sjukhuset, Entrance 40, floor 5, 751 8, Uppsala, Sweden

^11^ The George Institute for Global Health, University of New South Wales, Professorial Unit, Level 10, King George V Building, 83-117 Missenden Rd, Camperdown NSW 205, Australia

^12^ The George Institute for Global Health UK, Imperial College London, Central Working 4F, 80 Wood Lane, London W12 0BZ, UK

^13^ Department of Epidemiology, Johns Hopkins University, 615 North Wolfe Street, W6508, Baltimore, MD 21205, USA

^14^ The University of Texas School of Public Health, 1200 Pressler St. W-916, Houston, TX 77030, USA

### Steering Committee

Kazem Rahimi (Chair) (Nuffield Department of Women’s and Reproductive Health, University of Oxford, Oxford, UK), Koon Teo (Population Health Research Institute, McMaster University, Hamilton, Ontario, Canada) Barry R Davis (The University of Texas School of Public Health, Houston, Texas, USA), John Chalmers (The George Institute for Global Health, University of New South Wales, Sydney, Australia), Carl J Pepine (Department of Medicine, University of Florida, Gainesville, Florida, USA).

### Collaborating Trialists

A Adler (UKPDS [UK Prospective Diabetes Study]),

L Agodoa (AASK [African-American Study of Kidney Disease and Hypertension]),

A Algra (Dutch TIA Study [Dutch Transient Ischemic Attack Study]),

F W Asselbergs (PREVEND-IT [Prevention of Renal and Vascular End-stage Disease Intervention Trial]),

N Beckett (HYVET [Hypertension in the Very Elderly Trial]),

E Berge (deceased) (VALUE trial [Valsartan Antihypertensive Long-term Use Evaluation trial]),

H Black (CONVINCE [Controlled Onset Verapamil Investigation of Cardiovascular End Points]),

F P J Brouwers (PREVEND-IT),

M Brown (INSIGHT [International Nifedipine GITS Study: Intervention as a Goal in Hypertension]),

C J Bulpitt (EWPHE [European Working Party on High Blood Pressure in the Elderly], HYVET),

B Byington (PREVENT [Prospective Randomized Evaluation of the Vascular Effects of Norvasc Trial]),

J Chalmers (ADVANCE [Action in Diabetes and Vascular Disease: Preterax and Diamicron MR Controlled Evaluation], PROGRESS [Perindopril protection against recurrent stroke]),

W Cushman (ACCORD [Action to Control Cardiovascular Risk in Diabetes], ALLHAT, SPRINT [Systolic Blood Pressure Intervention Trial]),

J Cutler (ALLHAT [Antihypertensive and Lipid-Lowering Treatment to Prevent Heart Attack Trial]), B R Davis (ALLHAT),

R B Devereaux (LIFE [Losartan Intervention For Endpoint reduction in hypertension]), J Dwyer (IDNT [Irbesartan Diabetic Nephropathy Trial]),

R Estacio (ABCD [Appropriate Blood Pressure Control in Diabetes]),

R Fagard (SYST-EUR [SYSTolic Hypertension in EURope]),

K Fox (EUROPA [European trial on Reduction Of cardiac events with Perindopril among patients with stable coronary Artery disease]),

T Fukui (CASE-J [Candesartan Antihypertensive Survival Evaluation in Japan]),

A K Gupta (ASCOT-BPLA [Anglo-Scandinavian Cardiac Outcomes Trial – Blood Pressure Lowering Arm]), R R Holman (UKPDS),

Y Imai (HOMED-BP [Hypertension Objective Treatment Based on Measurement by Electrical Devices of Blood Pressure]),

M Ishii (JMIC-B [Japan Multicenter Investigation for Cardiovascular Diseases-B]), S Julius (VALUE),

Y Kanno (E-COST [Efficacy of Candesartan on Outcome in Saitama Trial]), S E Kjeldsen (VALUE, LIFE),

J Kostis (SHEP [Systolic Hypertension in the Elderly Program]),

K Kuramoto (NICS-EH [National Intervention Cooperative Study in Elderly Hypertensives]),

J Lanke (STOP Hypertension-2 [Swedish Trial in Old Patients with Hypertension-2], NORDIL [Nordic Diltiazem]),

E Lewis (IDNT),

J Lewis (IDNT),

M Lievre (DIABHYCAR [Non-insulin-dependent diabetes, hypertension, microalbuminuria or proteinuria, cardiovascular events, and ramipril study]),

L H Lindholm (CAPPP [Captopril Prevention Project], STOP Hypertension-2, NORDIL),

S Lueders (MOSES [The Morbidity and Mortality After Stroke, Eprosartan Compared With Nitrendipine for Secondary Prevention]),

S MacMahon (ADVANCE),

M Matsuzaki (COPE [The Combination Therapy of Hypertension to Prevent Cardiovascular Events]),

M H Mehlum (VALUE),

S Nissen (CAMELOT [Comparison of Amlodipine vs Enalapril to Limit Occurrences of Thrombosis]),

H Ogawa (HIJ-CREATE [Heart Institute of Japan Candesartan Randomized Trial for Evaluation in Coronary Heart Disease]),

T Ogihara (CASE-J, COLM [Combinations of OLMesartan], COPE),

T Ohkubo (HOMED-BP),

C Palmer (INSIGHT),

A Patel (ADVANCE),

C J Pepine (INVEST [International Verapamil SR-Trandolapril Study]),

M Pfeffer (PEACE [Prevention of Events with Angiotensin-Converting Enzyme Inhibition]),

N R Poulter (ASCOT [Anglo-Scandinavian Cardiac Outcomes Trial]),

H Rakugi (CASE-J, VALISH [Valsartan in Elderly Isolated Systolic Hypertension Study]),

G Reboldi (Cardio-Sis [CARDIOvascolari del Controllo della Pressione Arteriosa SIStolica]),

C Reid (ANBP2 [The Second Australian National Blood Pressure Study]),

G Remuzzi (BENEDICT [BErgamo NEphrologic DIabetes Complications Trial]),

P Ruggenenti (BENEDICT),

T Saruta (CASE-J),

J Schrader (MOSES), R Schrier (ABCD),

P Sever (ASCOT-BPLA),

P Sleight (CONVINCE, HOPE [Heart Outcomes Prevention Evaluation], ONTARGET [Ongoing Telmisartan Alone and in Combination with Ramipril Global Endpoint Trial],

TRANSCEND [Telmisartan Randomised AssessmeNt Study in ACE iNtolerant subjects with cardiovascular Disease]),

J A Staessen (SYST-EUR [Systolic Hypertension in Europe]),

H Suzuki (E-COST),

L Thijs (Syst-Eur),

K Ueshima (CASE-J, VALISH),

S Umemoto (COPE),

W H van Gilst (PREVEND-IT),

P Verdecchia (Cardio-Sis [CARDIOvascolari del Controllo della Pressione Arteriosa SIStolica]),

K Wachtell (LIFE),

L Wing (ANBP2),

M Woodward (ADVANCE, PROGRESS), Y Yui (JMIC-B),

S Yusuf (HOPE, ONTARGET, PRoFESS, TRANSCEND),

A Zanchetti (deceased) (ELSA [European Lacidipine Study on Atherosclerosis], VHAS [Verapamil in Hypertension and Atherosclerosis Study])

Z Y Zhang (Syst-Eur)

### Other members

C Anderson, C Baigent, BM Brenner, R Collins, D de Zeeuw, J Lubsen, E Malacco, B Neal, V Perkovic, B Pitt, A Rodgers, P Rothwell, G Salimi-Khorshidi, J Sundström, F Turnbull, G Viberti, J Wang

## Funding and support

This research was funded by the British Heart Foundation (BHF) (PG/18/65/33872 and FS/19/36/34346), the National Institute for Health Research (NIHR) Oxford Biomedical Research Centre and the Oxford Martin School. The funders had no role in the design or conduct of the study, data analysis, manuscript preparation, or approval to submit for publication The views expressed are those of the authors and not necessarily those of the National Health Service, the NIHR or the Department of Health and Social Care.

This manuscript was prepared using ACCORD, ALLHAT, PEACE and SHEP Research Materials obtained from the NHLBI Biologic Specimen and Data Repository Information Coordinating Centre and does not necessarily reflect the opinions or views of the ACCORD, ALLHAT, PEACE and SHEP, or the NHLBI. We acknowledge original depositors of the ANBP data and the Australian Data Archive, and declare that those who carried out the original analysis and collection of the data bear no responsibility for the further analysis or interpretation of the data.

## Ethics approval

The BPLTTC obtained approval to conduct this collaborative research from the Oxford Tropical Research Ethics Committee (OXTREX Reference: 545-14).

## Data sharing

The governance of BPLTTC and policies on data access and sharing policies are described elsewhere.^24^ Our data sharing agreements with our collaborators limit us from sharing the original data to third parties.

However, a governance framework exists for collaborative projects with external research investigators.

