## Supplementary material for "Effect of antihypertensive drug treatment on long-term blood pressure reduction: An individual patient-level data meta-analysis of 352,744 Participants from 51 large-scale randomised clinical trials"

**Figure S5.** Effects of blood pressure-lowering treatment on mean long-term blood pressure for each trial …………………………………………..……………………………………………………… **p 9**
**Figure S6.** Effects of blood pressure-lowering treatment on long-term mean blood pressure after excluding one trial at a time ………………………………………………………… …………………… **p 11**

**Table S9**. Supplementary data for Figure 2 and online eFigure3 showing mean blood pressure difference between comparison groups over follow-up time ………………………..…………………………. **p 37**

### Supplementary Methods

#### Model development and selection

Because the BPLTTC has individual participant-level data, we therefore made use of the full information on individual-level blood pressure measurements in the dataset for characterising longitudinal patterns of blood pressure, and estimating blood pressure difference between comparison groups. Using linear mixed models, we compared models to determine potential sources of heterogeneity, and identified the best fitting model for our data. Figure S1 shows the scheme for model specification and selection, using likelihood ratio test (for nested models) and the Akaike information criterion (for non-nested models), and implemented using the *lme* package in R. The model based on random intercepts (trial and participant), fixed treatment effect and random time (participant) fitted the data better than the base model (no treatment effect), or models with random treatment effects or fixed time effect. Further model assessment showed that the best fitting model included specifying a polynomial term for time and adjusting for baseline blood pressure, age and sex, which formed the basis for our final model as shown in Equation (1). We then implemented this model, and modified as appropriate, in subsequent analyses.

$$BP_{tij} = [\Upsilon_{000} + \Upsilon_{100}(time)_{tij}] + [r_{0ij} + r_{1i}(time)_{ti]} + \beta1(treatment) + \beta2(time) + \beta3(time)^2 + \beta4(time)^3 + e_{ij} \quad (1)$$

where:

- $i$=individual
- $j$=trial
- $BP_{tij}$ =blood pressure outcome
- $\Upsilon_{000}$=grand mean of the intercept over all individuals and group
- $\Upsilon_{100}(time)_{tij}$ =grand mean slope over all individuals and groups
- $r_{0ij}$ =random intercept (trial and individual)
- $r_{1i}(time)_{ti}$ =random slope (individual)

<u>Estimating follow-up blood pressure level and difference between comparison arms</u>

How to precisely measure differences in blood pressure between treatment arms across different trials is not clear. For example, blood pressure differences have been based on measures taken at a fixed timepoint during follow-up,[1] the average of all follow-up measures,[2-4] or the final follow-up measures.[5] In our analysis, we first examined how blood pressure differed between treatment arms across specific time points during follow-up. To estimate this blood pressure difference, we first split the data into specific time periods, and ran models on each time period separately. In developing the best-fitting model for the data used in these specific analyses, Equation (1) was modified by retaining a random intercept for individuals but excluded a random intercept for trials and random slope for time. To investigate how blood pressure differed across all follow-up periods across all participants, by patient subgroups and trial design, we used Equation (1) to estimate this difference by showing analyses that included all follow-up blood pressure measurements as well as by excluding follow-up measures taken <12 months from baseline, and modified the polynomial term for follow-up time accordingly. We have also illustrated how our model that took into account various potential sources of heterogeneity compared with a conventional approach based on simply obtaining the difference in blood pressure change from baseline and follow-up between treatment arms, which assumes fixed effects for treatment over time (both at trial and participant levels) albeit allowing for random intercept at trial level, in a sensitivity analysis.

<u>Estimating follow-up blood pressure level and difference between comparison arms in trials without follow-up measurements</u>

Mean blood pressure values at baseline for IDNT[6] were extracted from published literature for the treatment and control groups. For each participant of this trial, the baseline blood pressure was imputed as the mean value for the group they were randomized to. For both Cardio-Sis[7] and IDNT, the mean follow-up blood pressure values at the end of the trials in the treatment and control groups were extracted from published literature. These mean values were imputed as the follow-up blood pressure values for the relevant groups at one-year intervals for the length of the study duration.

For Cardio-Sis, we extracted data on the mean blood pressure difference (and 95% confidence interval) between treatment and control groups also from the published literature. As such data were not available for IDNT, we estimated these values by using simple linear regression models, with follow-up blood pressure as the outcome, and baseline blood pressure, treatment group, age and gender as covariates.

**Figure S1. Schematic diagram for model development and comparison.**

**<u>Model 0 (base model)</u>**
Random intercepts (trial and individual levels*)
No treatment effect
Fixed time effect (linear)

**Note:** **Trial and individual levels tested separately and together; all models compared with each other as well as with model without random intercepts; best fitting model included random intercepts for both trial and individual levels*

↓

**<u>Model 1</u>**
Random intercepts (trial and individual levels)
Fixed treatment effect
Fixed time effect (linear)

***Model 1 vs model 2: Treatment effect significant***

↓

**<u>Model 2</u>**
Random intercepts (trial and individual levels)
Fixed treatment effect
Fixed time effect (non-linear or polynomial* transformation for time)

***Model 2 vs 1: Cubic transformation for time significant***
**Models using quadratic and cubic transformation for time were tested separately; model that included cubic term for time fitted better than with its quadratic transformation. However, for some analyses, this polynomial time transformation was not necessary.*

↓

**<u>Model 3</u>**
Random intercepts (trial and individual levels)
Fixed treatment effect
Fixed time effect (cubic term)
Random slope for time (individual level*)

***Model 3 vs 2: Random slope for time at individual level significant***
**Models tested for trial and individual levels separately and together; best fitting model included random slope for time at individual level*

↓

**<u>Model 4</u>**
Random intercepts (trial and individual levels)
Fixed time effect (cubic term)
Random slope for time (individual-level)
Fixed treatment effect
Random slope for treatment effect

***Lack of convergence for models that included random slope for treatment effect (trial and individual levels)***

**FINAL MODEL:** ***Random intercepts (trial and individual levels), fixed treatment effect, fixed time effect (cubic term) and random slope for time (individual level) (Model 3)***

**Figure S2. Blood pressure trajectories for all blood pressure difference trials (blood pressure-lowering intensity and placebo-controlled trials combined).** Results are in red for active group and black for control group, from three months to five years of follow-up. Estimates based on separate models for treatment and control groups, with random intercepts at individual and trial levels, a random slope for time at the individual level (see ***Method*** for details) and adjusted for baseline blood pressure, age and sex. Baseline systolic/diastolic blood pressure for active and control groups was 146/82 mmHg. Estimated blood pressure at specific time points shown in online ***Table S8***.

**A. Systolic blood pressure**

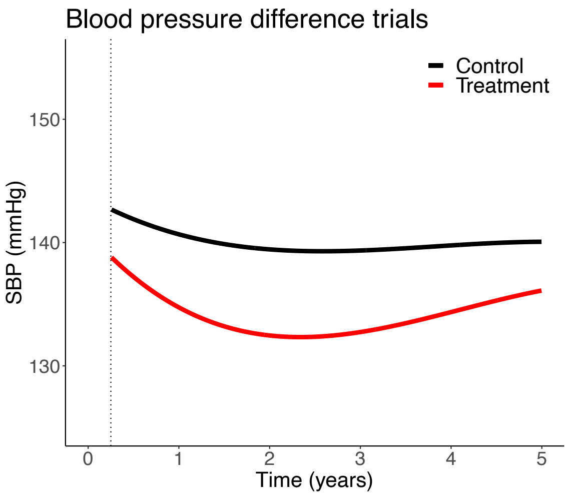

**B. Diastolic blood pressure**

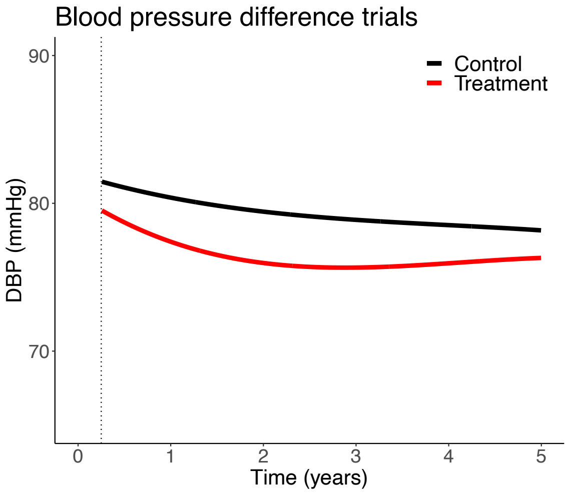

**Figure S3. Effects of blood pressure-lowering treatment on mean blood pressure at fixed follow-up time points and across all follow-up period for all blood pressure difference trials (blood pressure-lowering intensity and placebo-controlled trials combined).** For mean difference at fixed follow-up time periods, estimates based on separate models for each time period with a fixed treatment effect and random intercept for individuals. For mean difference achieved across all time period (showing results based on all follow-up blood pressure measures and measures obtained from 12 months until end of follow-up), estimates based on fixed treatment effect and random intercepts at individual and trial levels, a random slope for time at the individual level. All mean difference values were adjusted for baseline blood pressure, age and sex. The area of the square is inversely proportional to the variance of the estimated difference. Negative values indicate lower blood pressure in the active than in the control group. Additional information provided in online ***Table S9*** and ***Table S10***.

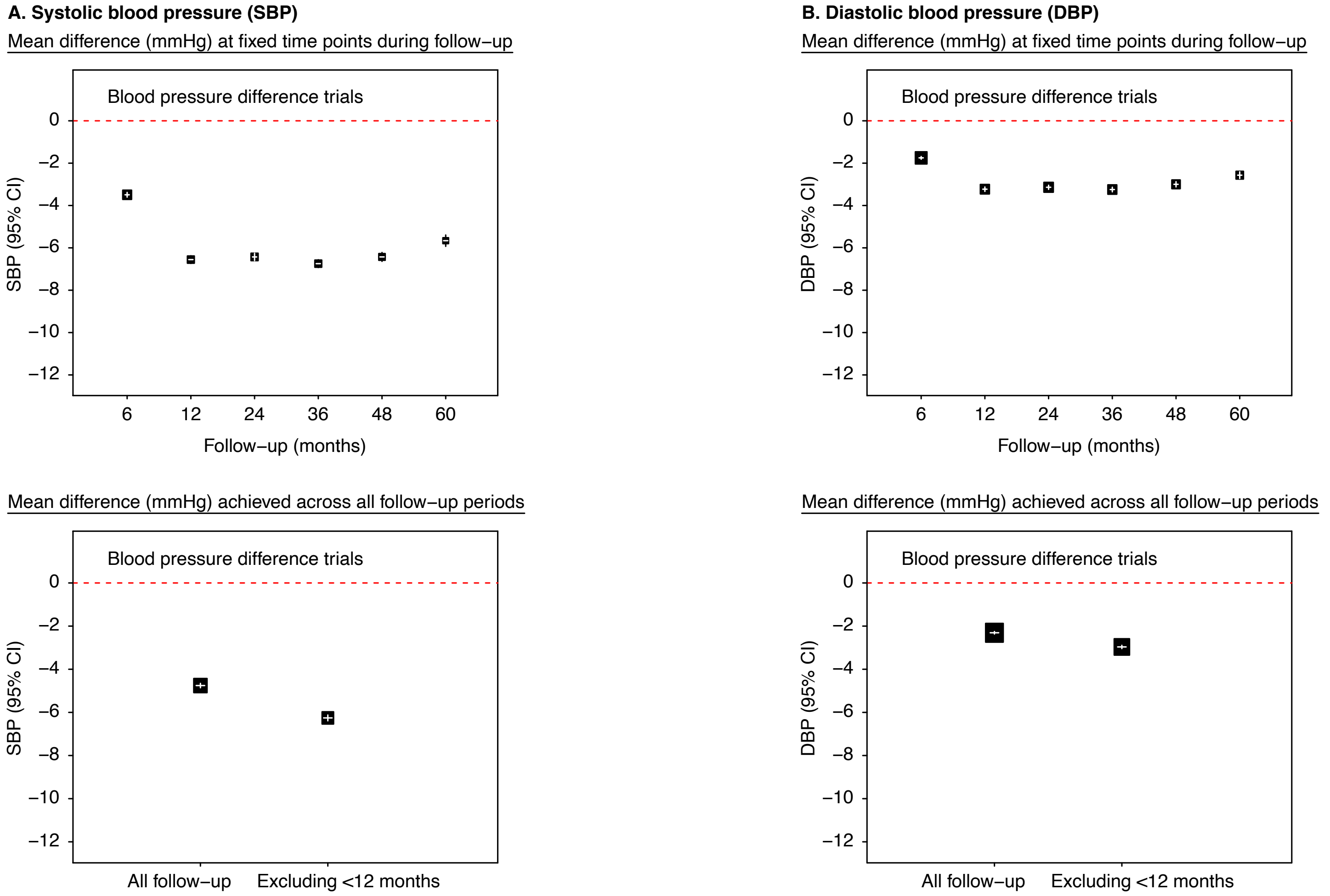

**Figure S4. Effects of blood pressure-lowering treatment on long-term mean blood pressure, by baseline characteristics in all blood pressure difference trials (blood pressure-lowering intensity and placebo-controlled trials combined**). Estimates based on fixed treatment effect and random intercepts at individual and trial levels, a random slope for time at the individual level (see ***Method*** for details) and adjusted for baseline blood pressure, age and sex except when these variables are used as stratification factors

#### A. Mean systolic blood pressure difference

| | All blood pressure difference trials | |
|---|---|---|
| | **N participants (N trials)** | **SBP difference (mmHg) (95% CI)** |
| ALL | 137,933 (29) | −6.3 (−6.4, −6.1) |
| ***By baseline characteristics*** | | |
| **1. SBP (mmHg)** | | |
| Less than 120 | 10,572 (23) | −5.7 (−6.1, −5.2) |
| 120–129 | 17,906 (23) | −5.9 (−6.2, −5.5) |
| 130–139 | 24,560 (23) | −6.2 (−6.5, −5.9) |
| 140–149 | 25,187 (23) | −5.6 (−5.9, −5.3) |
| 150–159 | 20,305 (27) | −5.2 (−5.6, −4.9) |
| 160 or higher | 39,304 (28) | −7.7 (−8.0, −7.4) |
| | *Heterogeneity:* | $\chi^2_1$ = 197.1; P<0.0001 |
| **2. Age (years)** | | |
| Less than 50 | 7082 (20) | −5.7 (−6.2, −5.2) |
| 50–64 | 56,219 (26) | −6.3 (−6.5, −6.1) |
| 65–79 | 63,232 (28) | −6.0 (−6.2, −5.8) |
| 80 or higher | 11,362 (19) | −8.5 (−9.1, −8.0) |
| | *Heterogeneity:* | $\chi^2_1$ = 15.4; P<0.0001 |
| **3. Sex** | | |
| Men | 86,934 (29) | −5.9 (−6.1, −5.7) |
| Women | 50,999 (29) | −6.9 (−7.1, −6.7) |
| | *Heterogeneity:* | $\chi^2_1$ = 50.1; P<0.0001 |
| **4. Body mass index (kg/m$^2$)** | | |
| Less than 25 | 38,492 (27) | −6.4 (−6.6, −6.1) |
| 25–29 | 53,960 (27) | −6.1 (−6.4, −5.9) |
| 30 or higher | 35,093 (27) | −7.3 (−7.6, −7.1) |
| | *Heterogeneity:* | $\chi^2_1$ = 2; P=0.156 |
| **5. Cardiovascular disease** | | |
| No | 57,968 (20) | −8.2 (−8.4, −8.0) |
| Yes | 77,046 (27) | −4.9 (−5.1, −4.7) |
| | *Heterogeneity:* | $\chi^2_1$ = 537.2; P<0.0001 |
| **6. Diabetes** | | |
| No | 81,744 (21) | −6.8 (−7.0, −6.6) |
| Yes | 43,947 (24) | −5.3 (−5.5, −5.1) |
| | *Heterogeneity:* | $\chi^2_1$ = 103.3; P<0.0001 |
| **7. Antihypertensive drug use** | | |
| No | 45,633 (27) | −7.6 (−7.9, −7.4) |
| Yes | 83,127 (27) | −5.3 (−5.5, −5.1) |
| | *Heterogeneity:* | $\chi^2_1$ = 268.8; P<0.0001 |

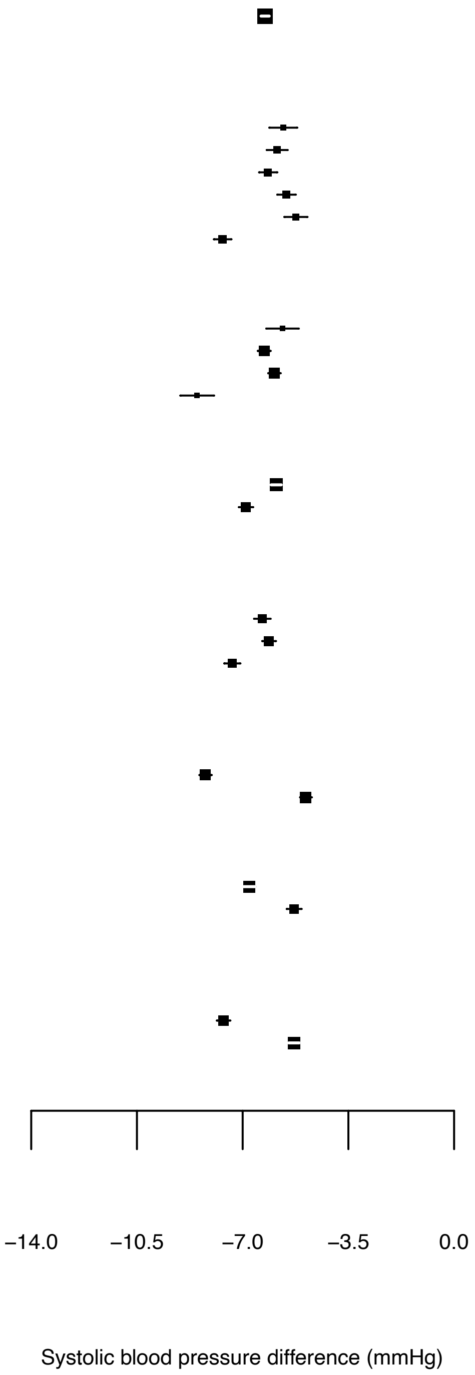

**Figure S4. Effects of blood pressure-lowering treatment on long-term mean blood pressure (cont'd)**
**B. Mean diastolic blood pressure difference**

| | All blood pressure difference trials: N participants (N trials) | All blood pressure difference trials: DBP difference (mmHg) (95% CI) |
|---|---|---|
| ALL | 137,933 (29) | −3.0 (−3.0, −2.9) |
| ***By baseline characteristics*** | | |
| **1. DBP (mmHg)** | | |
| Less than 70 | 13,843 (25) | −3.0 (−3.3, −2.7) |
| 70–79 | 33,002 (26) | −3.0 (−3.0, −2.9) |
| 80–89 | 52,331 (26) | −2.9 (−2.9, −2.6) |
| 90–99 | 28,652 (28) | −3.1 (−3.3, −2.9) |
| 100–109 | 8788 (24) | −3.6 (−4.0, −3.3) |
| 110 or higher | 1202 (21) | −4.3 (−5.4, −3.3) |
| | *Heterogeneity:* | $\chi^2_1 = 48.9$; P<0.0001 |
| **2. Age (years)** | | |
| Less than 50 | 7082 (20) | −3.3 (−3.6, −2.9) |
| 50–64 | 56,219 (26) | −3.1 (−3.3, −3.0) |
| 65–79 | 63,232 (28) | −2.7 (−2.8, −2.6) |
| 80 or higher | 11,362 (19) | −3.6 (−3.9, −3.3) |
| | *Heterogeneity:* | $\chi^2_1 = 62.5$; P<0.0001 |
| **3. Sex** | | |
| Men | 86,934 (29) | −2.9 (−3.0, −2.8) |
| Women | 50,999 (29) | −3.1 (−3.2, −3.0) |
| | *Heterogeneity:* | $\chi^2_1 = 6$; P=0.015 |
| **4. Body mass index (kg/m$^2$)** | | |
| Less than 25 | 38,492 (27) | −2.8 (−3.0, −2.7) |
| 25–29 | 53,960 (27) | −2.9 (−3.0, −2.7) |
| 30 or higher | 35,093 (27) | −3.6 (−3.8, −3.5) |
| | *Heterogeneity:* | $\chi^2_1 = 3$; P=0.082 |
| **5. Cardiovascular disease** | | |
| No | 57,968 (20) | −3.8 (−3.9, −3.6) |
| Yes | 77,046 (27) | −2.4 (−2.5, −2.3) |
| | *Heterogeneity:* | $\chi^2_1 = 277.6$; P<0.0001 |
| **6. Diabetes** | | |
| No | 81,744 (21) | −3.4 (−3.5, −3.2) |
| Yes | 43,947 (24) | −2.5 (−2.6, −2.3) |
| | *Heterogeneity:* | $\chi^2_1 = 110.3$; P<0.0001 |
| **7. Antihypertensive drug use** | | |
| No | 45,633 (27) | −3.7 (−3.8, −3.5) |
| Yes | 83,127 (27) | −2.5 (−2.6, −2.4) |
| | *Heterogeneity:* | $\chi^2_1 = 220.3$; P<0.0001 |

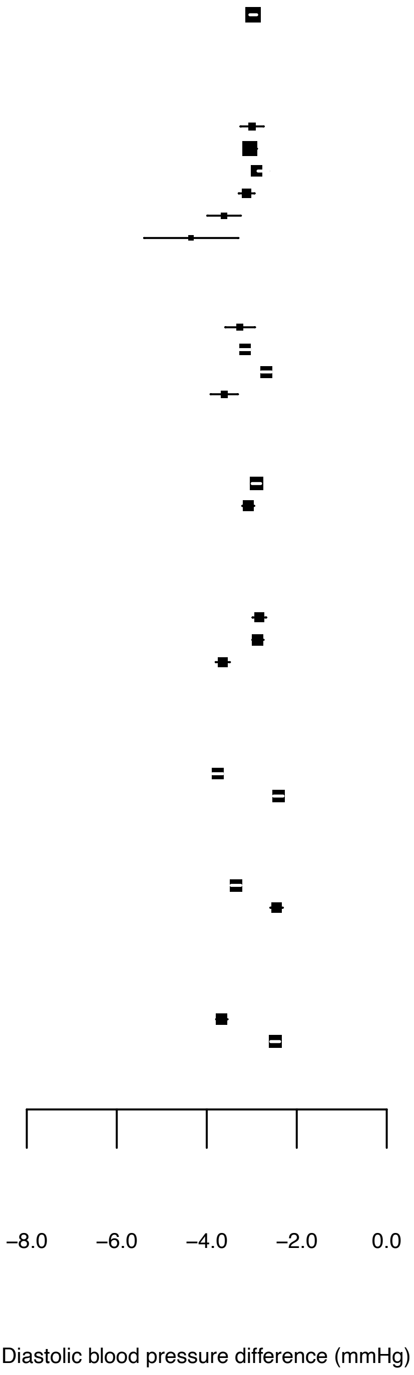

**Figure S5. Effects of blood pressure-lowering treatment on long-term blood pressure for each trial.** Estimates based on fixed treatment effect, a random intercept at the individual level, a random slope for time at the individual level (excluded from some trials due to non-convergence) (see ***Method*** for details) and adjusted for baseline blood pressure, age and sex. Acronyms described in full in ***Trial acronym legend*** in the Supplement.

**A. Mean systolic blood pressure difference**

| Trials | No. of participants Treatment / Control | Baseline SBP (SD) | SBP difference (mmHg) (95% CI) including total follow–up period | SBP difference (mmHg) (95% CI) excluding first 12 months |
|---|---|---|---|---|
| ***Placebo–controlled trials*** | | | | |
| ACTIVE I | 4518 / 4498 | 138 (17) | −2.5 ( −2.9, −2.1) | −2.6 ( −3.1, −2.0) |
| ADVANCE | 5569 / 5571 | 145 (22) | −5.4 ( −5.8, −5.1) | −5.4 ( −5.8, −4.9) |
| ANBP | 1721 / 1706 | 157 (15) | −5.5 ( −6.2, −4.9) | −7.5 ( −8.4, −6.7) |
| BENEDICT | 904 / 300 | 151 (14) | −1.5 ( −2.4, −0.6) | −0.9 ( −2.1, 0.3) |
| CAMELOT | 1336 / 655 | 129 (16) | −4.5 ( −5.7, −3.4) | −5.3 ( −6.7, −3.9) |
| DIABHYCAR | 2443 / 2469 | 145 (15) | −0.9 ( −1.5, −0.3) | −0.9 ( −1.6, −0.2) |
| Dutch TIA | 732 / 741 | 157 (25) | −2.8 ( −4.0, −1.6) | −3.1 ( −4.9, −1.4) |
| EUROPA | 6110 / 6108 | 137 (16) | −4.0 ( −4.3, −3.7) | −4.6 ( −5.0, −4.3) |
| EWPHE | 416 / 424 | 183 (17) | −14.1 (−15.6, −12.7) | −22.4 (−24.8, −20.0) |
| HOPE | 4645 / 4652 | 139 (20) | −2.5 ( −2.8, −2.1) | −3.0 ( −3.6, −2.4) |
| HYVET | 1933 / 1912 | 173 (9) | −5.8 ( −6.4, −5.2) | −13.1 (−14.1, −12.0) |
| IDNT | 1146 / 569 | 159 (20) | −1.9 ( −2.6, −1.2) | −1.9 ( −2.6, −1.2) |
| PART−2 | 308 / 309 | 133 (17) | −5.2 ( −6.5, −3.9) | −6.5 ( −8.0, −4.9) |
| PEACE | 4158 / 4132 | 133 (17) | −2.8 ( −3.1, −2.4) | −3.0 ( −3.5, −2.5) |
| PREVEND IT | 431 / 433 | 130 (18) | −6.0 ( −6.9, −5.0) | −5.6 ( −6.9, −4.3) |
| PREVENT | 417 / 408 | 129 (17) | −4.5 ( −5.5, −3.5) | −6.1 ( −7.4, −4.7) |
| PROFESS | 9873 / 9925 | 144 (17) | −3.5 ( −3.8, −3.2) | −3.4 ( −3.9, −3.0) |
| PROGRESS | 3051 / 3054 | 147 (19) | −7.2 ( −7.7, −6.8) | −9.2 ( −9.7, −8.6) |
| SHEP | 2365 / 2371 | 170 (9) | −9.7 (−10.3, −9.1) | −12.8 (−13.6, −12.1) |
| Syst−Eur | 2398 / 2297 | 174 (10) | −7.1 ( −7.6, −6.7) | −10.1 (−10.7, −9.5) |
| TRANSCEND | 2954 / 2972 | 141 (17) | −4.2 ( −4.6, −3.7) | −4.5 ( −5.1, −3.8) |
| All placebo−controlled trials | 57,430 / 55,504 | 145 (21) | −4.2 ( −4.3, −4.0) | −5.5 ( −5.7, −5.3) |
| ***BP−lowering intensity trials*** | | | | |
| AASK | 540 / 554 | 150 (24) | −9.9 (−11.1, −8.8) | −13.0 (−14.3, −11.7) |
| ABCD | 474 / 476 | 155 (17) | −4.8 ( −5.6, −4.0) | −7.7 ( −8.9, −6.6) |
| ACCORD | 2362 / 2371 | 139 (15) | −11.3 (−11.7, −10.9) | −13.9 (−14.4, −13.5) |
| Cardio−Sis | 558 / 553 | 158 (9) | −3.8 ( −5.2, −2.4) | −3.8 ( −5.2, −2.4) |
| HOMED−BP | 1759 / 1759 | 154 (18) | −1.3 ( −1.9, −0.7) | −2.0 ( −2.8, −1.1) |
| SPRINT | 4678 / 4683 | 140 (16) | −10.7 (−10.9, −10.4) | −14.9 (−15.3, −14.6) |
| UKPDS | 758 / 390 | 159 (17) | −8.7 (−10.0, −7.4) | −11.2 (−12.8, −9.7) |
| VALISH | 1545 / 1534 | 169 (9) | −3.2 ( −3.7, −2.7) | −5.0 ( −5.6, −4.4) |
| All BP−lowering intensity trials | 12,674 / 12,320 | 148 (19) | −8.2 ( −8.4, −8.0) | −11.4 (−11.6, −11.2) |
| ***Drug class comparison trials*** | | | | |
| AASK | 653 / 441 | 150 (24) | −1.7 ( −3.1, −0.4) | −1.3 ( −2.8, 0.3) |
| ABCD | 235 / 235 | 155 (17) | −0.5 ( −1.3, 0.4) | −0.8 ( −2.0, 0.5) |
| ALLHAT | 15,255 / 27,163 | 146 (16) | −1.4 ( −1.6, −1.3) | −2.0 ( −2.2, −1.7) |
| ANBP2 | 3039 / 3044 | 168 (13) | −1.1 ( −1.4, −0.7) | −0.9 ( −1.4, −0.4) |
| ASCOT−BPLA | 9639 / 9618 | 164 (18) | −3.2 ( −3.5, −2.9) | −2.2 ( −2.5, −1.9) |
| BENEDICT | 602 / 303 | 151 (14) | −1.9 ( −2.9, −1.0) | −2.0 ( −3.2, −0.8) |
| CAMELOT | 663 / 673 | 129 (16) | −0.6 ( −1.3, 0.2) | 0.1 ( −1.1, 1.4) |
| CAPPP | 5493 / 5492 | 161 (20) | −1.8 ( −2.1, −1.4) | −2.2 ( −2.6, −1.8) |
| CASE−J | 2349 / 2354 | 163 (14) | −1.4 ( −1.8, −1.0) | −1.7 ( −2.3, −1.2) |
| COLM | 2568 / 2573 | 158 (13) | −0.0 ( −0.5, 0.5) | −0.3 ( −0.9, 0.3) |
| CONVINCE | 8179 / 8297 | 150 (16) | −0.1 ( −0.3, 0.2) | 0.0 ( −0.3, 0.4) |
| COPE | 2183 / 1110 | 154 (12) | −0.2 ( −0.8, 0.4) | −0.4 ( −1.1, 0.4) |
| E−COST | 1053 / 995 | 164 (18) | −2.1 ( −2.9, −1.2) | −2.1 ( −2.9, −1.2) |
| ELSA | 1177 / 1157 | 160 (15) | −0.3 ( −1.1, 0.4) | 0.8 ( −0.1, 1.7) |
| HIJ−CREATE | 1024 / 1025 | 135 (18) | −0.6 ( −1.4, 0.2) | −0.4 ( −1.5, 0.7) |
| HOMED−BP | 2347 / 1171 | 154 (18) | −0.2 ( −0.9, 0.4) | −0.3 ( −1.2, 0.6) |
| IDNT | 579 / 567 | 160 (20) | −0.9 ( −1.3, −0.4) | −0.9 ( −1.3, −0.4) |
| INSIGHT | 3164 / 3157 | 173 (15) | −0.4 ( −0.7, −0.1) | −1.1 ( −1.5, −0.7) |
| INVEST | 10,648 / 10,672 | 151 (20) | −0.2 ( −0.5, 0.0) | −0.1 ( −0.5, 0.2) |
| JMIC−B | 828 / 822 | 146 (19) | −2.0 ( −2.9, −1.0) | −2.0 ( −3.3, −0.7) |
| LIFE | 4605 / 4588 | 174 (14) | −1.2 ( −1.6, −0.9) | −1.2 ( −1.6, −0.7) |
| MOSES | 671 / 681 | 151 (18) | −0.9 ( −1.7, −0.0) | −1.5 ( −2.7, −0.2) |
| NICS−EH | 204 / 210 | 172 (12) | −0.1 ( −1.8, 1.6) | 0.3 ( −2.0, 2.6) |
| NORDIL | 5471 / 5410 | 173 (18) | −2.2 ( −2.6, −1.9) | −3.3 ( −3.7, −2.9) |
| ONTARGET | 8502 / 17,118 | 142 (17) | −1.9 ( −2.2, −1.7) | −1.9 ( −2.3, −1.6) |
| STOP HPN−2 | 2213 / 4401 | 194 (15) | −1.7 ( −2.2, −1.2) | −2.1 ( −2.7, −1.5) |
| UKPDS | 400 / 358 | 159 (17) | −0.7 ( −2.2, 0.7) | −0.9 ( −2.7, 0.9) |
| VALUE | 7596 / 7649 | 154 (19) | −2.3 ( −2.6, −2.0) | −1.6 ( −2.0, −1.3) |
| VHAS | 707 / 707 | 169 (10) | −1.1 ( −1.7, −0.5) | −1.7 ( −2.7, −0.8) |
| All drug class comparison trials | 100,972 / 123,066 | 156 (21) | −1.3 ( −1.3, −1.2) | −1.2 ( −1.3, −1.1) |

−24.0 −17.5 −11.0 −4.5 2.0

Systolic blood pressure difference (mmHg)

−24.0 −17.5 −11.0 −4.5 2.0

Systolic blood pressure difference (mmHg)

**Figure S5. Effects of blood pressure-lowering treatment on long-term blood pressure for each trial (cont'd).**
**B. Mean diastolic blood pressure difference**

| Trials | No. of participants Treatment / Control | Baseline DBP (SD) | DBP difference (mmHg) (95% CI) including total follow-up period | DBP difference (mmHg) (95% CI) excluding first 12 months |
|---|---|---|---|---|
| ***Placebo-controlled trials*** | | | | |
| ACTIVE I | 4518 / 4498 | 82 (10) | −1.5 (−1.7, −1.2) | −1.4 ( −1.7, −1.1) |
| ADVANCE | 5569 / 5571 | 81 (10) | −2.2 (−2.4, −2.0) | −2.1 ( −2.3, −1.9) |
| ANBP | 1721 / 1706 | 100 (10) | −2.8 (−3.1, −2.4) | −3.8 ( −4.3, −3.3) |
| BENEDICT | 904 / 300 | 88 (10) | −1.3 (−1.8, −0.8) | −1.3 ( −1.9, −0.6) |
| CAMELOT | 1336 / 655 | 78 (10) | −2.8 (−3.4, −2.1) | −3.3 ( −4.2, −2.5) |
| DIABHYCAR | 2443 / 2469 | 82 (10) | −0.4 (−0.7, −0.0) | −0.4 ( −0.8, −0.1) |
| Dutch TIA | 732 / 741 | 91 (10) | −1.7 (−2.3, −1.1) | −2.0 ( −2.9, −1.2) |
| EUROPA | 6110 / 6108 | 82 (10) | −1.9 (−2.1, −1.8) | −2.2 ( −2.4, −2.0) |
| EWPHE | 416 / 424 | 101 (10) | −6.0 (−6.7, −5.3) | −9.5 (−10.6, −8.4) |
| HOPE | 4645 / 4652 | 79 (10) | −1.2 (−1.4, −1.0) | −1.4 ( −1.8, −1.1) |
| HYVET | 1933 / 1912 | 91 (10) | −2.8 (−3.1, −2.5) | −5.1 ( −5.7, −4.6) |
| IDNT | 1146 / 569 | 87 (11) | −2.8 (−3.0, −2.7) | −2.8 ( −3.0, −2.7) |
| PART−2 | 308 / 309 | 79 (10) | −2.9 (−3.6, −2.1) | −3.6 ( −4.5, −2.7) |
| PEACE | 4158 / 4132 | 77 (10) | −1.4 (−1.6, −1.2) | −1.5 ( −1.8, −1.2) |
| PREVEND IT | 431 / 433 | 76 (10) | −3.3 (−3.8, −2.8) | −2.8 ( −3.5, −2.1) |
| PREVENT | 417 / 408 | 79 (10) | −2.5 (−3.0, −1.9) | −3.3 ( −4.0, −2.6) |
| PROFESS | 9873 / 9925 | 84 (11) | −1.9 (−2.0, −1.7) | −1.8 ( −2.0, −1.6) |
| PROGRESS | 3051 / 3054 | 86 (10) | −3.2 (−3.4, −2.9) | −4.0 ( −4.3, −3.7) |
| SHEP | 2365 / 2371 | 77 (10) | −3.2 (−3.5, −2.9) | −4.2 ( −4.6, −3.9) |
| Syst−Eur | 2398 / 2297 | 86 (10) | −2.9 (−3.2, −2.7) | −4.0 ( −4.3, −3.7) |
| TRANSCEND | 2954 / 2972 | 82 (10) | −2.2 (−2.5, −2.0) | −2.2 ( −2.5, −1.8) |
| All placebo−controlled trials | 57,430 / 55,504 | 83 (11) | −1.9 (−2.0, −1.9) | −2.4 ( −2.5, −2.3) |
| ***BP−lowering intensity trials*** | | | | |
| AASK | 540 / 554 | 95 (10) | −6.5 (−7.3, −5.8) | −7.9 ( −8.6, −7.1) |
| ABCD | 474 / 476 | 98 (10) | −4.5 (−4.9, −4.1) | −6.9 ( −7.4, −6.3) |
| ACCORD | 2362 / 2371 | 76 (10) | −4.8 (−5.1, −4.6) | −6.2 ( −6.5, −5.9) |
| Cardio−Sis | 558 / 553 | 87 (10) | −1.5 (−2.4, −0.6) | −1.5 ( −2.4, −0.6) |
| HOMED−BP | 1759 / 1759 | 90 (10) | −0.5 (−0.9, −0.1) | −0.9 ( −1.5, −0.4) |
| SPRINT | 4678 / 4683 | 78 (10) | −4.8 (−5.0, −4.6) | −7.7 ( −7.9, −7.4) |
| UKPDS | 758 / 390 | 93 (10) | −4.1 (−4.8, −3.4) | −5.2 ( −6.0, −4.3) |
| VALISH | 1545 / 1534 | 82 (10) | −0.8 (−1.1, −0.5) | −1.8 ( −2.2, −1.3) |
| All BP−lowering intensity trials | 12,674 / 12,320 | 82 (12) | −3.7 (−3.9, −3.6) | −5.6 ( −5.8, −5.5) |
| ***Drug class comparison trials*** | | | | |
| AASK | 653 / 441 | 95 (10) | 0.1 (−0.7, 0.9) | 0.1 ( −0.8, 1.1) |
| ABCD | 235 / 235 | 98 (10) | 0.1 (−0.3, 0.6) | −0.2 ( −0.9, 0.5) |
| ALLHAT | 15,255 / 27,163 | 84 (10) | 0.1 (−0.0, 0.2) | 0.1 ( −0.1, 0.2) |
| ANBP2 | 3039 / 3044 | 91 (10) | −0.1 (−0.3, 0.1) | −0.0 ( −0.3, 0.2) |
| ASCOT−BPLA | 9639 / 9618 | 95 (10) | −1.8 (−1.9, −1.6) | −2.0 ( −2.1, −1.8) |
| BENEDICT | 602 / 303 | 88 (10) | −0.8 (−1.4, −0.3) | −1.0 ( −1.8, −0.3) |
| CAMELOT | 663 / 673 | 78 (10) | −0.1 (−0.6, 0.4) | 0.4 ( −0.4, 1.2) |
| CAPPP | 5493 / 5492 | 99 (10) | −1.1 (−1.2, −0.9) | −1.3 ( −1.5, −1.1) |
| CASE−J | 2349 / 2354 | 92 (10) | −0.8 (−1.1, −0.5) | −0.9 ( −1.3, −0.5) |
| COLM | 2568 / 2573 | 87 (10) | 0.4 ( 0.1, 0.7) | 0.4 ( −0.1, 0.8) |
| CONVINCE | 8179 / 8297 | 87 (10) | −0.6 (−0.7, −0.4) | −0.7 ( −0.9, −0.5) |
| COPE | 2183 / 1110 | 89 (10) | 0.3 (−0.1, 0.6) | 0.4 ( −0.2, 0.9) |
| E−COST | 1053 / 995 | 94 (10) | −0.4 (−1.0, 0.2) | −0.4 ( −1.0, 0.2) |
| ELSA | 1177 / 1157 | 98 (10) | 0.1 (−0.3, 0.5) | 0.4 ( −0.1, 0.9) |
| HIJ−CREATE | 1024 / 1025 | 76 (10) | −0.5 (−1.0, 0.0) | −0.5 ( −1.2, 0.2) |
| HOMED−BP | 2347 / 1171 | 90 (10) | 0.1 (−0.3, 0.5) | 0.2 ( −0.4, 0.8) |
| IDNT | 579 / 567 | 87 (11) | 0.0 (−0.2, 0.2) | 0.0 ( −0.2, 0.2) |
| INSIGHT | 3164 / 3157 | 99 (10) | 0.6 ( 0.4, 0.8) | 0.9 ( 0.7, 1.1) |
| INVEST | 10,648 / 10,672 | 87 (10) | −0.1 (−0.2, 0.0) | −0.2 ( −0.4, 0.0) |
| JMIC−B | 828 / 822 | 82 (10) | −1.3 (−1.9, −0.8) | −1.5 ( −2.3, −0.7) |
| LIFE | 4605 / 4588 | 98 (10) | 1.1 ( 0.9, 1.3) | 0.5 ( 0.3, 0.8) |
| MOSES | 671 / 681 | 87 (10) | −0.4 (−0.9, 0.1) | −0.5 ( −1.2, 0.1) |
| NICS−EH | 204 / 210 | 94 (10) | 0.5 (−0.5, 1.5) | 0.7 ( −0.5, 2.0) |
| NORDIL | 5471 / 5410 | 106 (10) | 0.0 (−0.1, 0.2) | −0.1 ( −0.3, 0.1) |
| ONTARGET | 8502 / 17,118 | 82 (10) | −1.1 (−1.2, −0.9) | −1.0 ( −1.2, −0.8) |
| STOP HPN−2 | 2213 / 4401 | 98 (10) | −0.7 (−0.9, −0.4) | −0.3 ( −0.6, 0.0) |
| UKPDS | 400 / 358 | 93 (10) | −1.0 (−1.8, −0.3) | −1.2 ( −2.2, −0.3) |
| VALUE | 7596 / 7649 | 87 (10) | −1.6 (−1.7, −1.4) | −1.3 ( −1.5, −1.1) |
| VHAS | 707 / 707 | 102 (10) | 0.1 (−0.2, 0.5) | 0.2 ( −0.3, 0.8) |
| All drug class comparison trials | 100,972 / 123,066 | 89 (12) | −0.5 (−0.5, −0.5) | −0.6 ( −0.7, −0.5) |

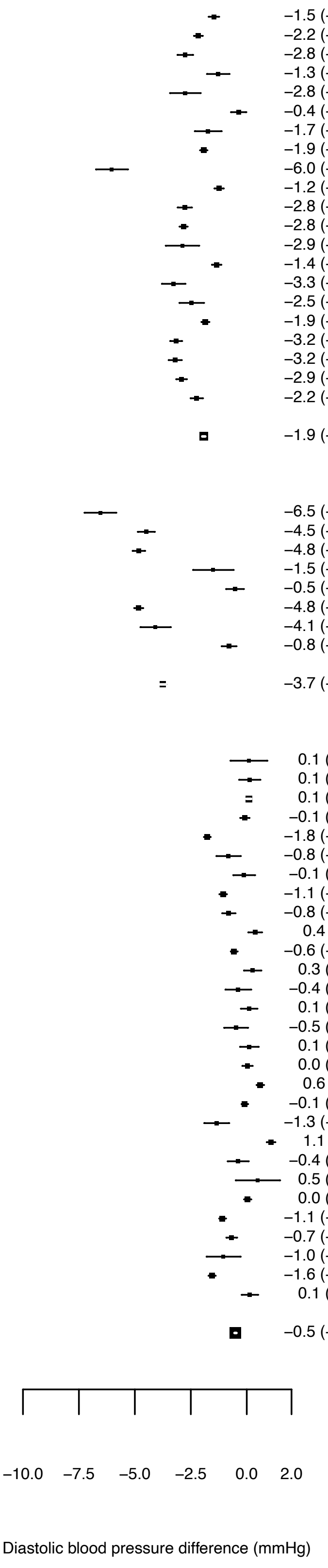

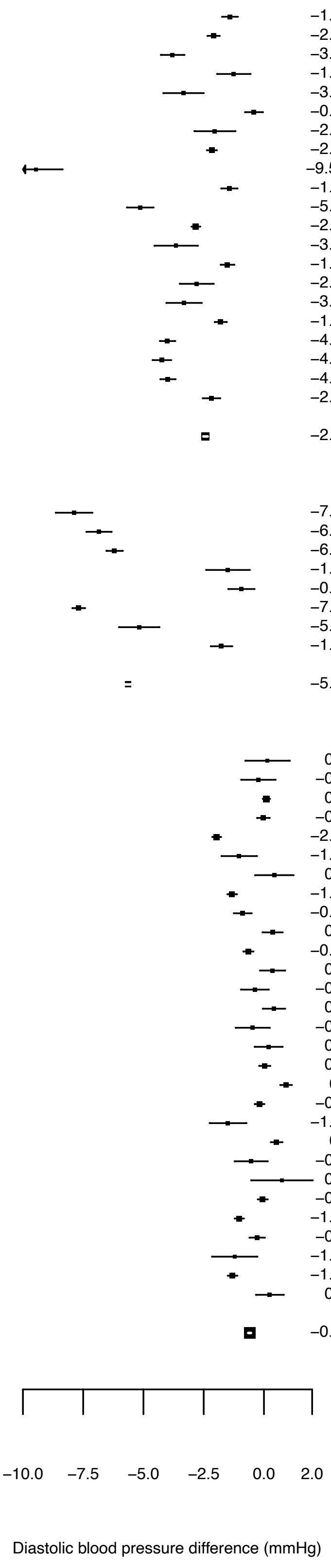

**Figure S6. Effects of blood pressure-lowering treatment on long-term mean blood pressure after excluding one trial at a time.** The estimate shown alongside a trial reflects the blood pressure difference after excluding that particular trial. Estimates based on fixed treatment effect, random intercepts at individual and trial levels, a random slope for time at the individual level (see ***Metho***d for details) and adjusted for baseline blood pressure, age and sex and using all follow-up blood pressure measurements. Acronyms described in full in ***Trial acronym legend*** in the Supplement.

| Trials | No. of participants Treatment / Control | Baseline SBP (SD) | SBP difference (mmHg) (95% CI) | Baseline DBP (SD) | DBP difference (mmHg) (95% CI) |
|---|---|---|---|---|---|
| ***Placebo–controlled trials*** | | | | | |
| ACTIVE I | 4518 / 4498 | 138 (17) | −4.2 (−4.3, −4.0) | 82 (10) | −1.9 (−2.0, −1.9) |
| ADVANCE | 5569 / 5571 | 145 (22) | −3.7 (−3.8, −3.6) | 81 (10) | −1.9 (−1.9, −1.8) |
| ANBP | 1721 / 1706 | 157 (15) | −4.0 (−4.1, −3.9) | 100 (10) | −1.9 (−2.0, −1.8) |
| BENEDICT | 904 / 300 | 151 (14) | −4.0 (−4.1, −3.9) | 88 (10) | −1.9 (−2.0, −1.8) |
| CAMELOT | 1336 / 655 | 129 (16) | −4.0 (−4.1, −3.9) | 78 (10) | −1.9 (−1.9, −1.8) |
| DIABHYCAR | 2443 / 2469 | 145 (15) | −4.1 (−4.2, −4.0) | 82 (10) | −2.0 (−2.0, −1.9) |
| Dutch TIA | 732 / 741 | 157 (25) | −4.0 (−4.1, −3.9) | 91 (10) | −1.9 (−2.0, −1.8) |
| EUROPA | 6110 / 6108 | 137 (16) | −4.0 (−4.1, −3.9) | 82 (10) | −1.9 (−2.0, −1.8) |
| EWPHE | 416 / 424 | 183 (17) | −3.9 (−4.0, −3.8) | 101 (10) | −1.9 (−1.9, −1.8) |
| HOPE | 4645 / 4652 | 139 (20) | −4.1 (−4.2, −4.0) | 79 (10) | −2.0 (−2.0, −1.9) |
| HYVET | 1933 / 1912 | 173 (9) | −3.9 (−4.0, −3.8) | 91 (10) | −1.9 (−1.9, −1.8) |
| IDNT | 1146 / 569 | 159 (20) | −4.0 (−4.1, −3.9) | 87 (11) | −1.9 (−2.0, −1.8) |
| PART–2 | 308 / 309 | 133 (17) | −4.0 (−4.1, −3.9) | 79 (10) | −1.9 (−2.0, −1.8) |
| PEACE | 4158 / 4132 | 133 (17) | −4.1 (−4.2, −4.0) | 77 (10) | −1.9 (−2.0, −1.9) |
| PREVEND IT | 431 / 433 | 130 (18) | −4.0 (−4.1, −3.9) | 76 (10) | −1.9 (−2.0, −1.8) |
| PREVENT | 417 / 408 | 129 (17) | −4.0 (−4.1, −3.9) | 79 (10) | −1.9 (−2.0, −1.8) |
| PROFESS | 9873 / 9925 | 144 (17) | −4.1 (−4.3, −4.0) | 84 (11) | −1.9 (−2.0, −1.9) |
| PROGRESS | 3051 / 3054 | 147 (19) | −3.9 (−4.1, −3.8) | 86 (10) | −1.9 (−1.9, −1.8) |
| SHEP | 2365 / 2371 | 170 (9) | −3.9 (−4.0, −3.8) | 77 (10) | −1.9 (−1.9, −1.8) |
| Syst–Eur | 2398 / 2297 | 174 (10) | −4.0 (−4.1, −3.8) | 86 (10) | −1.9 (−2.0, −1.8) |
| TRANSCEND | 2954 / 2972 | 141 (17) | −4.0 (−4.1, −3.9) | 82 (10) | −1.9 (−1.9, −1.8) |
| All placebo–controlled trials | 57,430 / 55,504 | 145 (21) | −4.0 (−4.1, −3.9) | 83 (11) | −1.9 (−2.0, −1.8) |
| ***BP–lowering intensity trials*** | | | | | |
| AASK | 540 / 554 | 150 (24) | −8.0 (−8.1, −7.8) | 95 (10) | −3.7 (−3.8, −3.5) |
| ABCD | 474 / 476 | 155 (17) | −8.4 (−8.6, −8.2) | 98 (10) | −3.9 (−4.0, −3.8) |
| ACCORD | 2362 / 2371 | 139 (15) | −7.5 (−7.7, −7.3) | 76 (10) | −3.7 (−3.9, −3.6) |
| Cardio–Sis | 558 / 553 | 158 (9) | −8.5 (−8.7, −8.3) | 87 (10) | −4.0 (−4.2, −3.9) |
| HOMED–BP | 1759 / 1759 | 154 (18) | −9.2 (−9.4, −9.1) | 90 (10) | −4.4 (−4.5, −4.3) |
| SPRINT | 4678 / 4683 | 140 (16) | −6.5 (−6.8, −6.2) | 78 (10) | −3.1 (−3.3, −3.0) |
| UKPDS | 758 / 390 | 159 (17) | −8.3 (−8.5, −8.1) | 93 (10) | −3.9 (−4.1, −3.8) |
| VALISH | 1545 / 1534 | 169 (9) | −9.0 (−9.2, −8.8) | 82 (10) | −4.4 (−4.5, −4.2) |
| All BP–lowering intensity trials | 12,674 / 12,320 | 148 (19) | −8.3 (−8.4, −8.1) | 82 (12) | −3.9 (−4.1, −3.8) |
| ***Drug class comparison trials*** | | | | | |
| AASK | 653 / 441 | 150 (24) | −1.4 (−1.5, −1.3) | 95 (10) | −0.5 (−0.6, −0.5) |
| ABCD | 235 / 235 | 155 (17) | −1.4 (−1.5, −1.3) | 98 (10) | −0.5 (−0.6, −0.5) |
| ALLHAT | 15,255 / 27,163 | 146 (16) | −1.4 (−1.5, −1.3) | 84 (10) | −0.6 (−0.7, −0.6) |
| ANBP2 | 3039 / 3044 | 168 (13) | −1.5 (−1.5, −1.4) | 91 (10) | −0.6 (−0.6, −0.5) |
| ASCOT–BPLA | 9639 / 9618 | 164 (18) | −1.1 (−1.2, −1.0) | 95 (10) | −0.4 (−0.5, −0.4) |
| BENEDICT | 602 / 303 | 151 (14) | −1.4 (−1.5, −1.3) | 88 (10) | −0.5 (−0.6, −0.5) |
| CAMELOT | 663 / 673 | 129 (16) | −1.4 (−1.5, −1.3) | 78 (10) | −0.5 (−0.6, −0.5) |
| CAPPP | 5493 / 5492 | 161 (20) | −1.4 (−1.4, −1.3) | 99 (10) | −0.6 (−0.6, −0.5) |
| CASE–J | 2349 / 2354 | 163 (14) | −1.4 (−1.5, −1.3) | 92 (10) | −0.6 (−0.7, −0.5) |
| COLM | 2568 / 2573 | 158 (13) | −1.4 (−1.5, −1.3) | 87 (10) | −0.6 (−0.7, −0.6) |
| CONVINCE | 8179 / 8297 | 150 (16) | −1.5 (−1.6, −1.4) | 87 (10) | −0.6 (−0.7, −0.5) |
| COPE | 2183 / 1110 | 154 (12) | −1.4 (−1.5, −1.3) | 89 (10) | −0.6 (−0.7, −0.6) |
| E–COST | 1053 / 995 | 164 (18) | −1.4 (−1.5, −1.3) | 94 (10) | −0.6 (−0.7, −0.6) |
| ELSA | 1177 / 1157 | 160 (15) | −1.4 (−1.5, −1.3) | 98 (10) | −0.6 (−0.7, −0.6) |
| HIJ–CREATE | 1024 / 1025 | 135 (18) | −1.4 (−1.5, −1.3) | 76 (10) | −0.6 (−0.7, −0.6) |
| HOMED–BP | 2347 / 1171 | 154 (18) | −1.4 (−1.5, −1.3) | 90 (10) | −0.6 (−0.7, −0.6) |
| IDNT | 579 / 567 | 160 (20) | −1.4 (−1.5, −1.3) | 87 (11) | −0.6 (−0.7, −0.6) |
| INSIGHT | 3164 / 3157 | 173 (15) | −1.4 (−1.5, −1.4) | 99 (10) | −0.6 (−0.7, −0.5) |
| INVEST | 10,648 / 10,672 | 151 (20) | −1.5 (−1.6, −1.4) | 87 (10) | −0.6 (−0.7, −0.6) |
| JMIC–B | 828 / 822 | 146 (19) | −1.4 (−1.5, −1.3) | 82 (10) | −0.6 (−0.7, −0.5) |
| LIFE | 4605 / 4588 | 174 (14) | −1.4 (−1.5, −1.3) | 98 (10) | −0.7 (−0.7, −0.6) |
| MOSES | 671 / 681 | 151 (18) | −1.4 (−1.5, −1.3) | 87 (10) | −0.6 (−0.7, −0.6) |
| NICS–EH | 204 / 210 | 172 (12) | −1.4 (−1.5, −1.3) | 94 (10) | −0.6 (−0.7, −0.6) |
| NORDIL | 5471 / 5410 | 173 (18) | −1.3 (−1.4, −1.2) | 106 (10) | −0.6 (−0.7, −0.6) |
| ONTARGET | 8502 / 17,118 | 142 (17) | −1.3 (−1.4, −1.2) | 82 (10) | −0.5 (−0.6, −0.5) |
| STOP HPN–2 | 2213 / 4401 | 194 (15) | −1.4 (−1.4, −1.3) | 98 (10) | −0.6 (−0.7, −0.6) |
| UKPDS | 400 / 358 | 159 (17) | −1.4 (−1.5, −1.3) | 93 (10) | −0.6 (−0.7, −0.6) |
| VALUE | 7596 / 7649 | 154 (19) | −1.3 (−1.4, −1.2) | 87 (10) | −0.5 (−0.6, −0.5) |
| VHAS | 707 / 707 | 169 (10) | −1.4 (−1.5, −1.3) | 102 (10) | −0.6 (−0.7, −0.6) |
| All drug class comparison trials | 100,972 / 123,066 | 156 (21) | −1.4 (−1.5, −1.3) | 89 (12) | −0.5 (−0.6, −0.5) |

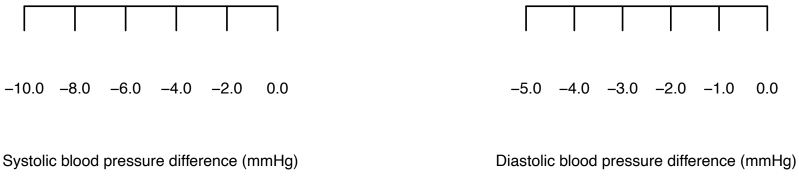

**Table S1**. **List of trials and interventions assigned to the active and control arms in the analyses**.

**A. Blood pressure difference trials**

| **Trial** | **Active group** | **Control group** |
|---|---|---|
| **Trials comparing blood pressure-lowering targets (8 trials)** | | |
| AASK | More intense treatment | Less intense treatment |
| ABCD | More intense treatment | Less intense treatment |
| ACCORD | More intense treatment | Less intense treatment |
| CARDIO-SIS | More intense treatment | Less intense treatment |
| HOMED-BP | More intense treatment | Less intense treatment |
| SPRINT | More intense treatment | Less intense treatment |
| UKPDS | More intense treatment | Less intense treatment |
| VALISH | More intense treatment | Less intense treatment |
| **Blood pressure-lowering trials comparing treatment versus placebo (21 trials)** | | |
| ACTIVE-I | ARB | Placebo |
| ADVANCE | ACEI and diuretic | Placebo |
| ANBP | Diuretic | Placebo |
| BENEDICT | ACEI, CCB and ACEI/CCB | Placebo |
| CAMELOT | CCB and ACEI | Placebo |
| DIABHYCAR | ACEI | Placebo |
| DUTCH-TIA | ß-blocker | Placebo |
| EUROPA | ACEI | Placebo |
| EWPHE | Diuretic | Placebo |
| HOPE | ACEI | Placebo |
| HYVET | Diuretic | Placebo |
| IDNT | ARB and CCB | Placebo |
| PART 2 | ACEI | Placebo |
| PEACE | ACEI | Placebo |
| PREVEND IT | ACEI | Placebo |
| PREVENT | CCB | Placebo |
| PROFESS | ARB | Placebo |
| PROGRESS | ACEI and/or diuretic | Placebo |
| SHEP | ß-blocker and diuretic | Placebo |
| SYST-EUR | CCB | Placebo |
| TRANSCEND | ARB | Placebo |

**Table S1. List of trials and interventions assigned to the active and control arms in the analyses (cont'd).**

**B. Drug comparison trials (29 trials)**

| Trial | Active group | Control group |
|---|---|---|
| AASK | ACEI and CCB | ß-blocker |
| ABCD | CCB | ACEI |
| ALLHAT | Diuretic | ACEI, CCB and $\alpha$-blocker |
| ANBP2 | Diuretic | ACEI |
| ASCOT-BPLA | CCB-based | ß-blocker-based |
| BENEDICT | ACEI and ACEI/CCB | CCB |
| CAMELOT | CCB | ACEI |
| CAPPP | ß-blocker and/or diuretic | ACEI |
| CASE-J | CCB | ARB |
| COLM | ARB and diuretic | ARB and CCB |
| CONVINCE | CCB | ß-blocker or diuretic |
| COPE | CCB/diuretic and CCB/ ß-blocker | CCB and ARB |
| E-COST | ARB | Conventional |
| ELSA | CCB | ß-blocker |
| HIJ-CREATE | ARB | non-ARB |
| HOMED-BP | CCB | ACEI and ARB |
| IDNT | ARB | CCB |
| INSIGHT | Diuretic | CCB |
| INVEST | CCB | non-CCB |
| JMIC-B | CCB | ACEI |
| LIFE | ARB | ß-blocker |
| MOSES | CCB | ARB |
| NICS-EH | Diuretic | CCB |
| NORDIL | ß-blocker and/or diuretic | CCB |
| ONTARGET | ARB/ACEI | ACEI and ARB |
| STOP Hypertension-2 | ß-blocker and/or diuretic | ACEI and CCB |
| UKPDS | ß-blocker | ACEI |
| VALUE | CCB-based | ARB-based |
| VHAS | Diuretic | CCB |

Acronyms are described in full in the Trial acronym legend in the Supplement; CCB – calcium channel-blocker; ACEI – angiotensin-converting enzyme inhibitor; ARB – angiotensin II receptor blocker.

**Table S2. Mean baseline blood pressure according to baseline characteristics and trial designs**.

A. Systolic blood pressure (mmHg)

| | **Blood pressure-lowering intensity trials** | **Placebo-controlled trials** | **All blood pressure difference trials** | **Drug class comparison trials** |
|---|---|---|---|---|
| | Mean (SD) | Mean (SD) | Mean (SD) | Mean (SD) |
| **ALL PARTICIPANTS** | **148 (19)** | **146 (20)** | **146 (20)** | **156 (21)** |
| By baseline characteristics | | | | |
| 1. Systolic blood pressure (mmHg) | | | | |
| <120 | 113 (6) | 112 (6) | 112 (6) | 112 (6) |
| 120 to 129 | 125 (3) | 124 (3) | 124 (3) | 124 (3) |
| 130 to 139 | 135 (3) | 133 (3) | 134 (3) | 134 (3) |
| 140 to 149 | 144 (3) | 143 (3) | 143 (3) | 144 (3) |
| 150 to 159 | 154 (3) | 153 (3) | 154 (3) | 154 (3) |
| ≥160 | 171 (9) | 171 (11) | 171 (11) | 175 (14) |
| 2. Age (years) | | | | |
| <50 | 146 (21) | 138 (18) | 139 (19) | 155 (19) |
| 50 to 65 | 142 (17) | 142 (19) | 142 (19) | 154 (20) |
| 65 to 79 | 149 (19) | 147 (20) | 148 (20) | 157 (21) |
| ≥80 | 154 (19) | 160 (21) | 159 (21) | 160 (23) |
| 3. Sex | | | | |
| Men | 145 (18) | 143 (19) | 143 (19) | 154 (20) |
| Women | 150 (19) | 151 (21) | 151 (21) | 158 (21) |
| 4. Body mass index (kg/$m^2$) | | | | |
| <25 | 155 (19) | 147 (21) | 148 (21) | 158 (22) |
| 25 to 30 | 147 (18) | 146 (20) | 146 (20) | 155 (21) |
| ≥30 | 142 (18) | 146 (20) | 145 (19) | 157 (21) |
| 5. Cardiovascular disease history | | | | |
| No | 148 (19) | 154 (21) | 152 (21) | 161 (20) |
| Yes | 144 (19) | 141 (19) | 141 (19) | 150 (20) |
| 6. Diabetes history | | | | |
| No | 149 (19) | 147 (22) | 148 (21) | 157 (21) |
| Yes | 145 (18) | 146 (19) | 146 (19) | 153 (19) |
| 7. Antihypertensive drug use | | | | |
| No | 147 (19) | 149 (21) | 148 (21) | 163 (19) |
| Yes | 149 (19) | 144 (20) | 144 (20) | 151 (20) |

**Table S2. Mean baseline blood pressure according to baseline characteristics and trial designs (cont'd).**

B. Diastolic blood pressure (mmHg)

| | Blood pressure-lowering intensity trials | Placebo-controlled trials | All blood pressure difference trials | Drug class comparison trials |
|---|---|---|---|---|
| | Mean (SD) | Mean (SD) | Mean (SD) | Mean (SD) |
| **ALL PARTICIPANTS** | **82 (12)** | **83 (11)** | **83 (11)** | **90 (12)** |
| By baseline characteristics | | | | |
| 1. Diastolic blood pressure (mmHg) | | | | |
| <70 | 63 (5) | 63 (5) | 63 (5) | 64 (5) |
| 70 to 79 | 75 (3) | 74 (3) | 74 (3) | 75 (3) |
| 80 to 89 | 84 (3) | 83 (3) | 83 (3) | 84 (3) |
| 90 to 99 | 94 (3) | 93 (3) | 93 (3) | 94 (3) |
| 100 to 109 | 103 (3) | 102 (3) | 103 (3) | 103 (3) |
| ≥110 | 116 (7) | 115 (7) | 115 (7) | 113 (5) |
| 2. Age (years) | | | | |
| <50 | 95 (13) | 87 (12) | 88 (12) | 99 (11) |
| 50 to 65 | 84 (11) | 84 (11) | 84 (11) | 92 (12) |
| 65 to 79 | 78 (11) | 81 (10) | 81 (11) | 87 (12) |
| ≥80 | 74 (11) | 84 (11) | 82 (12) | 84 (12) |
| 3. Sex | | | | |
| Men | 82 (13) | 83 (11) | 83 (11) | 90 (12) |
| Women | 82 (12) | 83 (11) | 83 (11) | 90 (12) |
| 4. Body mass index (kg/m$^2$) | | | | |
| <25 | 83 (12) | 82 (11) | 82 (11) | 89 (12) |
| 25 to 30 | 82 (13) | 83 (10) | 83 (11) | 90 (12) |
| ≥30 | 82 (13) | 84 (11) | 83 (11) | 90 (12) |
| 5. Cardiovascular disease history | | | | |
| No | 83 (12) | 84 (11) | 84 (12) | 93 (12) |
| Yes | 79 (14) | 82 (10) | 82 (11) | 86 (12) |
| 6. Diabetes history | | | | |
| No | 82 (13) | 84 (11) | 83 (12) | 91 (12) |
| Yes | 81 (12) | 82 (10) | 82 (11) | 86 (11) |
| 7. Antihypertensive drug use | | | | |
| No | 82 (12) | 84 (11) | 83 (12) | 95 (11) |
| Yes | 82 (12) | 82 (11) | 82 (11) | 87 (12) |

**Table S3. Characteristics of trials included in the BPLTTC study.**

A. Trials comparing different blood pressure-lowering targets intensity

| Trial | Setting | Inclusion criteria | Exclusion criteria | Recruitment period | Randomization groups, No. of participants (% women) | | | Treatment goals for blood pressure control | | Treatment | Follow-up duration (y) | No. of follow-up BP measures |
|---|---|---|---|---|---|---|---|---|---|---|---|---|
| | | | | | All | More intense | Less intense | More intense | Less intense | | | |
| AASK[8 9] | USA | Age 18-70 years, African-American, hypertension, renal disease (GFR=20-65 ml/min per 1.73$m^2$) | DBP <95 mmHg, diabetes, urine protein:creatinine ratio >25, recent malignant hypertension, secondary hypertension, non-blood pressure-related CKD, serious systemic disease, heart failure | Feb 1995 to Sept 1998 | 1094 (39) | 540 (38) | 554 (40) | Mean arterial pressure ≤92 mmHg | Mean arterial pressure 102-107 mmHg | 1 of 3: $\beta$-blocker (Metoprolol), ACEI (Ramipril), CCB (Amlodipine); Plus furosemide, doxazosin, clonidine, and hydralazine or minoxidil sequentially | 4.8 | 54 |
| ABCD[10-12] | USA | Age 0-74 years, with T2D, DBP ≥80 mmHg, not on antihypertensive treatment | Recent CAD or CeVD, heart failure, renal disease | Mar 1991 to May 1993 | 950 (39) | 474 (0.4) | 476 (38) | DBP <75 mmHg for hypertensives (DBP ≥90 mmHg) or DBP reduction by 10 mmHg for normotensives | DBP 80-89 mmHg for hypertensives (DBP ≥90 mmHg) or no change for normotensives | Hypertensive group: CCB (Nisoldipine) and ACEI (Enalapril); plus $\beta$-blocker (Metoprolol), diuretic (HCTZ), or others but not CCB or ACEI; Normotensive group: CCB (Nisoldipine), ACEI (Enalapril) or placebo | 4.7 | 8 |
| ACCORD[13] | USA and Canada | Age ≥40y years with CVD or ≥50 years with substantial atherosclerosis, T2D, HbA1c ≥7.5%, albuminuria, LVH or ≥2 CVD risk factors (dyslipidaemia, hypertension, smoking, obesity); SBP 130-180 mmHg and taking ≤3 antihypertensive drugs, 24-hour protein excretion rate <1g | Body mass index ≥45 kg/$m^2$, serum creatinine ≥132.6 $\mu$mol/l and other serious illness | Jan to Jun 2001, then Jan 2003 to Oct 2005 | 4733 (48) | 2362 (48) | 2371 (48) | SBP <120 mmHg | SBP <140 mmHg | Drug classes available in clinical practice | 4.7 | 20 |
| Cardio-Sis[7 14] | Italy | Age ≥55 years, SBP ≥150 mmHg, taking antihypertensive drug ≥12 weeks, ≥1 CV risk factor (smoking, dyslipidaemia, family history of premature CVD, prior TIA or stroke, established CAD or PAD | Fasting blood glucose ≥7 mmol/l, diabetes, serious conditions, renal disease, valvular heart disease, left ventricular hypertrophy, atrial fibrillation, substance misuse. | Feb 2005 to Feb 2007 | 1111 (59) | 558 (59) | 553 (59) | SBP <130 mmHg | SBP <140 mmHg | Diuretic (Furosemide), ACEI (Ramipril), ARB (Telmisartan), CCB (Amlodipine), $\beta$ -blocker (Bisoprolol), Clonidine | 4.7 | No data |
| HOMED-BP[15] | Japan | Self-measured SBP 135-179 mmHg or DBP 85-119 mmHg, but not if DBP <65 or SBP <110 mmHg (clinic SBP <220 mmHg and DBP <125 mmHg) | None specified | May 2001 to Oct 2009 | 3518 (50) | 1759 (50) | 1759 (50) | SBP <120 and DBP <80 mmHg | BP 125-134/80-84 mmHg | ACEI, ARB or CCB; Diuretic; $\beta$-blocker; then other drug class (avoid reaching BP <110/65 mmHg) | 4.9 | 8 |
| SPRINT[16] | USA and Puerto Rico | Age ≥50y years, SBP 130-180 mmHg, increased CVD risk (clinical/subclinical CVD other than stroke, CKD excluding polycystic kidney disease and with eGFR of 20-60 ml/min/1.73$m^2$ body surface area, 10-year Framingham CVD risk ≥15%, age ≥75y) | Diabetes or prior stroke | Nov 2010 to Mar 2013 | 9361 (36) | 4678 (36) | 4683 (35) | SBP <120 mmHg | SBP <140 mmHg | All major drug classes | 3.0 | 13 |
| UKPDS[17-19] | UK | Age 25-65 years, newly-diagnosed diabetes, and hypertension (untreated: SBP ≥160 mmHg and/or DBP ≥90 mmHg; treated: SBP ≥150 mmHg and/or DBP ≥85 mmHg) | Ketonuria, recent MI, angina, heart failure, >1 major vascular episode, serum creatinine >15 $\mu$mol/, retinopathy, malignant hypertension, uncorrected endocrine abnormality, severe concurrent illness | 1987 to 1991 | 1906 | 758 (46) | 390 (42) | BP <150/85 mmHg | BP <180/105 mmHg | ACEI (Captopril) and $\beta$-blocker (Atenolol) for more intense arm; For both arms: diuretic (Furosemide), CCB (Nifedipine), methlydopa then $\alpha$-blocker (Prazosin) | 7.9 | 5 |
| VALISH[20 21] | Japan | Age ≥70 to <85 years, isolated hypertension (SBP >160 mmHg and DBP <90 mmHg) | Secondary or malignant hypertension, BP ≥200/≥90 mmHg, recent CeVD or MI, recent/planned revascularisation, heart failure, aortic stenosis, valvular heart | Feb 2004 to Aug 2005 | 3079 (62) | 1545 (62) | 1534 (63) | SBP <140 mmHg | SBP ≥140 to <150 mmHg | ARB (Valsartan) then other antihypertensive agents such as diuretics and CCB but not other ARB drugs | 2.6 | 11 |

| | |
|---|---|
| | disease, atrial fibrillation/flutter, serious arrhythmia, renal/liver dysfunction |

**Table S3. Characteristics of trials included in the BPLTTC study (cont'd).**

B. Blood pressure-lowering trials comparing treatment with placebo

| Trial | Setting | Inclusion criteria | Exclusion criteria | Recruitment period | Randomisation group, No. of participants (% of women) | | | Drug intervention | Any additional treatment | Treatment goal for blood pressure control | Follow-up duration (years) | No. of follow-up BP measures |
|---|---|---|---|---|---|---|---|---|---|---|---|---|
| | | | | | All | Inter-vention | Placebo | | | | | |
| ACTIVE I[22] | Multi-country | Atrial fibrillation, ≥1 risk factor (age ≥75 years, on antihypertensive treatment, history of stroke, TIA or non-CNS embolism, LVEF <45%, PVD, or age 55-74 years with either CAD or diabetes) | Use of anticoagulant, peptic ulcer disease in past 6 months, history of intracerebral haemorrhage, thrombocytopaenia or mitral stenosis | Jun 2003 to May 2006 | 9016 (39) | 4518 (39) | 4498 (39) | ARB (Irbesartan) | None | None specified | 4.1 | 9 |
| ADVANCE[23] | Multi-country | Age ≥55 years T2D (diagnosed aged ≥30y), ≥1 major CVD or ≥1 CVD risk factor (microvascular disease, smoking, dyslipidaemia, microalbuminuria, T2D for ≥10 years, age ≥65 years) | HbA1c target (≤6.5%), definite indication for long-term insulin therapy | Jul 2001 to Mar 2003 | 11,140 (43) | 5569 (42) | 5571 (43) | ACEI (Perindopril) and diuretics (Indapamide) as fixed dose combination drug | At physician's discretion, but not thiazide diuretics, and only perindopril as ACEI allowed | None specified | 4.2 | 12 |
| ANBP[24 25] | Australia | Age 30-69 years with mild hypertension (DBP 95-110 mmHg and SBP <200 mmHg) | Antihypertensive treatment in past 3 months, recent angina or MI, stroke, hormone therapy, asthma, diabetes, gout, serious disease, tricyclic antidepressant use | 1973 to March 1979 | 3427 (37) | 1721 (37) | 1706 (36) | Diuretic (Chlorothiazide) | Methyldopa, propranolol, or pindolol, then hydralazine or clonidine | Reduce DBP to ≤90 mmHg (after two years, further reduced to 80 mmHg) | 3.6 | 4 |
| BENEDICT[26 27] | Italy | Age ≥40 years, untreated SBP ≥130 / DBP ≥85 mmHg or needing treatment to attain below these levels, T2D for <25 years, urinary albumin excretion rate <20 $\mu$g/min, serum creatinine ≤133 $\mu$mol/l | HbA1c ≥11%, nondiabetic renal disease | Around 2000 to 2003 | 1209 (48) | 907 (47) | 302 (50) | ACEI (Trandolapril), CCB (Verapamil), both ACEI and CCB | Diuretics (HCTZ or furosemide), doxazosin, prazosin, clonidine, methyldopa or $\beta$-blocker, minoxidil, or CCB | Reduce BP to 120/80 mmHg | 3.1 | 14 |
| CAMELOT[28] | Multi-country (US, Canada, Europe) | Age 30-79 years, coronary artery stenosis >20% by angiography, DBP <100 mmHg | Left middle coronary artery obstruction >50%, LVEF <40%, heart failure | Apr 1999 to Apr 2002 | 1991 (26) | 1336 (26) | 655 (27) | CCB (Amlodipine), ACEI (Enalapril) | Allowed to continue $\beta$-blocker, $\alpha$-blocker, diuretics | None specified | 1.6 | 7 |
| DIABHYCAR[29 30] | Multi-country | Age ≥50 years, T2D, urinary albumin excretion ≥20 mg/l in two consecutive urine samples | Serum creatinine >150 $\mu$mol/l, use of insulin, ACEI or ARB, heart failure, recent MI, urinary tract infection | Feb 1995 to Apr 1998 | 4912 (30) | 2443 (30) | 2469 (30) | ACEI (Ramipril) | Usual treatment | None specified | 3.9 | 7 |
| Dutch TIA Trial[31] | The Netherlands | TIA or non-disabling ischaemic stroke (Rankin Scale ≤3) in past 3 months | Cerebral ischaemia from identifiable causes other than arterial thrombosis or embolism | Feb 1986 to Mar 1989 | 1473 (36) | 732 (34) | 741 (38) | $\beta$-blocker (Atenolol) | None specified | None specified | 2.3 | 6 |
| EUROPA[32 33] | Multi-country (Europe) | Age ≥18 years, documented MI >3 months before screening, revascularisation >6 months before screening, >70% coronary obstruction | Heart failure, hypotension, uncontrolled hypertension, renal insufficiency, serum potassium >5.5 mmol/L | Oct 1997 to Jun 2000 | 12,218 (15) | 6110 (14) | 6108 (15) | ACEI (Perindopril) | None specified | None specified | 4.2 | 9 |
| EWPHE[34 35] | Multi-country | Age ≥60 years, BP 160-239/90-119 mmHg | Curable causes of high BP, retinopathy, heart failure, stroke history, hepatitis/cirrhosis, gout, malignancy, diabetes requiring insulin treatment | From 1972 | 840 (70) | 416 (69) | 424 (71) | Diuretic (HCTZ or triamterene) | Methyldopa | Reduce BP but at unspecified levels | 4.6 | 11 |
| HOPE[36] | Multi-country | Age ≥55 years, CAD, stroke, PVD or diabetes, plus ≥1 risk factor (hypertension, dyslipidaemia, | Heart failure, left ejection fraction <40%, using ACEI or Vitamin E, | Dec 1993 to Jun 1995 | 9297 (27) | 4656 (28) | 4652 (26) | ACEI (Ramipril) | None specified | None specified | 4.5 | 3 |

| | | | | | | | | | | | | |
|---|---|---|---|---|---|---|---|---|---|---|---|---|
| | | smoking, or documented microalbuminuria) | uncontrolled hypertension, nephropathy, or recent MI or stroke | | | | | | | | | |
| HYVET[37] | Multi-country | Age ≥80y years, sustained SBP ≥160 mmHg | Accelerated or secondary hypertension, recent haemorrhagic stroke, heart failure, serum creatinine >150 $\mu$mol/L, serum potassium <3.5 or >5.5 mmol/L, gout, and dementia | From 2000 | 3845 (60) | 1933 (61) | 1912 (60) | Diuretic (Indapamide) | ACEI (Perindopril) | Reduce BP to 150/80 mmHg | 2.1 | 5 |
| IDNT[6] | USA | Age 30-70 years, T2D, hypertension (BP ≥135/85 mmHg or taking anti-hypertensive drug), proteinuria, serum creatinine ($\mu$mol/l): 88 to 265 (women) or 106 to 265 (men) | None specified | Mar 1996 to Feb 1999 | 1715 (34) | 1146 (36) | 569 (29) | ARB (Irbesartan) and CCB (Amlodipine) | Others except ACEI, ARB and CCB | SBP <135 (10 mmHg lower if baseline value >145 mm Hg); DBP <85 mmHg | 2.6 | No data |
| PART 2[38] | New Zealand | Age ≤75 years, diagnosis (in past 5 year) of MI, documented CAD, TIA or intermittent claudication | Heart failure, serious nonvascular disease, SBP >160 mmHg, DBP >100 mm Hg, DBP <100 mmHg during pre-randomization run-in period | Not specified; Publication in 2000 | 617 (18) | 308 (18) | 309 (18) | ACEI (Ramipril) | None | None specified | 4.6 | 5 |
| PEACE[39] | Multi-country (USA, Puerto Rico, Canada and Italy) | Age ≥50 years, documented CAD | Unstable angina, severe valvular heart disease, recent revascularisation, planned elective revascularisation, limited 5-year survival, serum creatinine >177 $\mu$mol/l, serum potassium >5.5 mmol/l | Nov 1996 to Jun 2000 | 8290 (18) | 4158 (19) | 4132 (17) | ACEI (Trandolapril) | None | None specified | 4.7 | 7 |
| PREVEND IT[40] | The Netherlands | Microalbuminuria, SBP <160/100 mmHg (no previous antihypertension treatment) | Creatinine clearance <60% of normal age-adjusted value | Apr 1998 to Jun 1999 | 864 (35) | 431 (34) | 433 (36) | ACEI (Fosinopril) | None | None specified | 3.8 | 16 |
| PREVENT[41 42] | USA and Canada | Age 30-80 years, documented CAD, DBP <95 mmHg, cholesterol <325 mg/dl, fasting blood glucose <200 mg/dl | Contraindication for dihydropyridines, uncontrolled hypertension, diabetes and other major illness | Nov 1992 to Sep 1994 | 825 (20) | 417 (20) | 408 (20) | CCB (Amlodipine) | None | None specified | 3.0 | 3 |
| PROFESS[43 44] | Multi-country | Age ≥55 years with ischaemic stroke <90 days before randomization (later modified to include age 50 to 54 years or had stroke 90 to 120 days before randomisation if with ≥2 additional risk factors: diabetes, hypertension, smoker, obesity previous CVD, end-organ damage or hyperlipidaemia) and remained stable[a] | Haemorrhagic stroke, severe disability after the qualifying stroke, contraindication to treatments | Sep 2003 to Jul 2006 | 19,798 (36) | 9873 (35) | 9925(36) | ARB (Telmisartan) | At physician's discretion to control blood pressure: diuretic, then $\beta$-blocker or CCB, then ACEI but not ARB. | None specified | 2.5 | 6 |
| PROGRESS[45 46] | Multi-country (Asia, Australasia and Europe) | Stroke or TIA in past 5 years | Indication or contraindication for ACEI | Jun 1995 to Nov 1997 | 6105 (30) | 3051 (30) | 3054 (30) | ACEI (Perindopril) and/or diuretic (Indapamide) | None | Intensity of lowering based on clinical intention determined before randomization | 3.9 | 4 |
| SHEP[47] | USA | Age ≥60 years, isolated systolic hypertension (BP 160-219/<90 mmHg, not on treatment) | Major CVD, cancer, alcoholic liver disease, renal dysfunction, competing risk of SHEP primary endpoint or presence of medical management exclusions | Mar 1985 to Jan 1988 | 4736 (57) | 2365 (56) | 2371 (57) | Diuretic (Chlorthalidone) and $\beta$-blocker (Atenolol) | None | If baseline SBP >180 mmHg: reduce to <160 mmHg; If baseline SBP 160-179 mmHg, reduce by 20 mmHg | 5.0 | 4 |

| | | | | | | | | | | | | |
|---|---|---|---|---|---|---|---|---|---|---|---|---|
| Syst-Eur[48] | Multi-country | Age ≥60 years, sitting SBP 160-219 mmHg, sitting DBP <95 mmHg, and standing SBP ≥140 mmHg | Secondary hypertension, retinal haemorrhage/papilloedema, heart failure, dissecting aortic aneurysm, serum creatinine ≥180 $\mu$mol/l, recent severe nosebleeds, stroke or MI, dementia, disorders prohibiting standing position, severe CVD/non-CVD | Dec 1988 to Jan 1997 | 4695 (67) | 2398 (67) | 2297 (66) | CCB (Nitrendipine) | Combined with or replaced by ACEI (Enalapril), diuretic (HCTZ), or both | Reduce sitting SBP by ≥20 mmHg to <150 mmHg | 2.6 | 3 |
| TRANSCEND[49,50] | Multi-country | Intolerant to ACEI and with established CAD, PVD, CeVD or diabetes with end-organ damage | Heart failure, valvular/cardiac outflow tract obstruction, pericarditis, congenital heart disease, unexplained syncope, recent revascularisation, SBP >160 mmHg, heart transplantation, subarachnoid haemorrhage, significant renal stenosis, renal or hepatic dysfunction | Nov 2001 to May 2004 | 5926 (43) | 2954 (43) | 2972 (43) | ARB (Telmisartan) | None | None specified | 4.9 | 9 |

**Table S3. Characteristics of trials included in the BPLTTC study (cont'd).**

#### C. Trials comparing different drug classes

| Trial | Country | Inclusion criteria | Exclusion criteria | Recruitment period | Randomisation groups | No. of participants (% women) | Additional (open-label) treatment | Treatment target for blood pressure control | Follow-up duration (y) | No. of follow-up BP measures |
|---|---|---|---|---|---|---|---|---|---|---|
| AASK[8 9] | USA | Age 18-70 years, African-American, hypertension, renal disease (GFR=20-65 ml/min per 1.73$m^2$) | DBP <95 mmHg, diabetes, urine protein:creatinine ratio >25, recent malignant or hypertension, non-blood pressure-related CKD, serious systemic disease, heart failure | Feb 1995 to Sept 1998 | All<br>ACEI (Ramipril)<br>CCB (Amlodipine)<br>$\beta$-blocker (Metoprolol) | 1094 (39)<br>436 (39)<br>217 (39)<br>441 (39) | Furosemide, doxazosin, clonidine, hydralazine and minoxidil (sequentially) | None specified | 4.8 | 54 |
| ABCD[10-12] | USA | Age 0-74 years, with T2D, DBP ≥80 mmHg, not on antihypertensive treatment | Recent CAD or CeVD, heart failure, renal disease | Mar 1991 to May 1993 | All<br>CCB (Nisoldipine)<br>ACEI (Enalapril) | 470 (33)<br>235 (32)<br>235 (33) | $\beta$-blocker (Metoprolol), diuretic (HCTZ), or others but not CCB or ACEI | None specified | 4.7 | 8 |
| ALLHAT[51 52] | Multi-country | Age ≥55y years stage 1 or 2 hypertension plus ≥1 risk factor (MI or stroke >6 months previously, left ventricular hypertrophy, T2D, smoking, HDL <0.91 mmol/l), other atherosclerotic CVD | Symptomatic or hospitalisation for heart failure, LVEF <35% | Feb 1994 to Jan 1998 | All<br>Diuretic (Chlorthalidone)<br>CCB (Amlodipine)<br>ACEI (Lisinopril)<br>$\alpha$-blocker (Doxazosin) | 42,418 (47)<br>15,255 (47)<br>9048 (47)<br>9054 (46)<br>9061 (46) | Atenolol, clonidine or reserpine | BP <140/90 mmHg | 4.8 | 3 |
| ANBP2[53] | Australia | Age 65-84 years, SBP ≥160 mmHg or DBP ≥90 mmHg (if SBP≥140 mmHg), no recent CVD | Serious illness, plasma creatinine >221 $\mu$mol/l, malignant hypertension, dementia | April 1995 to Jun 1998 | All<br>ACEI (Enalapril)<br>Diuretic (HCTZ) | 6083 (51)<br>3044 (50)<br>3039 (52) | $\beta$-blocker, CCB and $\alpha$-blocker | Lower SBP by 20 mmHg to <160 mmHg (<140 mmHg if tolerated,) and by 10 mmHg DBP to <90 mmHg (<80 mmHg if tolerated) | 4.1 | 9 |
| ASCOT-BPLA[54 55] | Multi-country | Age 40-79 years, untreated (SBP ≥160 or DBP ≥100 mmHg) or treated hypertension (SBP ≥140 or DBP ≥90 mmHg), ≥3 CVD risk factors (documented LVH, abnormal ECG, T2D, PAD, previous stroke or TIA, male sex, age ≥55 years, microalbuminuria or proteinuria, smoking, TC:HDL ≥6, family history of premature coronary heart disease | Previous MI, current treatment for angina, recent CeVD, fasting triglycerides >4.5 mmol/l, heart failure, arrhythmia, haematological or biochemical abnormality at screening | Feb 1998 to May 2000 | All<br>CCB (Amlodipine-based)<br>$\beta$-blocker (Atenolol-based) | 19,257 (23)<br>9639 (23)<br>9618 (23) | For CCB arm: plus ACEI (Perindopril); For $\beta$-blocker arm: plus diuretic (bendroflumethiazide) and potassium | With diabetes: BP <140/90 mmHg; Without diabetes: BP <130/80 mmHg | 5.3 | 12 |
| BENEDICT[26 27] | Italy | Age ≥40 years, untreated SBP ≥130 or DBP ≥85 mmHg or needing treatment to attain below these levels, T2D for <25 years, urinary albumin excretion rate <0 $\mu$g/min on two consecutive measures, serum creatinine ≤133 $\mu$mol/l | HbA1c ≥11%, nondiabetic renal disease | Around 2000 to 2003 | All (excluding placebo)<br>ACEI (Trandolapril)<br>CCB (Verapamil)<br>ACEI (Trandolapril) and CCB (Verapamil) | 907 (47)<br>302 (48)<br>303 (46)<br>302 (45) | Diuretic (HCTZ or furosemide), then doxazosin, prazosin, clonidine, methyldopa or $\beta$-blocker, then minoxidil, or CCB | BP 120/80 mmHg | 3.1 | 14 |
| CAMELOT[28] | Multi-country | Age 30-79 years, coronary artery stenosis >20% by angiography and DBP <100 mmHg | Left middle coronary artery obstruction >50%, LVEF <40%, heart failure | Apr 1999 to Apr 2002 | All (excluding placebo)<br>CCB (Amlodipine)<br>ACEI (Enalapril) | 1336 (26)<br>663 (24)<br>673 (28) | Allowed to continue $\beta$-blocker, $\alpha$-blocker, diuretic | None specified | 1.6 | 7 |
| CAPPP[56 57] | Sweden and Finland | Age 25-66 years, DBP ≥100 mmHg on two occasions | Secondary hypertension, serum creatinine >150 $\mu$mol/, condition requiring $\beta$-blocker treatment | Dec 1989 to Apr 1995 | All<br>ACEI (Captopril)<br>Conventional: $\beta$-blocker (Atenolol or metoprolol) and/or diuretic (HCTZ, bendofluazide) | 10,985 (47)<br>5492 (45)<br>5493 (48) | Diuretic and CCB if necessary | Supine DBP <90 mmHg | 5.8 | 6 |
| CASE-J[58 59] | Japan | Age 20-85 years, ≥1 high-risk factor: SBP ≥180 or DBP ≥110 mmHg, T2D, history of angina pectoris, MI, stroke, TIA >6 months | BP ≥200/120 mmHg, T1D, heart failure, ejection fraction <40%, atrial fibrillation, cancer | Sep 2001 to Jan 2003 | All<br>ARB (Candesartan)<br>CCB (Amlodipine) | 4703 (45)<br>2354 (46)<br>2349 (43) | Allowed to continue background treatment (diuretic, $\alpha$-blocker, $\beta$- | BP (mmHg) by age (years): <60: <130/85; 60-69: <140/90; 70- | 3.1 | 6 |

| | | | | | | | | | | |
|---|---|---|---|---|---|---|---|---|---|---|
| | | prior to screening, LVH, proteinuria or serum creatinine ≥1.3 mg/100 ml, peripheral artery obstruction | | | | | blocker); Can add other treatment except ARB, CCB, ACEI | 79: <150/90; ≥80: <160/90 | | |
| COLM[60 61] | Japan | Age 65-84 years, hypertension (treated: BP ≥140/90 mmHg; untreated: BP ≥160/100 mmHg), CVD history or CVD risk factors (diabetes, dyslipidaemia) | Secondary/malignant hypertension, recent major CVD, revascularisation, angina pectoris hospitalisation or severe heart failure, atrial fibrillation, hepatic or renal dysfunction | Apr 2007 to Sep 2008 | All | 5141 (48) | $\beta$-blocker, $\alpha$-blocker, ACEI | BP <140/90 mmHg | 3.0 | 8 |
| | | | | | ARB (Olmesartan) and CCB (Amlodipine or azelnidipine) | 2568 (48) | | | | |
| | | | | | ARB (Olmesartan) and diuretic (HCTZ, Trichlormethiazide, or indapamide) | 2573 (48) | | | | |
| CONVINCE[62 63] | Multi-country | Age ≥55 years, hypertension, ≥1 CVD risk factor (e.g., diabetes, smoking) | Heart failure, dysrhythmia, secondary hypertension, recent MI or stroke, renal disease, other serious disease, BP ≥190/110 mmHg without treatment | Sep 1996 to Dec 1998 | All | 16476 (55) | Additional treatment if necessary | BP <140/90 mmHg | 2.8 | 4 |
| | | | | | CCB (Verapamil) | 8179 (56) | | | | |
| | | | | | $\beta$-blocker (Atenolol) or diuretic (HCTZ) | 8297 (56) | | | | |
| COPE[64] | Japan | Age 40-85 years, BP ≥140/90 mmHg | SBP ≥200 or DBP ≥120 mmHg, secondary hypertension, diabetes, recent CVD or revascularisation, heart failure, atrial fibrillation/flutter, hepatic or renal dysfunction, congenital or rheumatic heart disease, cancer | Jun 2003 to Nov 2006 | All | 3293 (49) | Additional treatment if necessary | BP <140/90 mmHg | 3.6 | 5 |
| | | | | | CCB/ARB (Benidipine/ARB) | 1110 (49) | | | | |
| | | | | | CCB/$\beta$-blocker (Benidipine/$\beta$-blocker) | 1089 (49) | | | | |
| | | | | | CCB/Diuretic (Benidipine/Thiazide) | 1094 (49) | | | | |
| E-COST[65] | Japan | Age 35-79 years, BP 140-180/90-110 mmHg | Diabetes, dysglycemia, secondary hypertension, recent MI or stroke, angina pectoris requiring $\beta$-blocker treatment, heart failure, left ventricular ejection fraction <40% | Sept to Dec 1999 | All | 2048 (52) | Additional treatment but not ARB or ACEI | BP <140/90 mmHg | 3.1 | 2 |
| | | | | | ARB (Candesartan) | 1053 (56) | | | | |
| | | | | | Conventional (mainly CCB and $\beta/\alpha$-blocker) | 995 (48) | | | | |
| ELSA[66 67] | Multi-country | Age 45-79 years, BP 150-210/95-115 mmHg | Recent MI or stroke, and T2D | Possibly between 1994 to 1998 | All | 2334 (45) | Diuretic (HCTZ) | DBP <95 mmHg | 3.4 | 10 |
| | | | | | CCB (Lacidipine) | 1177 (46) | | | | |
| | | | | | $\beta$-blocker (Atenolol) | 1157 (45) | | | | |
| HIJ-CREATE[68] | Japan | Age 20-80 years, CAD hospitalisation and hypertension (BP ≥140/90 mmHg or antihypertensive treatment use) | Secondary hypertension, recent AMI or CeVD, severe aortic valve stenosis, cardiomyopathy, serum creatinine >2 mg/dl, serum potassium >5 mmol/l, hepatic dysfunction, malignancy | Jun 2001 to Apr 2004 | All | 2049 (20) | | BP <130/85 mmHg | 4.0 | 5 |
| | | | | | ARB (Candesartan) | 1024 (18) | | | | |
| | | | | | Non-ARB (including ACEI) | 1025 (21) | | | | |
| HOMED-BP[15] | Japan | Self-measured SBP 135-179 mmHg or DBP 85-119 mmHg, but not if DBP <65 mmHg or SBP <110 mmHg (clinic SBP <220 mmHg and DBP <125 mmHg) | None specified | May 2001 to Oct 2009 | All | 3518 (50) | Diuretic; $\beta$-blocker; then other drugs (avoid reaching BP <110/65 mmHg) | Either BP <120/80 mmHg or BP 125-134/80-84 mmHg | 4.9 | 8 |
| | | | | | ACEI | 1172 (50) | | | | |
| | | | | | ARB | 1175 (50) | | | | |
| | | | | | CCB | 1171 (50) | | | | |
| IDNT[6] | USA | Age 30-70 years, T2D, hypertension (BP ≥135/85 mmHg or taking antihypertensive drug), proteinuria, serum creatinine of 88 to 265 $\mu$mol/l (women) or 106 to 265 $\mu$mol/l (men) | None specified | Mar 1996 to Feb 1999 | All (except placebo) | 1146 (36) | Others except ACEI, ARB and CCB | SBP <135 mmHg (10 mmHg lower if baseline value >145 mmHg); and DBP <85 mmHg | 2.6 | No data |
| | | | | | ARB (Irbesartan) | 579 (35) | | | | |
| | | | | | CCB (Amlodipine) | 567 (37) | | | | |
| INSIGHT[69] | Multi-country | Age 55-80 years, hypertensive (SBP ≥150 or DBP ≥95 mmHg, or SBP ≥160 mmHg), ≥1 other risk factor (TC ≥6.43 mmol/l, smoking, family history of premature MI, CAD, other CVD | None specified | Sep 1994 to Jun 1996 | All | 6321 (54) | $\beta$-blocker or ACEI, then others except CCB or diuretic | SBP/DBP reduction by 20/10 mmHg or SBP/DBP <140/90 mmHg | 2.8 | 15 |
| | | | | | CCB (Nifedipine) | 3157 (54) | | | | |
| | | | | | Diuretic (Co-amilozide [HCTZ + amiloride]) | 3164 (53) | | | | |
| INVEST[70] | Multi-country | Age ≥50 years, documented CAD, essential hypertension requiring drug therapy, heart failure Class I-III[b] | Patients taking $\beta$-blocker within two weeks of randomization or for recent MI | From Jan 1998 | All | 21,230 (52) | ACEI (Trandolapril) and/or diuretic (HCTZ) | SBP/DBP <140/90 mmHg; <130/85 mmHg with diabetes or renal impairment | 2.8 | 5 |
| | | | | | CCB (Verapamil) | 10,648 (52) | | | | |
| | | | | | Non-CCB (Atenolol) | 10,672 (52) | | | | |
| JMIC-B[71] | Japan | | | | All | 1650 (31) | | BP <150/90 mmHg | 2.3 | 5 |

| | | | | | | | | | | |
|---|---|---|---|---|---|---|---|---|---|---|
| | | Age <75 years, hypertension (BP ≥160/≥95 mmHg or both SBP ≥150 and DBP ≥90 mmHg, or antihypertensive treatment), CAD or meeting both criteria: history of >2 anginal attacks per week with stable frequency and ST-segment depression of ≥1 mm on stress test (or detection of MI with myocardial scintigraphy) | MI, unstable angina, DBP ≥120 mmHg, secondary hypertension, symptomatic CeVD, heart failure, atrial fibrillation/arrhythmias, renal or hepatic dysfunction, uncontrollable diabetes and familial hypercholesterolaemia | Jan 1994 to Jul 1997 | CCB (Nifedipine)<br>ACEI (Enalapril, imidapril or lisinopril) | 828 (32)<br>822 (30) | $\alpha$-blocker (doxazosin, bunazosin or prazosin); nitrates or $\beta$-blocker for angina if needed | | | |
| LIFE[72 73] | Multi-country | Age 55-80 years, hypertension (SBP 160-200 mmHg; DBP 95-115 mmHg), electrocardiogram signs of LVH | Secondary hypertension, recent MI or stroke, angina pectoris requiring treatment, heart failure or left ejection fraction ≤40% | June 1995 to May 1997 | All<br>ARB (Losartan)<br>$\beta$-blocker (Atenolol) | 9193 (54)<br>4605 (54)<br>4588 (54) | Diuretic (HCTZ) and other antihypertensive treatment except ACEI, ARB and $\beta$-blocker | BP 140/90 mmHg | 4.9 | 15 |
| MOSES[74] | Germany and Austria | Hypertension requiring treatment, documented TIA, ischaemic stroke or cerebral haemorrhage | Internal carotid artery occlusion or stenosis >70%, heart failure, age >85 years, on anticoagulant for cardiac arrhythmia, high-grade aortic or mitral valve stenosis, unstable angina | Oct 1998 to Feb 2002 | All<br>ARB (Eprosartan)<br>CCB (Nitrendipine) | 1352 (46)<br>681 (46)<br>671 (45) | Diuretic, $\beta$-blocker, $\alpha$-blocker or centrally-acting drugs; ACEI, ARB or CCB only if clinically necessary | BP <140/90 mmHg | 3.3 | 7 |
| NICS-EH[75] | Japan | Age ≥60 years, SBP 160-220 mmHg and DBP <115 mmHg and no cardiovascular complications | None specified | Oct 1989 to Apr 1992 | All<br>CCB (Nicardipine)<br>Diuretic (Trichlormethiazide) | 414 (67)<br>204 (60)<br>210 (74) | Titration but no additional treatment | BP response sufficient as determined by the investigator | 3.2 | 3 |
| NORDIL[76] | Norway and Sweden | Age 50-74 years, untreated hypertension (DBP ≥100 mmHg on two occasions); if previously treated, DBP ≥100 mmHg on two consecutive visits at one week apart during run-in period and no treatment was given | Age <50 or ≥70y, bradycardia, secondary hypertension, atrial fibrillation, recent CeVD or MI, heart failure | Oct 1992 to Oct 1999 | All<br>CCB (Diltiazem)<br>$\beta$-blocker and/or diuretic (Thiazide) | 10,881 (51)<br>5410 (51)<br>5471 (51) | CCB group: plus ACEI, diuretic, $\alpha$-blocker or any other if necessary; $\beta$-blocker group: ACEI or $\alpha$-blocker or any other drug except CCB if necessary | | 4.2 | 8 |
| ONTARGET[49 77] | Multi-country | CAD, PAD, CeVD or diabetes with end-organ damage | Heart failure, pericarditis, congenital heart disease, unexplained syncope , planned revascularisation <3 months of consent, uncontrolled hypertension, heart transplant, subarachnoid haemorrhage, renal artery disease, proteinuria, hepatic dysfunction, volume or sodium depletion, primary hyper-aldosteronism, hereditary fructose intolerance, other serious conditions | Jan 2002 to Aug 2003 | All<br>ACEI (Ramipril)<br>ARB (Telmisartan)<br>ACEI (Ramipril) and ARB (Telmisartan) | 25,620 (27)<br>8576 (27)<br>8542 (26)<br>8502 (26) | None | None specified | 4.8 | 9 |
| STOP Hyper-tension-2[78] | Sweden | Aged 70-84 years, SBP ≥180 mmHg and/or DBP ≥105 mmHg | Not specified | Sep 1992 to Dec 1994 1987 to 1991 | All<br>Conventional: $\beta$-blocker (Atenolol or metoprolol), diuretic (HCTZ) or both<br>ACEI (Enalapril or lisinopril<br>CCB (Felodipine or isradipine) | 6614 (67)<br>2213 (68)<br>2205 (66)<br>2196 (66) | | BP <160/95 mmHg | 4.5 | 10 |
| UKPDS[17-19] | UK | Age 25-65 years, newly-diagnosed diabetes, and hypertension (untreated: SBP ≥160 mmHg and/or DBP ≥90 mmHg; treated: SBP ≥150 mmHg and/or DBP ≥85 mmHg) | Ketonuria, recent MI, angina, heart failure, >1 major vascular episode, serum creatinine >175 umol/L, retinopathy, malignant hypertension, uncorrected endocrine abnormality, severe concurrent illness | | All<br>ACEI (Captopril)<br>$\beta$-blocker (Atenolol) | 758 (46 )<br>400 (49)<br>358 (43) | Sequentially: Diuretic (furosemide), CCB (nifedipine), methyldopa, ARB (prazosin) | BP <150/85 mmHg | 7.9 | 5 |
| VALUE[79 80] | Multi-country | Age ≥50 years, hypertension, CVD, CVD risk factors (male sex, age >50 years, diabetes, current smoking, high | Renal artery stenosis, recent CAD or CeVD, severe hepatic disease or chronic renal failure, heart failure, on | Sept 1997 to Dec 1999 | All<br>ARB (Valsartan-based)<br>CCB (Amlodipine-based) | 15,245 (42)<br>7649 (42)<br>7596 (42) | Diuretic (HCTZ, then other antihypertensive drugs except ARB (ACEI | BP <140/90 mmHg | 4.2 | 12 |

| | | | | | | | | | | |
|---|---|---|---|---|---|---|---|---|---|---|
| | | cholesterol, LVH, proteinuria, serum creatinine 150 to 265 $\mu$mol/l) | monotherapy with $\beta$-blocker for CAD and hypertension | | | | or CCB if clinically indicated other than for hypertension) | | | |
| VHAS[81] | Italy | Age 40-65 years, BP ≥160/95 mmHg | Secondary hypertension, recent stroke or TIA, CAD, PAD, bradycardia, arrhythmias, heart failure, renal or hepatic dysfunction, hyperuricaemia, hypokalemia, T1D, familial dyslipidemia, serious concomitant disease | Probably early 1990s (publication in 1997) | All | 1414 (51) | ACEI (Captopril ) | Sitting DBP ≤90 mmHg; ≤95 mmHg if lowered by ≥10% from baseline values | 1.7 | 2 |
| | | | | | CCB (Verapamil) | 707 (50) | | | | |
| | | | | | Diuretic (Chlorthalidone) | 707 (52) | | | | |

Trial name acronyms are described in full in the Trial acronym legend in the Supplement; Data on participant numbers and proportions, mean duration of follow-up and mean number of follow-up blood pressure measurements are from the datasets provided to the collaboration; BP – blood pressure; GFR – glomerular filtration rate; DBP – diastolic blood pressure; CKD – chronic kidney disease; T2D – type 2 diabetes; CAD – coronary artery disease; CeVD – cerebrovascular disease; CVD – cardiovascular disease; HbA1c - glycated haemoglobin; SBP – systolic blood pressure; TIA – transient ischaemic attack; PAD – peripheral artery disease; MI – myocardial infarction; CNS – central nervous system; LVEF – low ventricular ejection fraction; HDL – high-density lipoprotein; ECG – electrocardiogram; TC – total cholesterol; LVH – left ventricular hypertrophy; T1D – type 1 diabetes; CCB – calcium channel-blocker; ACEI – angiotensin-converting enzyme inhibitor; ARB – angiotensin II receptor blocker; HCTZ – hydrochlorothiazide; [a]Excludes 534 participants with heart failure at baseline; [b]Excludes 1346 participants with heart failure at baseline.

**Table S4. Risk of bias assessment of each trial.**

| Trial | Risk of bias arising from randomisation | Risk of bias due to effect of assignment to intervention | Risk of bias due to missing outcome data | Risk of bias due to measurement of outcome | Risk of bias due to reporting of result | Overall risk of bias |
|---|---|---|---|---|---|---|
| AASK | Low | Low | Low | Low | Low | Low |
| ABCD | Low | Low | Low | Low | Low | Low |
| ACCORD | Low | Some | Low | Low | Low | Low |
| ACTIVE I | Low | Low | Low | Low | Low | Low |
| ADVANCE | Low | Low | Low | Low | Low | Low |
| ALLHAT | Low | Low | Low | Low | Low | Low |
| ANBP | Low | Low | Low | Low | Low | Low |
| ANBP2 | Low | Some | Low | Low | Low | Low |
| ASCOT-BPLA | Low | Some | Low | Low | Low | Low |
| BENEDICT | Low | Low | Low | Low | Low | Low |
| CAMELOT | Low | Low | Low | Low | Low | Low |
| CAPPP | Low | Some | Low | Low | Low | Low |
| CARDIO-SIS | Low | Some | Some | Low | Low | Some |
| CASE-J | Low | Some | Low | Low | Low | Low |
| COLM | Low | Some | Low | Low | Low | Low |
| CONVINCE | Low | Low | Low | Low | Low | Low |
| COPE | Low | Some | Low | Low | Low | Low |
| DIABHYCAR | Low | Low | Low | Low | Low | Low |
| Dutch TIA Trial | Low | Low | Low | Low | Low | Low |
| E-COST | High | Some | Low | Low | Low | High |
| ELSA | Low | Low | Low | Low | Low | Low |
| EUROPA | Low | Low | Low | Low | Low | Low |
| EWPHE | Low | Low | Low | Low | Low | Low |
| HIJ-CREATE | Low | Some | Low | Low | Low | Low |
| HOMED-BP | Low | Some | Low | Low | Low | Low |
| HOPE | Low | Some | Low | Low | Low | Low |
| HYVET | Low | Low | Low | Low | Low | Low |
| IDNT | Low | Low | Some | Low | Low | Some |
| INSIGHT | Low | Low | Low | Low | Low | Low |
| INVEST | Low | Some | Low | Low | Low | Low |
| JMIC-B | Low | Some | Low | Low | Low | Low |
| LIFE | Low | Low | Low | Low | Low | Low |
| MOSES | Low | Some | Low | Low | Low | Low |
| NICS-EH | Low | Some | Low | Low | Low | Some |
| NORDIL | Low | Some | Low | Low | Low | Low |
| ONTARGET | Low | Some | Low | Low | Low | Low |
| PART 2 | Low | Low | Low | Low | Low | Low |
| PEACE | Low | Low | Low | Low | Low | Low |
| PREVEND IT | Low | Low | Low | Low | Low | Low |
| PREVENT | Low | Low | Low | Low | Low | Low |
| PROFESS | Low | Low | Low | Low | Low | Low |
| PROGRESS | Low | Low | Low | Low | Low | Low |
| SHEP | Low | Low | Low | Low | Low | Low |
| SPRINT | Low | Some | Low | Low | Low | Low |
| STOP HYPERTENSION-2 | Low | Some | Low | Low | Low | Low |
| SYST-EUR | Low | Some | Low | Low | Low | Low |
| TRANSCEND | Low | Low | Low | Low | Low | Low |
| UKPDS | Low | Some | Low | Low | Low | Low |
| VALISH | Low | Low | Low | Low | Low | Low |
| VALUE | Low | Low | Low | Low | Low | Low |
| VHAS | Low | Low | Low | Low | Low | Low |

Trial name acronyms are described in full in the Trial acronym legend in the Supplement.

**Table S5. Indicators of numbers of blood pressure-lowering drug classes given and adherence to assigned treatment, separately by trial design.**

A. Blood pressure-lowering intensity trials

| Trial | Comparison groups | Number of drugs given | | Adherence to randomized treatment assignment during follow-up | |
|---|---|---|---|---|---|
| | | No. | Description | % | Description |
| **AASK** | Less intense | 3 | Mean number of drugs given | 81.7 | On randomised treatment |
| | More intense | 2.4 | Mean number of drugs given | 80.1 | On randomised treatment |
| **ABCD**[†] | Less intense | - | Not reported | - | Mean achieved SBP during last 4 years of follow-up: 137 mmHg |
| | More intense | - | Not reported | - | Mean achieved SBP during last 4 years of follow-up: 128 mmHg |
| **ACCORD** | Less intense | 2.1 | Mean number of drugs given after year 1 follow-up | - | Mean achieved SBP after year 1 follow-up: 134 mmHg |
| | More intense | 3.4 | Mean number of drugs given after year 1 follow-up | - | Mean achieved SBP after year 1 follow-up: 119 mmHg |
| **CARDIO-SIS** | Less intense | 2.9 | Mean number of drugs given at year 2 follow-up | - | Mean achieved SBP at end of study: 136 mmHg |
| | More intense | 2.9 | Mean number of drugs given at year 2 follow-up | - | Mean achieved SBP at end of study: 132 mmHg |
| **HOMED-BP** | Less intense | 1.74 | Mean defined daily dose at last follow-up | 68.3 | Meeting target SBP (125 to 134 mmHg) at end of study |
| | More intense | 1.82 | Mean defined daily dose at last follow-up | 42.6 | % meeting target SBP (<125 mmHg) at end of study |
| **SPRINT** | Less intense | 1.8 | Mean number of drugs given | - | Mean achieved SBP at end of study: 135 mmHg |
| | More intense | 2.7 | Mean number of drugs given | - | Mean achieved SBP at end of study: 122 mmHg |
| **UKPDS** | Less intense | 11% | Taking ≥3 drug classes at year 9 follow-up | 43.0 | % of total person-years not receiving treatment |
| | More intense | 29% | Taking ≥3 drug classes at year 9 follow-up | 77.0 | % of total person-years on assigned treatment |
| **VALISH** | Less intense | 1.6 | Mean number of drugs given at end of follow-up | - | Mean achieved SBP at end of study: 142 mmHg |
| | More intense | 1.6 | Mean number of drugs given at end of follow-up | - | Mean achieved SBP at end of study: 137 mmHg |

**Table S5. Indicators of numbers of blood pressure-lowering drug classes given and adherence to assigned treatment, separately by trial design (cont'd).**

B. Placebo-controlled trials

| Trial | Comparison groups | Number of drugs given | | Adherence to randomized treatment assignment during follow-up | |
|---|---|---|---|---|---|
| | | No. | Description | % | Description |
| **ACTIVE I** | Placebo | 2.15 | Mean number of non-study drugs at year 2 | 69.7 | Did not discontinue assigned treatment |
| | ARB | 2 | Mean number of non-study drugs at year 2 | 69.7 | Did not discontinue assigned treatment |
| **ADVANCE** | Placebo | 1.57 | Mean number of non-study drugs by end of follow-up | 74.0 | On randomized treatment by end of follow-up |
| | ACEI and diuretic | 1.28 | Mean number of non-study drugs by end of follow-up | 73.0 | On randomized treatment by end of follow-up |
| **ANBP** | Placebo | 1.52 | Mean number of drugs given (calculated from numbers taking 1, 2 or ≥2 corresponding placebo) | 63.3 | On randomized treatment by end of study |
| | Diuretic | 1.82 | Mean number of drugs given (calculated from numbers taking 1, 2 or ≥2 drugs) | 66.1 | On randomized treatment by end of study |
| **BENEDICT** | Placebo | 2.18 | Mean number of drugs given | - | Not reported |
| | ACEI | 1.99 | Mean number of drugs given | - | Not reported |
| | CCB | 2.03 | Mean number of drugs given | - | Not reported |
| | ACEI and CCB | 1.87 | Mean number of drugs given | - | Not reported |
| **CAMELOT** | Placebo | - | NA | 68.9 | Did not discontinue assigned treatment |
| | CCB | 8.6 | Mean dose received (g) | 70.7 | Did not discontinue assigned treatment |
| | ACEI | 17.4 | Mean dose received (g) | 64.9 | Did not discontinue assigned drug |
| **DIABHYCAR** | Placebo | 0.22 | Mean number of non-study drugs given | 59.0 | On randomized treatment by end of study |
| | ACEI | 0.2 | Mean number of non-study drugs given | 59.3 | On randomized treatment by end of study |
| **DUTCH-TIA** | Placebo | - | Not reported | 67.7 | On randomized treatment at year 3 follow-up |
| | ß-blocker | - | Not reported | 64.2 | On randomized treatment at year 3 follow-up |
| **EUROPA** | Placebo | - | NA | 84.0 | On randomized treatment at year 3 follow-up |
| | ACEI | 7.05 | Maximum mean daily dose (mg) | 81.0 | On randomized treatment at year 3 follow-up |
| **EWPHE** | Placebo | 0.63 | Mean number of non-study drugs given | 63.0 | Did not discontinue assigned treatment |
| | Diuretic | 0.35 | Mean number of non-study drugs given | 64.2 | Did not discontinue assigned treatment |
| **HOPE** | Placebo | 0.18 | Mean number of non-study drugs given | 87.7 | On randomized treatment at final visit (12.3 % received intervention drug) |
| | ACEI | 0.14 | Mean number of non-study drugs given | 78.8 | On randomized treatment at final visit |
| **HYVET** | Placebo | - | NA | - | Not reported |
| | Diuretic | 0.73 | Mean number of non-study drug given at year 2 | - | Not reported |
| **IDNT** | Placebo | 3.3 | Mean number of non-study drugs given | - | Not reported |
| | ARB and CCB | 3 | Mean number of non-study drugs given | - | Not reported |

| | | | | | |
|---|---|---|---|---|---|
| **PART 2** | Placebo | 7 | % given non-study drug at year 4 follow-up | 75.0 | On randomized treatment at year 4 follow-up |
| | ACEI | 2.9 | % given non-study drug at year 4 follow-up | 72.0 | On randomized treatment at year 4 follow-up |
| **PEACE** | Placebo | 8.3 | % taking non-study drug at year 3 | 77.7 | On randomized treatment at year 3 follow-up |
| | ACEI | 74.5 | % taking study and non-study drug (same class) at year 3 | 68.6 | On randomized treatment at year 3 follow-up |
| **PREVEND IT** | Placebo | - | Not reported | 63.3 | Compliance above 75% at year 4 follow-up |
| | ACEI | - | Not reported | 70.7 | Compliance above 75% at year 4 follow-up |
| **PREVENT** | Placebo | 0.67 | Mean number of non-study drugs given | 83.0 | Pill count compliance |
| | CCB | 0.56 | Mean number of non-study drugs given | 79.0 | Pill count compliance |
| **PROFESS** | Placebo | 2.5 | % on non-study ARB at penultimate visit (≥30% on non-study BP-lowering drugs by end of study) | 70.8 | % on randomized treatment at year 3 follow-up |
| | Telmisartan | 2.3 | % on non-study ARB at final visit (≥25% on non-study BP-lowering drugs by end of study) | 68.3 | % on randomized treatment at year 3 follow-up |
| **PROGRESS** | Placebo | - | Not reported | 87.0 | On randomized treatment by end of follow-up |
| | ACEI and/or diuretic | - | Not reported | 86.0 | On randomized treatment by end of follow-up |
| **SHEP** | Placebo | 44.4 | % given study drug at year 5 follow-up | 55.6 | On randomized treatment at year 5 follow-up |
| | ß-blocker and diuretic | - | Over two-thirds given step 1 and/or step 2 drug at year 5 follow-up | 89.7 | On randomized treatment at year 3 follow-up |
| **SYST-EUR** | Placebo | - | Some given treatment but could not be estimated | 47.7 | On randomized treatment at year 4 follow-up |
| | CCB | 1.35 | Mean number of drugs given | 60.4 | On randomized treatment at year 4 follow-up |
| **TRANSCEND** | Placebo | 1.6 | Mean number of non-study drugs given by end of study | 40.7 | On randomized treatment by end of study (more may have received non-study drugs) |
| | ARB | 1.4 | Mean number of non-study drugs given by end of study | 80.8 | On randomized treatment by end of study |

**Table S5. Indicators of numbers of blood pressure-lowering drug classes given and adherence to assigned treatment, separately by trial design (cont'd).**

C. Drug class comparison trials

| Trial | Comparison groups | Number of drugs given | | Adherence to randomized treatment assignment during follow-up | |
|---|---|---|---|---|---|
| | | No. | Description | % | Description |
| **AASK** | ACEI | 2.7 | Mean number of drugs given | 76.8 | On randomized treatment during follow-up |
| | CCB | 2.7 | Mean number of drugs given | 83.4 | On randomized treatment during follow-up |
| | $\beta$-blocker | 2.8 | Mean number of drugs given | 83.6 | On randomized treatment during follow-up |
| **ABCD**‡ | CCB | 2.8 | Mean number of drugs given | 45.1 | Did not discontinue assigned treatment |
| | ACEI | 2.9 | Mean number of drugs given | 39.6 | Did not discontinue assigned treatment |
| **ALLHAT** | Diuretic | 1.8 | Mean number of drugs given | 71.2 | On randomized treatment at year 5 follow-up |
| | CCB | 1.9 | Mean number of drugs given | 72.1 | On randomized treatment at year 5 follow-up |
| | ACEI | 2 | Mean number of drugs given | 61.2 | On randomized treatment at year 5 follow-up |
| | $\alpha$-blocker[82] | 1.2 | Mean number of drugs given at year 4 follow-up | 75.0 | On randomized treatment at year 4 follow-up |
| **ANBP2** | ACEI | 6.0% | Given ≥3 drugs | 58.0 | On randomized treatment by end of study |
| | Diuretic | 5.0% | Given ≥3 drugs | 62.0 | On randomized treatment by end of study |
| **ASCOT-BPLA** | CCB-based | 2.2 | Mean number of drugs given | 50.0 | On randomized treatment (with and without other antihypertensive drugs) |
| | $\beta$-blocker-based | 2.3 | Mean number of drugs given | 55.0 | On randomized treatment (with and without other antihypertensive drugs) |
| **BENEDICT** | Placebo | 2.18 | Mean number of drugs given | - | Not reported |
| | ACEI | 1.99 | Mean number of drugs given | - | Not reported |
| | CCB | 2.03 | Mean number of drugs given | - | Not reported |
| | ACEI and CCB | 1.87 | Mean number of drugs given | - | Not reported |
| **CAMELOT** | Placebo | - | NA | 69.3 | Did not discontinue assigned drug |
| | CCB | 8.6 | Mean dose received (g) | 70.7 | Did not discontinue assigned drug |
| | ACEI | 17.4 | Mean dose received (g) | 64.9 | Did not discontinue assigned drug |
| **CAPPP** | $\beta$-blocker and/or diuretic | - | Not reported | - | Not reported |
| | ACEI | - | No reported | - | Not reported |
| **CASE-J** | ARB | 1.5 | Mean number of drugs given | 96.5 | Receiving >80% of assigned drug during follow-up |
| | CCB | 1.4 | Mean number of drugs given | 96.0 | Receiving >80% of assigned drug during follow-up |
| **COLM** | ARB and CCB | 2.1 | Mean number of drugs given | - | Not reported |
| | ARB and diuretic | 2.1 | Mean number of drugs given | - | Not reported |
| **CONVINCE** | CCB | - | Proportion given >1 drug increased over time | 60.6 | Did not discontinue assigned drug |

| | | | | |
|---|---|---|---|---|
| | $\beta$-blocker or diuretic | - | Proportion given >1 drug increased over time | 60.3 | Did not discontinue assigned drug |
| **COPE** | CCB and ARB | 1.22 | Mean number of drugs given | 78.6 | Did not discontinue assigned drug |
| | CCB and $\beta$-blocker | 1.26 | Mean number of drugs given | 74.1 | Did not discontinue assigned drug |
| | CCB and diuretic | 1.30 | Mean number of drugs given | 76.2 | Did not discontinue assigned drug |
| **E-COST** | Conventional | 71.5% | Taking ≥3 drugs | 81.9 | On randomized treatment by end of study |
| | ARB | 21.5% | Taking ≥3 drugs | 77.3 | On randomized treatment by end of study |
| **ELSA** | CCB | 31.8% | Taking additional drug (diuretic) | - | Not reported |
| | $\beta$-blocker | 35.9% | Taking additional drug (diuretic) | - | Not reported |
| **HIJ-CREATE** | non-ARB | 2.8 | Mean number of drugs given | 77.0 | On randomized treatment (prescribed ARB at the end of study) |
| | ARB | 2 | Mean number of drugs given | 97.5 | On randomised treatment (prescribed ACEI at end of study) |
| **HOMED-BP** | ACEI | - | Not reported | - | Not reported |
| | ARB | - | Not reported | - | Not reported |
| | CCB | - | Not reported | - | Not reported |
| **IDNT** | ARB | 4 | Mean number of drugs given | - | Not reported |
| | CCB | 4 | Mean number of drugs given | - | Not reported |
| **INSIGHT** | Diuretic | 1.72 | Minimum mean number of drugs given at year 3 | 78.1 | On randomized treatment at year 3 follow-up |
| | CCB | 1.65 | Minimum mean number of drugs given at year 3 | 70.7 | On randomized treatment at year 3 follow-up |
| **INVEST** | CCB | 1.4 | Mean number of drugs given at year 2 follow-up | 81.5 | On randomized treatment at year 2 follow-up (Step 1 drug only) |
| | non-CCB | 1.4 | Mean number of drugs given at year 2 follow-up | 77.5 | On randomized treatment at year 2 follow-up (Step 1 drug only) |
| **JMIC-B** | CCB | 2.02 | Mean number of drugs given | - | Not reported |
| | ACEI | 1.03 | Mean number of drugs given | - | Not reported |
| **LIFE** | ARB | 2.15 | Minimum mean number of drugs | 84.0 | On randomized treatment |
| | $\beta$-blocker | 2.11 | Minimum mean number of drugs | 80.0 | On randomized treatment |
| **MOSES** | CCB | 1.34 | Minimum mean number of drugs | - | Not reported |
| | ARB | 1.44 | Minimum mean number of drugs | - | Not reported |
| **NICS-EH** | Diuretic | 1.1 | Mean number of drugs given | - | Not reported |
| | CCB | 1.1 | Mean number of drugs given | - | Not reported |
| **NORDIL** | CCB | 1.4 | Mean number of drugs given | 77.0 | On randomized treatment |
| | $\beta$-blocker and/or diuretic | 1.5 | Mean number of drugs given | 93.0 | On randomized treatment |
| **ONTARGET** | ARB | 1.1 | Mean number of drugs given | 85.6 | On randomized treatment at year 4 follow-up |
| | ACEI | 1 | Mean number of drugs given | 84.7 | On randomized treatment at year 4 follow-up |
| | ARB and ACEI | 1.1 | Mean number of drugs given | 73.6 | On randomized treatment at year 4 follow-up |

| | | | | | |
|---|---|---|---|---|---|
| **STOP Hypertension-2** | $\beta$-blocker and/or diuretic | 1.6 | Mean number of drugs given | 62.3 | On randomized treatment at final visit |
| | ACEI | 1.6 | Mean number of drugs given | 61.3 | On randomized treatment at final visit |
| | CCB | 1.6 | Mean number of drugs given | 66.2 | On randomized treatment at final visit |
| **UKPDS** | ACEI | 27% | ≥3 agents in tight control | 78.0 | On randomized treatment at final visit |
| | $\beta$-blocker | 31% | ≥3 agents in tight control | 65.1 | On randomized treatment at final visit |
| **VALUE** | ARB-based | 2.1 | Mean number of drugs given | 73.7 | On randomized treatment during follow-up |
| | CCB-based | 2 | Mean number of drugs given | 74.9 | On randomized treatment during follow-up |
| **VHAS** | CCB | 1.2 | Mean number of drugs given | 44.1 | On randomized treatment (staying on monotherapy) at year 2 follow-up |
| | Diuretic | 1.3 | Mean number of drugs given | 38.8 | On randomized treatment (staying on monotherapy) at year 2 follow-up |

Trial name acronyms are described in full in the Trial acronym legend in the Supplement; SBP – systolic blood pressure; CCB – calcium channel-blocker; ACEI – angiotensin-converting enzyme inhibitor; ARB – angiotensin II receptor blocker; †Normotensive patient cohort only; ‡Hypertensive patient cohort only.

**Table S6. Characteristics of participants at baseline for each trial.**

| Trial | Participants, N (% women) | Age (years), mean (SD) | SBP (mmHg), mean (SD) | DBP (mmHg), mean (SD) | Body mass index (kg/m²), mean (SD) | % Current smokers (N) | % Caucasian / European (N) | % Prevalent disease (N) | | | | |
|---|---|---|---|---|---|---|---|---|---|---|---|---|
| | | | | | | | | Cardiovascular disease | Coronary heart disease | Cerebrovascular disease/stroke | Chronic kidney disease | Diabetes |
| AASK | 1094 (39) | 54 (11) | 150 (24) | 95 (10) | 30.6 (6.6) | 29 (321) | 0 (0) | 52 (564) | 52 (564) | - | 100 (1094) | 0 (0) |
| ABCD | 950 (39) | 58 (8) | 155 (17) | 98 (10) | 31.7 (5.7) | 14 (65) | 66 (310) | 9 (89) | 9 (41) | 2 (11) | 19 (90) | 100 (470) |
| ACCORD | 4733 (48) | 63 (7) | 139 (15) | 76 (10) | 32.1 (5.5) | 13 (626 | 59 (2781) | 34 (1593) | 14 (650) | 6 (307) | 0 (0) | 100 (4733) |
| ACTIVE | 9016 (39) | 70 (10) | 138 (17) | 82 (10) | 29.1 (5.8) | 8 (698) | 75 (6769) | 35 (3177) | 2294 (25.4%) | 14 (1230) | 0 (0) | 20 (1785) |
| ADVANCE | 11,140 (43) | 66 (6) | 145 (22) | 81 (10) | 28.3 (5.2) | 14 (1550) | 60 (6687) | 31 (3461) | 2380 (21.4%) | 13 (1438) | - | 100 (11,140) |
| ALLHAT | 42,418 (47 | 67 (8) | 146 (16) | 84 (10) | 29.6 (5.9) | 22 (9269) | 51 (21561) | 46 (19,389) | - | - | - | 36 (15,283) |
| ANBP | 3427 (37) | 50 (9) | 157 (15) | 100 (10) | 26.7 (3.9) | 26 (891) | - | 0 (16) | 0 (16) | 0 (0) | 7 (240) | 0 (0) |
| ANBP2 | 6083 (51) | 73 (5) | 168 (13) | 91 (10) | 27.1 (4.2) | 7 (431) | 98 (5984) | 8 (474) | 4 (229) | 5 (276) | 0 (6) | 7 (441) |
| ASCOT-BPLA | 19,257 (23) | 63 (9) | 164 (18) | 95 (10) | 28.7 (4.6) | 33 (6277) | 95 (18357) | 36 (7008) | 27 (5284) | 11 (2121) | 62 (12,017) | 27 (5145) |
| BENEDICT | 1204 (47) | 62 (8) | 151 (14) | 88 (10) | 29.1 (4.7) | 12 (146) | 100 (1204) | - | - | - | 0 (0) | 100 (1209) |
| CAMELOT | 1991 (26) | 58 (10) | 129 (16) | 78 (10) | 29.8 (5.3) | 26 (528) | 89 (1783) | 93 (1858) | 93 (1857) | 4 (81) | | 18 (364) |
| CAPPP | 10,985 (47) | 52 (8) | 161 (20) | 99 (10) | 27.8 (4.4) | 22 (2431) | - | 3 (351) | 2 (201) | 1 (160) | 0 (28) | 5 (572) |
| CARDIOSIS | 1111 (59) | 67 (7) | 158 (9) | 87 (10) | 27.8 (4.2) | 20 (226) | - | 18 (202) | 11 (128) | 8 (91) | 0 (0) | 0 (0) |
| CASE-J | 4703 (45) | 64 (11) | 163 (14) | 92 (10) | 24.5 (3.7) | 22 (1025) | 0 (0) | 22 (1028) | 13 (596) | 10 (473) | 58 (2720) | 43 (2018) |
| COLM | 5141 (48) | 74 (5) | 158 (13) | 87 (10) | 24.3 (3.4) | 11 (551) | 0 (0) | 24 (1225) | 11 (563) | 15 (751) | 2 (108) | 26 (1362) |
| CONVINCE | 16,476 (56) | 66 (7) | 150 (16) | 87 (10) | - | 23 (3795) | 84 (13845) | 27 (4458) | 23 (3820) | 6 (1019) | 0 (0) | 20 (3239) |
| COPE | 3293 (49) | 64 (11) | 154 (12) | 89 (10) | 24.5 (3.4) | 21 (700) | 0 (0) | 7 (219) | 3 (109) | 4 (126) | 46 (1530) | 0 (0) |
| DIABHYCAR | 4912 (30) | 65 (8) | 145 (15) | 82 (10) | 29.2 (4.6) | 15 (756) | - | 15 (739) | 15 (739) | - | - | 100 (4912) |
| Dutch TIA Trial | 1473 (36) | 64 (10) | 157 (25) | 91 (10) | - | 47 (693) | - | 100 (1473) | 9 (138) | 100 (1473) | 100 (1711) | 5 (79) |
| E-COST | 2048 (53) | 64 (11) | 164 (18) | 94 (10) | - | - | 0 (0) | 10 (213) | - | 10 (213) | 11 (228) | 0 (0) |
| ELSA | 2334 (46) | 57 (7) | 160 (15) | 98 (10) | 27.2 (3.8) | 20 (478) | 98 (2294) | 13 (305) | 13 (305) | - | 7 (163) | 4 (98) |
| EUROPA | 12,218 (15) | 61 (9) | 137 (16) | 82 (10) | 27.4 (3.5) | 15 (1862) | 99 (12,064) | 100 (12,218) | 100 (12,218) | 2 (222) | 0 (0) | - |
| EWPHE | 840 (70) | 71 (8) | 183 (17) | 101 (10) | 26.4 (4.5) | 17 (143) | - | 15 (124) | 9 (73) | 8 (63) | 0 (2) | 72 (8.6%) |
| HIJ-CREATE | 2049 (20) | 65 (9) | 135 (18) | 76 (10) | 24.6 (3) | 25 (509) | - | 100 (2049) | 100 (2049) | 10 (205) | 0 (3) | 780 (38.1%) |
| HOMED-BP | 3518 (50) | 60 (10) | 154 (18) | 90 (10) | 24.4 (3.5) | 21 (743) | 0 (0) | 3 (106) | 2 (61) | 1 (46) | - | 538 (15.3%) |
| HOPE | 9297 (27) | 66 (7) | 139 (20) | 79 (10) | 27.7 (4.4) | 14 (1319) | 90 (8343) | 80 (7477) | 80 (7477) | - | - | 3577 (38.5%) |
| HYVET | 3845 (60) | 84 (3) | 173 (9) | 91 (10) | 24.7 (3.7) | 7 (253) | - | 10 (374) | 3 (121) | 7 (261) | - | 388 (10.1%) |
| IDNT | 1715 (34) | 59 (8) | 159 (20) | 87 (11) | 30.8 (5.8) | - | 73 (1241) | - | - | - | 100 (1715) | 1715 (100%) |
| INSIGHT | 6321 (54) | 65 (6) | 173 (15) | 99 (10) | 28.2 (4.6) | 28 (1793) | - | 11 (671) | 11 (671) | - | - | 1302 (20.6%) |

| | | | | | | | | | | | | |
|---|---|---|---|---|---|---|---|---|---|---|---|---|
| INVEST | 21,320 (51) | 66 (10) | 151 (20) | 87 (10) | 29.2 (7.1) | 12 (2809) | 48 (10,925) | 100 (21,320) | 100 (21,320) | 7 (1629) | 2 (424) | 6400 (28.3%) |
| JMIC-B | 1650 (31) | 65 (85) | 146 (19) | 82 (10) | 24 (2.9) | 34 (563) | 0 | 100 (1650) | 100 (1650) | - | - | 372 (22.5%) |
| LIFE | 9193 (54) | 67 (7) | 174 (14) | 98 (10) | 28 (4.8) | 16 (1499) | 92 (8503) | 19 (1771) | 16 (1469) | 4 (401) | - | 1195 (13%) |
| MOSES | 1352 (46) | 68 (10) | 151 (18) | 87 (10) | 27.5 (4.3) | 18 (247) | 100 (1352) | 100 (1352) | 26 (355) | 100 (1352) | 5 (72) | 498 (36.8%) |
| NICS-EH | 414 (67) | 70 (7) | 172 (12) | 94 (10) | 23.4 (3.1) | 9 (38) | 0 | 4 (16) | 4 (0.9%) | 3 (12) | - | 17 (4%) |
| NORDIL | 10,881 (51) | 60 (7) | 173 (18) | 106 (10) | 27.8 (4.3) | 22 (2442) | - | 7 (740) | 496 (4.6%) | 2 (271) | 0 (31) | 727 (6.7%) |
| ONTARGET | 25,620 (27) | 67 (7) | 142 (17) | 82 (10) | 28.2 (4.8) | 13 (3225) | 73 (18,708) | 87 (22,315) | 75 (19,102) | 70 (17,927) | - | 9612 (37.5%) |
| PART 2 | 617 (18) | 60 (8) | 133 (17) | 79 (10) | 26.8 (3.6) | 16 (100) | 100 (615) | 74 (457) | 68 (420) | 10 (62) | - | 51 (8.3%) |
| PEACE | 8290 (18) | 64 (8) | 133 (17) | 77 (10) | - | 14 (1177) | - | 100 (8290) | 100 (8290) | 4 (357) | - | 1380 (16.6%) |
| PREVEND IT | 864 (35) | 51 (12) | 130 (18) | 76 (10) | 26.4 (4.4) | 40 (345) | 96 (830) | 3 (24) | 2 (13) | 2 (13) | 100 (864) | 22 (2.5%) |
| PREVENT | 825 (20) | 57 (10) | 129 (17) | 79 (10) | 28 (4.8) | 25 (204) | 89 (732) | 100 (825) | 100 (825) | 7 (55) | 1 (8) | 98 (11.9%) |
| PROFESS | 19,798 (36) | 66 (8) | 144 (17) | 84 (11) | 26.8 (5) | 21 (4231) | 57 (11,200) | 100 (19,798) | 15 (2933) | 100 (19,798) | - | 28 (5587) |
| PROGRESS | 6105 (30) | 64 (10) | 147 (19) | 86 (10) | 25.7 (3.8) | 21 (1279) | 0 (9) | 100 (6105) | 16 (983) | 100 (6105) | - | 761 (12.5%) |
| SHEP | 4736 (57) | 72 (7) | 170 (9) | 77 (10) | 27.1 (4.8) | 13 (597) | 79 (3731) | 6 (284) | 5 (232) | 0 (66) | 0 (0) | 476 (10.1%) |
| SPRINT | 9361 (36) | 68 (9) | 140 (15.6) | 78 (10) | 29.9 (5.8) | 13 (1240) | 58 (5399) | 20 (1877) | 20 (1877) | 0 (0) | 28 (2646) | 0 (0%) |
| STOP Hypertension 2 | 6614 (67) | 76 (4) | 194 (15) | 98 (10) | 26.7 (4) | 9 (594) | - | 16 (1072) | 10 (647) | 8 (502) | - | 719 (10.9%) |
| SYST-EUR | 4695 (67) | 70 (7) | 174 (10) | 86 (10) | 27 (4.1) | 7 (343) | - | 6 (286) | 3 (164) | 3 (124) | 0 (20) | 449 (9.6%) |
| TRANSCEND | 5926 (43) | 68 (7) | 141 (17) | 82 (10) | 28.2 (4.8) | 10 (582) | 61 (3621) | 88 (5222) | 75 (4418) | 71 (4207) | - | 2118 (35.7%) |
| UKPDS | 1148 (45) | 56 (8) | 159 (17) | 93 (10) | 29.6 (5.5) | 23 (256) | 87 (1000) | 3 (35) | 1 (15) | 2 (20) | - | 1148 (100%) |
| VALISH | 3079 (62) | 76 (4) | 169 (9) | 82 (10) | 23.5 (3.4) | 15 (450) | - | 12 (371) | 6 (194) | 7 (202) | 31 (944) | 399 (13%) |
| VALUE | 15,245 (42) | 67 (8) | 154 (19) | 87 (10) | 28.6 (5) | 24 (3664) | 89 (13,617) | 60 (9169) | 46 (6981) | 20 (3014) | - | 4823 (31.6%) |
| VHAS | 1414 (51) | 54 (7) | 169 (10) | 102 (10) | 27.1 (4.1) | 18 (256) | 100 (1414) | 12 (164) | - | - | 3 (48) | 108 (7.6%) |

Acronyms are described in full in the Trial acronym legend in the Supplement; SBP – systolic blood pressure; DBP – diastolic blood pressure.

**Table S7. Blood pressure measurement methods used in the trials included in the BPLTTC.**

| Study | Specified method of blood pressure measurement |
|---|---|
| AASK | Mean of last two readings of three consecutive seated BP readings after five minutes rest. A Hawksley random zero sphygmomanometer was used. |
| ABCD | Determined at two separate visits. |
| ACCORD | Average of three measurements in the sitting position, using an automated device (Omron 907), after five minutes rest. |
| ACTIVE I | Measured in duplicated in five minute intervals. |
| ADVANCE | Mean of two measurements in the sitting position, taken using a standardized automated sphygmomanometer (Omron HEM-705CP,Tokyo, Japan), after the patient was rested for at least five minutes. |
| ALLHAT | Mean of four measurements, two taken at two separate visits. |
| ANBP | Mean of four measurements over two visits, taken using a random zero or London School of Hygiene sphygmomanometer in sitting position after 5 minutes of rest. |
| ANBP2 | Mean of measurements at two visits, taken using a mercury sphygmomanometer in the sitting position. |
| ASCOT-BPLA | Measured three times using a semi-automated device in the sitting position, after five minutes of rest. Mean of last two readings used. |
| BENEDICT | Mean of three morning measurements. |
| CAMELOT | Measured using a manual cuff and stethoscope. |
| CAPPP | Mean of two measurements taken in supine position using conventional mercury sphygmomanometer. |
| Cardio-Sis | Mean of three consecutive readings at every visit, taken using standard mercury sphygmomanometers after sitting for ten minutes. |
| CASE-J | Mean of two measurements, taken in the sitting position. |
| COLM | Mean of two stable measurements that differed by less than 5 mm Hg, taken at least twice at intervals of one to two minutes. |
| CONVINCE | Not provided. |
| COPE | Mean of two stable measurements (difference <5mmHg), taken using the upper arm after five minutes rest in a sitting position. |
| DIABHYCAR | Measured once using a mercury sphygmomanometer in the sitting position. |
| Dutch TIA | A single measurement in the sitting position using a mercury manometer. |
| E-COST | Taken in sitting position. |
| ELSA | Three measurements taken in sitting position using a mercury manometer. |
| EUROPA | Two measurements taken with a standard sphygmomanometer in sitting position, after five minutes rest. |
| EWPHE | Taken in sitting position. |
| HIJ-CREATE | Measured using a standard cuff mercury sphygmomanometer in sitting position after five minutes of rest. |
| HOMED-BP | Clinic: Mean of two measurements taken using a validated sphygmomanometer (OMRON HEM-907IT, Omron Healthcare, Kyoto, Japan) in the sitting position after two minutes rest; Home: as with clinic but taken in the morning, 1 hour after waking up, before breakfast and before taking BP treatment, and mean of readings over 5 days prior to clinic visit. |
| HOPE | Mean of two measurements in the sitting position after fifteen minutes rest, taken using a mercury sphygmomanometer. |
| HYVET | Two measurements taken in the standing position, using a mercury sphygmomanometer or a validated automated device. |
| IDNT | Taken in the sitting position. |
| INSIGHT | Mean of three measurements after a five minute rest, taken using a mercury sphygmomanometer. |
| INVEST | Mean of two measurements taken in the sitting position as described in JNC VI. |
| JMIC-B | Average of two last measurements of three measurements, taken in sitting or supine position, whichever had been decided upon initially. |
| LIFE | Sitting blood pressure, measured at the trough. |
| MOSES | Not provided. |
| NICS-EH | Not provided. |
| NORDIL | Measured on two separate occasions. |

| | |
|---|---|
| ONTARGET | Average of two readings was used. Each blood pressure reading consisted of measurement using an automated validated device (OMRON, model HEM-757) after 3 minutes of rest while sitting. |
| PART 2 | Average of two measurements using a standard mercury sphygmomanometer following standard protocol. |
| PEACE | Not provided. |
| PREVEND IT | Ten consecutive measurements were taken using an automatic Dinamap XL model; last two measurements were averaged to determine BP measurement. |
| PREVENT | Not provided. |
| PROFESS | Average of two measures taken twice, at least two minutes apart, using a standard and validated Omron sphygmomanometer (Omron Healthcare Inc) with an appropriately sized cuff applied to the upper nondominant arm at heart level.[44 83] |
| PROGRESS | Two measurements of BP were taken using a mercury sphygmomanometer. |
| SHEP | Average of four seated BP measurements, taken using a Hawksley random zero manometer. |
| SPRINT | Measured after rest, preferably using an automated device, but a manual device could be used if necessary. |
| STOP Hypertension-2 | After five minutes of rest, BP was measured in the supine position. |
| Syst-Eur | Average of six sitting BP measurements and six standing BP measurements, two in each position at each of three baseline visits taken one month apart. |
| TRANSCEND | Average of two readings was used. Each blood pressure reading consisted of measurement using an automated validated device (OMRON, model HEM-757) after 3 minutes of rest while sitting. |
| UKPDS | Mean of last three of four BP measurements taken after 5 minutes of rest while sitting at consecutive clinic visits using an automated validated device (Copal UA-251 or a Takeda UA-751). |
| VALISH | Blood pressure taken in sitting position at two separate visits within 2 to 4 weeks. |
| VALUE | Measured after sitting for five minutes. |
| VHAS | Lowest value of three measurements, taken in the sitting position after a 10-minute rest, at consecutive one-minute intervals. |

Acronyms are described in full in the Trial acronym legend in the Supplement; BP – blood pressure.

**Table S8**. **Supplementary data for Figure 1 and online Figure S2, showing estimated mean blood pressure separately for each comparison arm at specific time points during follow-up, by trial design.**

| Comparison arm | Follow-up period (months) | | | | | | | | | | | | |
|---|---|---|---|---|---|---|---|---|---|---|---|---|---|
| | Baseline | 3 | 6 | 9 | 12 | 18 | 24 | 30 | 36 | 42 | 48 | 54 | 60 |
| **Blood pressure-lowering intensity trials** | | | | | | | | | | | | | |
| Systolic blood pressure (mmHg) | | | | | | | | | | | | | |
| More intense | 148 | 138 | 135 | 133 | 131 | 129 | 127 | 126 | 127 | 127 | 128 | 130 | 131 |
| Less intense | 148 | 143 | 142 | 141 | 141 | 140 | 140 | 139 | 140 | 140 | 140 | 140 | 140 |
| Diastolic blood pressure (mmHg) | | | | | | | | | | | | | |
| More intense | 83 | 77 | 75 | 74 | 73 | 72 | 71 | 70 | 70 | 70 | 70 | 71 | 71 |
| Less intense | 82 | 78 | 78 | 77 | 77 | 76 | 76 | 75 | 75 | 75 | 75 | 75 | 75 |
| **Placebo-controlled trials** | | | | | | | | | | | | | |
| Systolic blood pressure (mmHg) | | | | | | | | | | | | | |
| Active | 146 | 140 | 138 | 137 | 136 | 135 | 134 | 134 | 134 | 135 | 136 | 136 | 137 |
| Placebo | 146 | 143 | 143 | 142 | 141 | 141 | 140 | 140 | 140 | 140 | 140 | 141 | 140 |
| Diastolic blood pressure (mmHg) | | | | | | | | | | | | | |
| Active | 83 | 80 | 79 | 79 | 78 | 78 | 77 | 77 | 77 | 77 | 77 | 77 | 77 |
| Placebo | 83 | 82 | 82 | 81 | 81 | 80 | 80 | 80 | 79 | 79 | 79 | 79 | 79 |
| **Blood pressure difference trials** | | | | | | | | | | | | | |
| Systolic blood pressure (mmHg) | | | | | | | | | | | | | |
| Active | 146 | 139 | 137 | 136 | 135 | 133 | 132 | 132 | 133 | 133 | 134 | 135 | 136 |
| Placebo | 146 | 143 | 142 | 141 | 141 | 140 | 139 | 139 | 139 | 140 | 140 | 140 | 140 |
| Diastolic blood pressure (mmHg) | | | | | | | | | | | | | |
| Active | 83 | 80 | 79 | 78 | 77 | 76 | 76 | 76 | 76 | 76 | 76 | 76 | 76 |
| Placebo | 83 | 81 | 81 | 81 | 80 | 79 | 79 | 79 | 79 | 79 | 78 | 78 | 78 |
| **Drug class comparison trials** | | | | | | | | | | | | | |
| Systolic blood pressure (mmHg) | | | | | | | | | | | | | |
| Active | 156 | 150 | 148 | 146 | 144 | 141 | 139 | 139 | 138 | 139 | 139 | 140 | 140 |
| Control | 155 | 149 | 147 | 145 | 144 | 141 | 140 | 139 | 139 | 139 | 139 | 140 | 140 |
| Diastolic blood pressure (mmHg) | | | | | | | | | | | | | |
| Active | 90 | 86 | 85 | 84 | 83 | 81 | 80 | 80 | 79 | 79 | 79 | 79 | 79 |
| Control | 89 | 86 | 85 | 84 | 83 | 81 | 80 | 80 | 79 | 79 | 79 | 79 | 79 |

Estimates based on separate models for each comparison arm, with random intercepts at individual and trial levels, a random slope for time at the individual level (see Method for details) and adjusted for baseline blood pressure, age and sex; Data concatenated up to five years of follow-up.

**Table S9**. **Supplementary data for Figure 2 and online Figure S3 showing mean blood pressure difference between comparison groups over follow-up time.**

| Follow-up period, months | Systolic blood pressure (mmHg) difference (95% CI) | Diastolic blood pressure (mmHg) difference (95% CI) |
|---|---|---|
| Blood pressure-lowering intensity trials (8 trials) | | |
| 0 to 6 | -5.4 (-5.6 to -5.1) | -2.7 (-2.8 to -2.6) |
| 6 to 12 | -10.2 (-10.5 to -9.8) | -5.3 (-5.5 to -5.1) |
| 12 to 24 | -11.0 (-11.3 to -10.8) | -5.6 (-5.8 to -5.4) |
| 24 to 36 | -12.1 (-12.5 to -11.8) | -6.2 (-6.4 to -6.0) |
| 36 to 48 | -12.1 (-12.5 to -11.7) | -6.1 (-6.3 to -5.8) |
| 48 to 60 | -11.1 (-11.7 to -10.5) | -5.5 (-5.8 to -5.1) |
| Placebo-controlled trials (20 trials) | | |
| 0 to 6 | -3.0 (-3.1 to -2.9) | -1.5 (-1.6 to -1.4) |
| 6 to 12 | -5.6 (-5.8 to -5.4) | -2.7 (-2.8 to -2.5) |
| 12 to 24 | -5.3 (-5.5 to -5.1) | -2.5 (-2.6 to -2.4) |
| 24 to 36 | -5.3 (-5.5 to -5.0) | -2.4 (-2.5 to -2.3) |
| 36 to 48 | -4.9 (-5.2 to -4.7) | -2.2 (-2.3 to -2.0) |
| 48 to 60 | -4.5 (-4.8 to -4.2) | -1.9 (-2.1 to -1.8) |
| All blood pressure difference trials (28 trials) | | |
| 0 to 6 | -3.5 (-3.6 to -3.4) | -1.8 (-1.8 to -1.7) |
| 6 to 12 | -6.6 (-6.7 to -6.4) | -3.2 (-3.3 to -3.1) |
| 12 to 24 | -6.4 (-6.6 to -6.3) | -3.1 (-3.2 to -3.0) |
| 24 to 36 | -6.7 (-6.9 to -6.6) | -3.2 (-3.4 to -3.1) |
| 36 to 48 | -6.4 (-6.6 to -6.2) | -3.0 (-3.1 to -2.9) |
| 48 to 60 | -5.7 (-5.9 to -5.4) | -2.6 (-2.7 to -2.4) |
| Drug class comparison trials (29 trials) | | |
| 0 to 6 | -1.0 (-1.1 to -1.0) | -0.4 (-0.4 to -0.3) |
| 6 to 12 | -1.9 (-2.1 to -1.8) | -0.7 (-0.8 to -0.6) |
| 12 to 24 | -1.6 (-1.8 to -1.5) | -0.8 (-0.8 to -0.7) |
| 24 to 36 | -1.5 (-1.6 to -1.3) | -0.7 (-0.7 to -0.6) |
| 36 to 48 | -1.5 (-1.7 to -1.4) | -0.8 (-0.9 to -0.7) |
| 48 to 60 | -1.6 (-1.8 to -1.5) | -1.0 (-1.1 to -0.9) |

Model based on fixed treatment effect (see Method), and estimates adjusted for baseline blood pressure, age and sex; Negative values indicate lower blood pressure in the active than in the control group.

**Table S10. Supplementary data for Figure 2 and online Figure S3 showing achieved mean blood pressure difference between active and control groups with and without taking into account early follow-up measurements.**

| Follow-up period, months | Systolic blood pressure (mmHg) difference (95% CI) | Diastolic blood pressure (mmHg) difference (95% CI) |
|---|---|---|
| Blood pressure-lowering intensity trials (8 trials) | | |
| All follow-up | -8.3 (-8.4 to -8.1) | -3.9 (-4.1 to -3.8) |
| From 12 months to end of follow-up | -11.2 (-11.4 to -11.0) | -5.6 (-5.8 to -5.5) |
| Placebo-controlled trials (20 trials) | | |
| All follow-up | -4.0 (-4.1 to -3.9) | -1.9 (-2.0 to -1.8) |
| From 12 months to end of follow-up | -5.1 (-5.3 to -5.0) | -2.3 (-2.4 to -2.2) |
| Blood pressure difference trials (29 trials) | | |
| All follow-up | -4.8 (-4.9 to -4.7) | -2.3 (-2.4 to -2.3) |
| From 12 months to end of follow-up | -6.3 (-6.5 to -6.2) | -3.0 (-3.1 to -2.9) |
| Drug class comparison trials (29 trials) | | |
| All follow-up | -1.4 (-1.5 to -1.3) | -0.5 (-0.6 to -0.5) |
| From 12 months to end of follow-up | -1.4 (-1.5 to -1.3) | -0.6 (-0.7 to -0.6) |

Model based on fixed treatment effect and random intercept for trials (see Method for details) and adjusted for baseline blood pressure, age and sex; Negative values indicate lower blood pressure in the active than in the control group.

**Table S11. Comparison of models estimating mean blood pressure difference between comparison arms.**

| Model | SBP (mmHg) | | DBP (mmHg) | |
|---|---|---|---|---|
| | Mean difference (95% CI) | AIC | Mean difference (95% CI) | AIC |
| **BLOOD PRESSURE-LOWERING INTENSITY TRIALS** | | | | |
| ***All follow-up*** | | | | |
| Model 1: Fixed effect (treatment and time) and random intercept (trial) | -10.4 (-10.5 to -10.3) | 3,033,977 | -5.3 (-5.4 to -5.3) | 2,680,852 |
| Model 2: Random intercepts (trial and individual levels), fixed treatment effect, fixed time effect (cubic term) and random slope for time (individual level) | -8.3 (-8.5 to -8.1) | 2,957,226 | -3.9 (-4.1 to -3.8) | 2,570,953 |
| ***From 12 months to end of follow-up*** | | | | |
| Model 1: Fixed effect (treatment and time) and random intercept (trial) | -12.1 (-12.2 to -12.0) | 1,998,991 | -6.2 (-6.3 to -6.1) | 1,780,048 |
| Model 2: Random intercepts (trial and individual levels), fixed treatment effect, fixed time effect (cubic term) and random slope for time (individual level) | -11.2 (-11.4 to -11.0) | 1,942,811 | -5.6 (-5.8 to -5.5) | 1,696,790 |
| **PLACEBO-CONTROLLED TRIALS** | | | | |
| ***All follow-up*** | | | | |
| Model 1: Fixed effect (treatment and time) and random intercept (trial) | -4.6 (-4.6 to -4.5) | 7,326,530 | -2.1 (-2.1 to -2.1) | 6,313,968 |
| Model 2: Random intercepts (trial and individual levels), fixed treatment effect, fixed time effect (cubic term) and random slope for time (individual level) | -4.0 (-4.1 to -3.9) | 7,101,069 | -1.9 (-2.0 to -1.8) | 6,114,559 |
| ***From 12 months to end of follow-up*** | | | | |
| Model 1: Fixed effect (treatment and time) and random intercept (trial) | -5.1 (-5.2 to -5.0) | 4,224,464 | -2.3 (-2.4 to -2.3) | 3,642,401 |
| Model 2: Random intercepts (trial and individual levels), fixed treatment effect, fixed time effect (cubic term) and random slope for time (individual level) | -5.1 (-5.3 to -5.0) | 4,082,606 | -2.3 (-2.4 to -2.2) | 3,515,852 |
| **ALL BLOOD PRESSURE DIFFERENCE TRIALS** | | | | |
| ***All follow-up*** | | | | |
| Model 1: Fixed effect (treatment and time) and random intercept (trial) | -6.0 (-6.1 to -6.0) | 11,550,184 | -3.0 (-3.0 to -2.9) | 10,018,329 |
| Model 2: Random intercepts (trial and individual levels), fixed treatment effect, fixed time effect (cubic term) and random slope for time (individual level) | -4.8 (-4.9 to -4.7) | 11,174,395 | -2.3 (-2.4 to -2.3) | 9,647,478 |
| ***From 12 months to end of follow-up*** | | | | |
| Model 1: Fixed effect (treatment and time) and random intercept (trial) | -7.1 (-7.2 to -7.1) | 6,690,469 | -3.5 (-3.5 to -3.4) | 5,815,605 |
| Model 2: Random intercepts (trial and individual levels), fixed treatment effect, fixed time effect (cubic term) and random slope for time (individual level) | -6.3 (-6.4 to -6.2) | 6,438,360 | -3.0 (-3.0 to -2.9) | 5,561,642 |
| **DRUG COMPARISON TRIALS** | | | | |
| ***All follow-up*** | | | | |
| Model 1: Fixed effect (treatment and time) and random intercept (trial) | -1.4 (-1.4 to -1.3) | 16,539,815 | -0.6 (-0.6 to -0.6) | 14,228,432 |
| Model 2: Random intercepts (trial and individual levels), fixed treatment effect, fixed time effect (cubic term) and random slope for time (individual level) | -1.4 (-1.5 to -1.3) | 16,078,396 | -0.5 (-0.6 to -0.5) | 13,790,959 |
| ***From 12 months to end of follow-up*** | | | | |
| Model 1: Fixed effect (treatment and time) and random intercept (trial) | -1.3 (-1.4 to -1.3) | 10,516,160 | -0.7 (-0.7 to -0.6) | 9,069,610 |
| Model 2: Random intercepts (trial and individual levels), fixed treatment effect, fixed time effect (cubic term) and random slope for time (individual level) | -1.4 (-1.5 to -1.3) | 10,193,302 | -0.6 (-0.7 to -0.6) | 8,751,146 |

SBP – systolic blood pressure; DBP – diastolic blood pressure; AIC: Aikeke information criterion; Adjusted for baseline blood pressure, age and sex; Negative values indicate lower blood pressure in the active than in the control group.

**TRIAL ACRONYM LEGEND**

| Trial acronym | Full name or description |
|---|---|
| **AASK** | African American Study of Kidney Disease and Hypertension |
| **ABCD** | Appropriate Blood Pressure Control in Diabetes |
| **ACCORD** | Action to Control Cardiovascular Risk in Diabetes blood pressure trial |
| **ACTIVE I** | Atrial Fibrillation Clopidogrel Trial with Irbesartan for Prevention of Vascular Events |
| **ADVANCE** | Action in Diabetes and Vascular disease: preterAx and diamicroN-MR Controlled Evaluation |
| **ALLHAT** | Antihypertensive and Lipid Lowering Treatment to Prevent Heart Attack Trial |
| **ANBP** | Australian National Blood Pressure Study |
| **ANBP2** | Second Australian National Blood Pressure Study |
| **ASCOT-BPLA** | Anglo-Scandinavian Cardiac Outcomes Trial-Blood Pressure Lowering Arm |
| **BENEDICT** | BErgamo NEphrologic DIabetes Complications Trial |
| **CAMELOT** | Comparison of Amlodipine vs Enalapril to Limit Occurrences of Thrombosis |
| **CAPPP** | Captopril Prevention Project |
| **Cardio-Sis** | Studio Italiano Sugli Effetti Cardiovascolari del Controllo della Pressione Arteriosa Sistolica |
| **CASE-J** | Candesartan Antihypertensive Survival Evaluation in Japan Trial |
| **COLM** | Combination of OLMesartan study |
| **CONVINCE** | Controlled ONset Verapamil INvestigation of Cardiovascular Endpoints trial |
| **COPE** | Combination Therapy of Hypertension to Prevent Cardiovascular Events |
| **DIABHYCAR** | Noninsulin-dependent diabetes, hypertension, microalbuminuria or proteinuria, cardiovascular events, and ramipril |
| **Dutch TIA Trial** | Dutch Transient Ischemic Attack Trial |
| **E-COST** | Efficacy of Candesartan on Outcome in Saitama Trial |
| **ELSA** | European Lacidipine Study on Atherosclerosis |
| **EUROPA** | EUropean trial on Reduction Of cardiac events with Perindopril in patients with stable coronary Artery disease |
| **EWPHE** | European Working Party on High Blood Pressure in the Elderly |
| **HIJ-CREATE** | Heart Institute of Japan Candesartan Randomized Trial for Evaluation in Coronary Artery Disease |
| **HOMED-BP** | Hypertension Objective Treatment Based on Measurement by Electrical Devices of Blood Pressure |
| **HOPE** | Heart Outcomes Prevention Evaluation |
| **HYVET** | Hypertension in the Very Elderly Trial |
| **IDNT** | Irbesartan Diabetic Nephropathy Trial |
| **INSIGHT** | International Nifedipine GITS study: Intervention as a Goal in Hypertension Treatment |
| **INVEST** | International Verapamil-Trandolapril Study |
| **JMIC-B** | Japan Multicenter Investigation for Cardiovascular Diseases-B |
| **LIFE** | Losartan Intervention For Endpoint reduction |
| **MOSES** | Morbidity and Mortality After Stroke, Eprosartan Compared With Nitrendipine for Secondary Prevention |
| **NICS-EH** | National Intervention Cooperative Study in Elderly Hypertensives |
| **NORDIL** | Nordic Diltiazem Study |
| **ONTARGET** | Ongoing Telmisartan Alone and in Combination with Ramipril Global Endpoint Trial |
| **PART 2** | Prevention of Atherosclerosis with Ramipril Trial |
| **PEACE** | Prevention of Events with Angiotensin Converting Enzyme Inhibition |
| **PREVEND IT** | Prevention of Renal and Vascular Endstage Disease Intervention Trial |
| **PREVENT** | Prospective Randomized Evaluation of the Vascular Effects of Norvasc Trial |
| **PROFESS** | Prevention Regimen for Effectively Avoiding Second Strokes |
| **PROGRESS** | Perindopril Protection Against Recurrent Stroke Study |
| **SHEP** | Systolic Hypertension in the Elderly Program |
| **SPRINT** | Systolic Blood Pressure Intervention Trial |
| **STOP Hypertension-2** | Swedish Trial in Old Patients with Hypertension-2 |
| **Syst-Eur** | Systolic Hypertension in Europe |
| **TRANSCEND** | Telmisartan Randomized Assessment Study in ACE Intolerant Subjects with Cardiovascular Disease |
| **UKPDS** | UK Prospective Diabetes Study |
| **VALISH** | Valsartan in Elderly Isolated Systolic Hypertension |
| **VALUE** | Valsartan Antihypertensive Long-term Use Evaluation |
| **VHAS** | Verapamil in Hypertension and Atherosclerosis Study |
